# What is the effectiveness and cost-effectiveness of at-home time-limited reablement service for improving an individual’s independence and health outcomes and reducing the need for long term care: a rapid review

**DOI:** 10.1101/2025.06.13.25329562

**Authors:** Bethany Anthony, Jacob Davies, Kalpa Pisavadia, Sofie Roberts, Greg Flynn, Limssy Varghese, Rachel Granger, Madison Montgomery, Rebecca Rosi, Elizabeth Gillen, Juliet Hounsome, Dyfrig Hughes, Jane Noyes, Deborah Fitzsimmons, Rhiannon Tudor Edwards, Adrian Edwards, Alison Cooper, Ruth Lewis

## Abstract

Local authorities and health services in Wales are tasked with reablement, to help individuals who are at risk of frailty maintain and improve independence. However, social care resource constraints, mean that the balance of funds and workforce may be focussed on acute health care and long-term care in the community. This review aimed to identify evidence on the effectiveness and cost-effectiveness of at-home time-limited reablement services for improving an individuals independence and health outcomes and reducing the need for long term care.

Eighteen studies were included in the review: 16 reported on clinical effectiveness and three on cost-effectiveness (one study reported on both). Key clinical-effectiveness findings included person-related and service-level outcomes.

**Implications for practice:** There is international evidence that reablement led by an allied health professional (AHP), such as occupational therapists, or a multi-professional team that includes allied health professionals (AHPs) is effective in improving mobility including activities of daily living (ADL), quality of life and falls outcomes.

There is also evidence that demonstrated that reablement led by AHPs can reduce the need for long term care in terms of the use of domiciliary care services and admissions to residential care.

Three economic evaluations found reablement services to be cost-effective, although there were some methodological flaws that limited the certainty of findings.

There is a need for more studies from a UK perspective - only three UK-based studies were identified, with the rest reflecting an international body of evidence.

**Report Contributors:** *Review Team:* Bethany Anthony, Jacob Davies, Kalpa Pisavadia, Sofie Roberts, Greg Flynn, Limssy Varghese, Rachel Granger, Madison Montgomery, Rebecca Rosi, Elizabeth Gillen, Juliet Hounsome, Dyfrig Hughes, Jane Noyes, Deborah Fitzsimmons, Rhiannon Tudor Edwards.

*Methodological Advice:* Ruth Lewis

*Evidence Centre Team:* Adrian Edwards, Ruth Lewis, Alison Cooper, Elizabeth Doe and Micaela Gal involved in stakeholder engagement, review of report and editing.

*Public Partner:* Rashmi Kumar

*Stakeholder(s):* Denise Moultrie, Ruth Crowder, Kerrie Phipps, and Rhian Matthews

*Disclaimer:* The views expressed in this publication are those of the authors, not necessarily Health and Care Research Wales. The Health and Care Research Wales Evidence Centre and authors of this work declare that they have no conflict of interest.

**EXECUTIVE SUMMARY:** *What is a Rapid Review?:* Our rapid reviews use a variation of the systematic review approach, abbreviating or omitting some components to generate evidence to inform stakeholders promptly whilst maintaining attention to bias.

*Who is this Rapid Review for?:* The review question was suggested by Health, Social Care and Early Years, Welsh Government.

*Background / Aim of Rapid Review:* Reablement is defined by NICE (2017) as a community-based intervention that aims to increase a service user’s independence by helping them recover lost skills and confidence. Local authorities and health services in Wales are tasked with reablement, to help individuals who are at risk of frailty maintain and improve independence. However, social care resource constraints, mean that the balance of funds and workforce may be focussed on acute health care and long-term care in the community. The review aimed to identify evidence on the effectiveness and cost-effectiveness of at -home time-limited reablement services for improving an individual’s independence and health outcomes and reducing the need for long term care.

*Results of the Rapid Review:* The evidence base
The review included evidence available up until December 2024.
Eighteen studies were included: 16 reported on clinical effectiveness (11 were controlled trials, 4 uncontrolled before and after studies, and one controlled cohort study); and three on cost-effectiveness (one study reported on both). Ten studies evaluated a step-up reablement model, two studies focused on the step-down reablement, four studies evaluated a reablement both step-up and step-down reablement, whilst the focus was unclear for the remaining two studies. Step-up reablement is defined as early / preventative intervention when an aspect of functioning deteriorates, following injury (e.g. fall) or a period of ill health and mitigating risk of hospital admission. Step-down reablement is the intervention received immediately following discharge with a view to regaining function.
Twelve studies evaluated a reablement model led by a multi-professional team, which included allied health professionals (AHPs).^1^ Five studies evaluated an AHP-led reablement model. One study evaluated a reablement model led by a nurse case manager coordinator. The reablement intervention was delivered by a range of people including: an occupational therapist (n=1 study); a member of the multiprofessional team or multi-professional workforce (n=10); a multi-professional team consisting of a range of different AHPs only (n=1); or non-health professional such as care manager, homecare personnel, health care assistant, home care aid, or carer (n=6).

*Key clinical-effectiveness findings:* Person-related outcomes
A significant amount of evidence on the effectiveness of reablement interventions on person-related outcomes was identified. Reablement interventions were **effective in improving people’s mobility and their ability to undertake activities of daily living (ADL), increasing quality of life, and reducing falls**. Reablement interventions may be effective in improving individuals coping, in terms of sense of coherence (how individuals perceive life as comprehensible, manageable & meaningful). Reablement was not found to be effective for improving grip strength or increasing clients’ social support. Service-level outcomes
A significant amount of evidence on the effectiveness of reablement interventions on service-level outcomes was identified. Reablement was **effective in reducing** the need for **long term home care services** and **residential care admissions**. Reablement was also effective in reducing the number of **outpatient treatments** compared with usual domiciliary care.
There were contradictory findings on reablement’s effectiveness in reducing hospital admissions, community care service use, and social care service use. There were inconsistent findings on reablement’s effectiveness in reducing emergency department visits. Type of reablement model
One study conducted in England found that a reablement model that was led and delivered by occupational therapists resulted in greater improvements in activities of daily living, quality of life, and falls, compared to reablement led and delivered by social care workers. Although the sample size was small and findings were not statistically significant, the Occupational Therapy-led model showed promising trends. In terms of community and social care resource use, participants in the Occupational Therapy-led reablement group were less likely to use health services, including GP visits and community support such as meals at home, suggesting potential efficiency benefits compared to the social care worker–led model. Key cost-effectiveness findings
Three economic evaluations found **reablement services to be cost-effective**, although there were some methodological flaws in the studies, that limited the certainty of findings.

*Research Implications and Evidence Gaps:*

- There is a need for more studies from a UK perspective - only three UK-based studies were identified, with the rest reflecting an international body of evidence. While the international research is useful when it aligns with how reablement services are delivered in the UK, it is essential to consider the unique context of Wales where reablement services are often hosted within or delivered by local authorities. Noting there is currently significant challenge reported by occupational therapists in local authority in undertaking research activity.
- The economic evaluations had methodological flaws that limited the certainty of review findings, evidencing a need for future economic evaluations on the topic.

*Policy and Practice Implications:*

- There is international evidence that reablement led by a multi-professional team that includes allied health professionals (AHPs)^1^, such as occupational therapists, is effective in improving mobility including activities of daily living, quality of life and falls outcomes.
- There is also evidence that demonstrated that reablement led by AHPs can reduce the need for long term care in terms of the use of domiciliary care services and admissions to residential care.
- While not all reablement services currently utilise AHPs, the evidence from this review makes a strong case for their inclusion in Wales. AHP-led reablement has been shown to improve person-centred outcomes and reduce the need for long-term care, supporting the case for targeted investment in AHP roles within reablement services.
- Three economic evaluations identified reablement as a cost-effective alternative to domiciliary care services.
- Given UK’s ageing population, the evidence from the Science Evidence Advice (SEA) 2023 report of the future prevalence and impact of frailty, and the high costs associated with ongoing care needs for people at risk of frailty, our findings make the case for investing in time-limited reablement interventions in Wales.

*Economic considerations:*

- Reablement programmes may provide cost savings to commissioners and the health and social care systems through prevention of or reduced length of hospital admission, reductions in hospital readmission and preventing or reducing domiciliary and residential care demand.
- Frailty has a sizeable impact on healthcare resource use in the UK. Total additional costs of frailty-related healthcare resource use are £8 billion per year when adjusted to 2025 prices.

## 1. BACKGROUND

### 1.1 Who is this review for?

This Rapid Review was conducted as part of the Health and Care Research Wales Evidence Centre Work Programme. The above question was suggested by Health, Social Services and Early Years, Welsh Government. The intended audience is Welsh Government, health boards and local authorities. This research will be used to help underpin the possible funding decisions and process developments and priorities. It will also be used to guide Welsh Government policy decisions regarding the provisions for reablement services.

### 1.2 Background and purpose of this review

Local authorities and health services in Wales are tasked with reablement, aimed at helping individuals regain independence when their functioning decreases and/or as a result of a hospital admission. However, due to social care resource constraints, funds often shift toward post-hospitalisation reablement only and/or toward long-term care, reducing resources for reablement. Traditional reablement research has focused on ‘step down’ reablement (support after hospital discharge). However, a growing interest in ‘step up’ preventative reablement has emerged. This involves proactive intervention for individuals at risk of deteriorating, aiming to prevent hospital admissions, improve health outcomes, and reduce long-term care costs. Many health boards and local authorities, stretched for resources, prioritise discharge support over preventative care at present, often caused by financial and staffing constraints. Identifying the financial benefits of reablement may help make a case for increased investment, aligning with community-based health strategies and reducing hospital admissions.

An agreed all Wales definition of reablement refers to an enabling approach with services that provide rehabilitation for all people with physical or mental disabilities. It helps them adapt to their condition by learning or re-learning the skills needed to function in everyday life. The focus is on promoting and optimising functional independence by practicing activities, rather than interventions aimed at improving underlying impairments (All Wales Rehabilitation Framework, 2022).

Reablement typically lasts for up to six weeks, with a focus on achieving specific goals within that timeframe. Delivered in the individual’s own home, wherever that may be, allowing for practical application of skills in familiar surroundings. A personalised plan is created with the individual, focusing on restoring function and independence and improving their ability to perform daily tasks that matter to them. Helping individuals become more self-sufficient and confident in their abilities, rather than relying on ongoing support. A multi-professional approach is required to provide a holistic approach to care, usually comprising of allied health professional-led (AHP-led) delivery, occupational therapists, reablement assessors (if not the occupational therapist), physiotherapists, social workers, reablement support workers and rehabilitation or therapy support workers. In some areas, other workforce may be included such as home care or domiciliary care workers. The reablement service is often hosted within or delivered by local authorities, however there are currently different models of reablement service provision in Wales and not all include allied health professionals (AHPs).

The aim of this review was to identify evidence on the effectiveness and cost-effectiveness of at-home time-limited reablement services for improving an individual’s independence and health outcomes and reducing the need for long term care.

### 1.3 NICE guidance on reablement services

This section summarises the NICE 2017 guidance (NG74) on intermediate care, including reablement. The NICE guidelines referring specifically to reablement will be the primary focus of this summary and will be considered throughout this rapid review.

‘Reablement’ is referred to by NICE (2017) as a community-based intervention that aims to increase a service user’s independence by helping them recover lost skills and confidence. NICE guideline NG74 concentrates on the referral, assessment and delivery of intermediate care (NICE, 2017). The term ‘intermediate care’ refers to multi-professional services that aim to increase a service user’s independence and support rehabilitation while minimising unnecessary hospitalisation and admissions to long-term care facilities. Intermediate care is comprised of four service models, which include reablement. In most cases, the intervention lasts up to 6 weeks and is delivered by a multidisciplinary team and social care practitioners. Referral to the service can be arranged through either health or social care practitioners. The NICE guidance recommends that reablement is offered as the first line intervention for service users requiring home care, provided that an assessment has established that reablement could help the service user regain independence (NICE, 2017).

As one of the service models, reablement follows the core principles of intermediate care; therefore, the service is goal-driven, focusing on maximising independence and overall wellbeing in addition to taking a person-centred approach to account for varying needs and preferences. The delivery of the service is flexible and individualised to the requirements of the service user. Regular goal reviews are recommended to manage the progression towards set goals and to adjust the duration of the reablement service (NICE, 2017).

NICE (2017) states that there is no evidence on the effectiveness and cost-effectiveness of repeated periods of reablement, or reablement that lasts longer than 6 weeks. Moreover, the data for costs, outcomes and the optimal duration of reablement for different population groups is limited. Therefore, NICE (2017) recommends that further research is conducted to fill the research gaps around reablement.

## 2. RESULTS

### 2.1 Overview of the evidence base

This section summarises the results of the rapid review, which involved reviewing the international literature of reablement interventions. Evidence from studies reporting on clinical effectiveness including person-related outcomes and service-level outcomes is presented in Section 2.2. Evidence from studies reporting on cost-effectiveness is reported in Section 2.3. Section 2.4 provides an overview of other relevant outcomes including costs and implementation outcomes reported in the included studies. This report also includes a summary of the findings of a previously identified qualitative evidence synthesis exploring older people and their carers’ experiences and perceptions of reablement services, which is is provided in Section 3.

A detailed summary of the criteria used for selecting studies for inclusion in the review along with the methods used for conducting the review are presented in Section 6. The rapid review search strategies are presented in Appendix 1. After the removal of duplicates, the database searches identified 4,344 records (see Figure 1 in Section 6.1 for an outline of the study selection process). Following title and abstract screening, 46 papers were retrieved for full text screening. Eighteen studies that evaluated 11 specific rehabilitation interventions were included in this rapid review.

**Figure 1.**
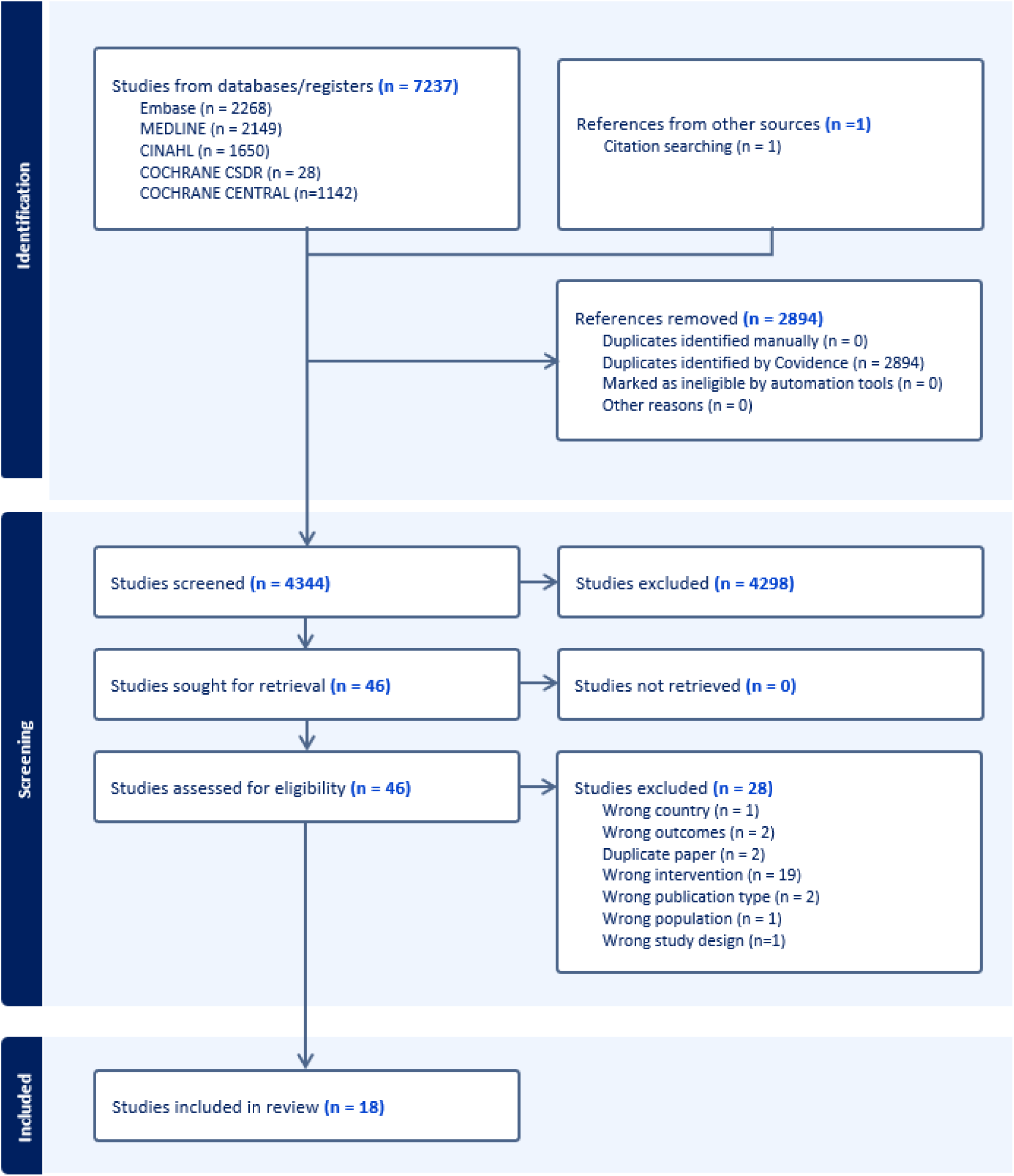
PRISMA 2020 flow diagram of included records (Page et al., 2021)

Of the 18 included studies, 15 reported on the clinical effectiveness of reablement service, two reported on cost-effectiveness, and one reported on both clinical and cost-effectiveness. Hence, the total number of studies reporting on clinical effectiveness was 16. Two studies (Glendinning et al., 2010; Kjerstad and Tuntland, 2016) reporting on cost-effectiveness were trial based economic evaluations. The clinical-effectiveness data from the comparative trial was reported in the same publication for the economic evaluation by Glendinning et al. (2010) and in a sperate publication for the economic evaluation by Kjerstad and Tuntland (2016). This means that Glendinning et al. (2010) is included in both Section 2.2 (on clinical effectiveness) and Section 2.3 (on cost-effectiveness). The clinical-effectiveness data from the RCT used in the economic evaluation reported by Kjerstad and Tuntland (2016) is reported by Tuntland et al., 2015, which is included in Section 2.2. In terms of other relevant outcomes, relating to intervention delivery, reported by included studies, five studies reported on service level costs (Lewin et al., 2014; Lewin et al., 2013b; Beresford et al., 2019; Glendinning et al., 2010; Kjerstad and Tuntland 2016), and three studies reported on implementation outcomes (Beresford et al., 2019; Glendinning et al., 2010; Whitehead et al., 2016).

Our eligibility criteria for reablement interventions were developed with input from stakeholders and based on inclusion of the following common key components (Cochrane et al., 2016; Clotworthy et al., 2021):

- Targeting participants who are aged 65+ or suffering from frailty
- Include a time-limited (typically 6-12 weeks) intensive (multiple visits) programme
- Delivered at the person’s home
- Developed or managed by an allied health professional (such as physical/occupational therapists) and involve a multi-professional team. (But does not have to be delivered by a health professional.)
- Focused on maximising independence, functional ability and social participation
- Person-centred and goal directed

Some interventions included in this review did not strictly comply with all of the criteria listed above. These are highlighted in Table 1. Six studies did not report on the duration of the programme, or the involvement of a multi-professional team. However, stakeholders deemed these studies relevant for inclusion in the review, as they contribute to the development of a comprehensive definition of what could constitute a reablement service for Wales. A summary of the interventions included in the review is also provided in Table 2, which outlines the characteristics of included studies reporting on clinical effectiveness. A more detailed summary of interventions described in the included studies is available in the appendices (section 9.4).

**Table 1:**
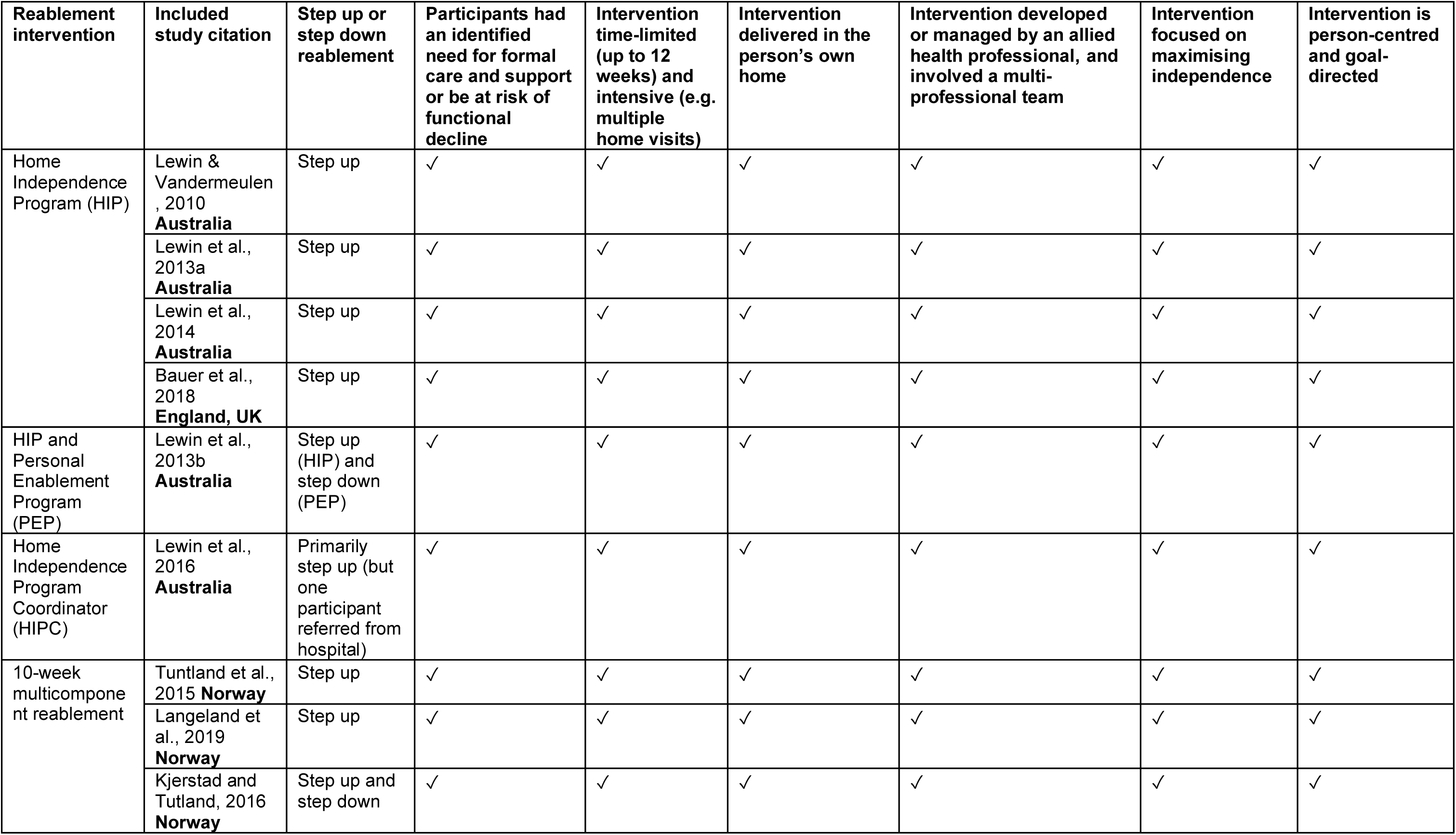
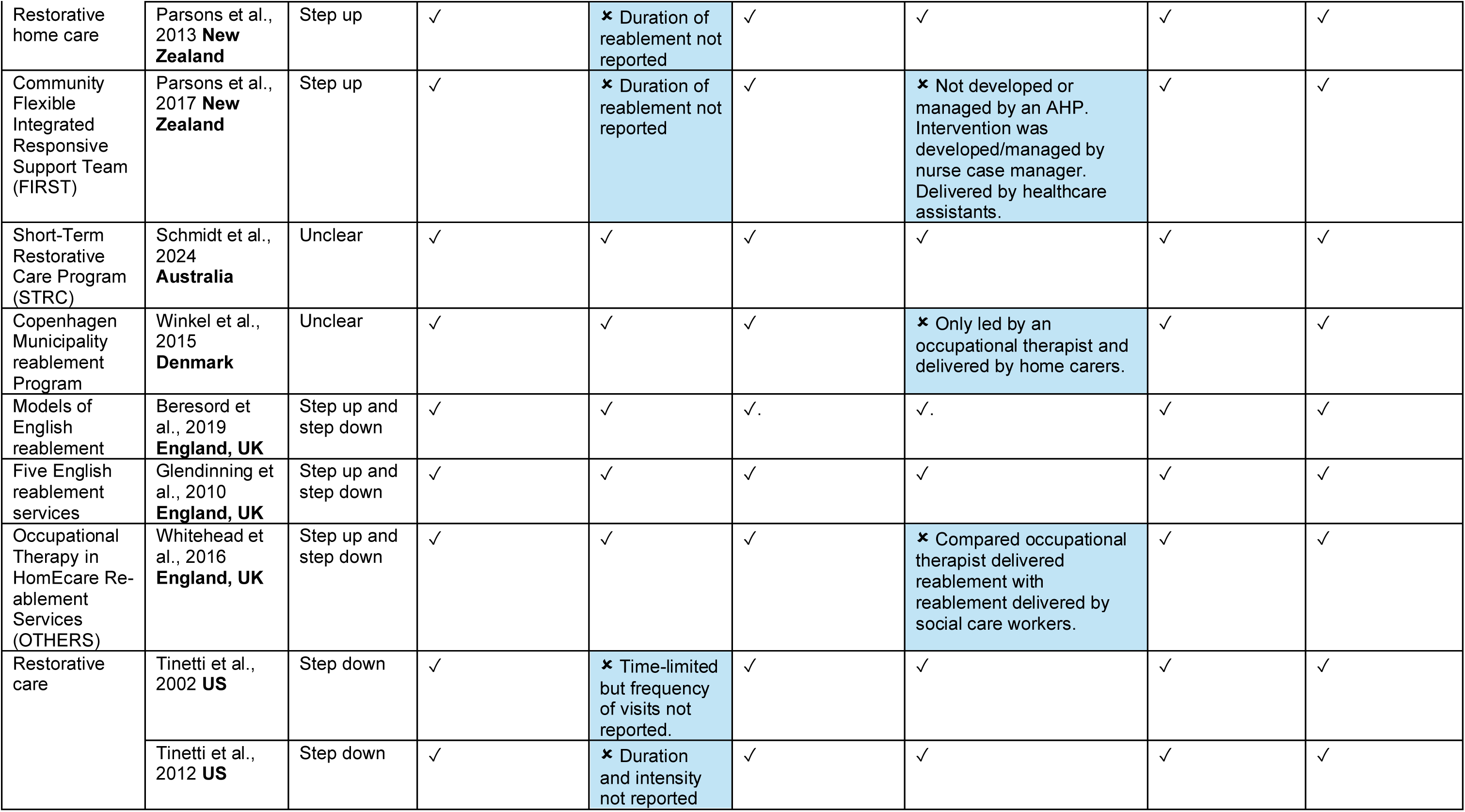
Reablement interventions and studies included in the review and their assessment against key intervention components.

| Reablement intervention | Included study citation | Step up or step down reablement | Participants had an identified need for formal care and support or be at risk of functional decline | Intervention time-limited (up to 12 weeks) and intensive (e.g. multiple home visits) | Intervention delivered in the person's own home | Intervention developed or managed by an allied health professional, and involved a multi-professional team | Intervention focused on maximising independence | Intervention is person-centred and goal-directed |
| --- | --- | --- | --- | --- | --- | --- | --- | --- |
| Home Independence Program (HIP) | Lewin & Vandermeulen, 2010<br>**Australia** | Step up | ✓ | ✓ | ✓ | ✓ | ✓ | ✓ |
|  | Lewin et al., 2013a<br>**Australia** | Step up | ✓ | ✓ | ✓ | ✓ | ✓ | ✓ |
|  | Lewin et al., 2014<br>**Australia** | Step up | ✓ | ✓ | ✓ | ✓ | ✓ | ✓ |
|  | Bauer et al., 2018<br>**England, UK** | Step up | ✓ | ✓ | ✓ | ✓ | ✓ | ✓ |
| HIP and Personal Enablement Program (PEP) | Lewin et al., 2013b<br>**Australia** | Step up (HIP) and step down (PEP) | ✓ | ✓ | ✓ | ✓ | ✓ | ✓ |
| Home Independence Program Coordinator (HIPC) | Lewin et al., 2016<br>**Australia** | Primarily step up (but one participant referred from hospital) | ✓ | ✓ | ✓ | ✓ | ✓ | ✓ |
| 10-week multicomponent reablement | Tuntland et al., 2015<br>**Norway** | Step up | ✓ | ✓ | ✓ | ✓ | ✓ | ✓ |
|  | Langeland et al., 2019<br>**Norway** | Step up | ✓ | ✓ | ✓ | ✓ | ✓ | ✓ |
|  | Kjerstad and Tutland, 2016<br>**Norway** | Step up and step down | ✓ | ✓ | ✓ | ✓ | ✓ | ✓ |
| Restorative home care | Parsons et al., 2013 **New Zealand** | Step up | ✓ | ✗ Duration of reablement not reported | ✓ | ✓ | ✓ | ✓ |
| Community Flexible Integrated Responsive Support Team (FIRST) | Parsons et al., 2017 **New Zealand** | Step up | ✓ | ✗ Duration of reablement not reported | ✓ | ✗ Not developed or managed by an AHP. Intervention was developed/managed by nurse case manager. Delivered by healthcare assistants. | ✓ | ✓ |
| Short-Term Restorative Care Program (STRC) | Schmidt et al., 2024 **Australia** | Unclear | ✓ | ✓ | ✓ | ✓ | ✓ | ✓ |
| Copenhagen Municipality reablement Program | Winkel et al., 2015 **Denmark** | Unclear | ✓ | ✓ | ✓ | ✗ Only led by an occupational therapist and delivered by home carers. | ✓ | ✓ |
| Models of English reablement | Beresford et al., 2019 **England, UK** | Step up and step down | ✓ | ✓ | ✓. | ✓. | ✓ | ✓ |
| Five English reablement services | Glendinning et al., 2010 **England, UK** | Step up and step down | ✓ | ✓ | ✓ | ✓ | ✓ | ✓ |
| Occupational Therapy in HomeCare Reablement Services (OTHERS) | Whitehead et al., 2016 **England, UK** | Step up and step down | ✓ | ✓ | ✓ | ✗ Compared occupational therapist delivered reablement with reablement delivered by social care workers. | ✓ | ✓ |
| Restorative care | Tinetti et al., 2002 **US** | Step down | ✓ | ✗ Time-limited but frequency of visits not reported. | ✓ | ✓ | ✓ | ✓ |
|  | Tinetti et al., 2012 **US** | Step down | ✓ | ✗ Duration and intensity not reported | ✓ | ✓ | ✓ | ✓ |

**Table 2:**
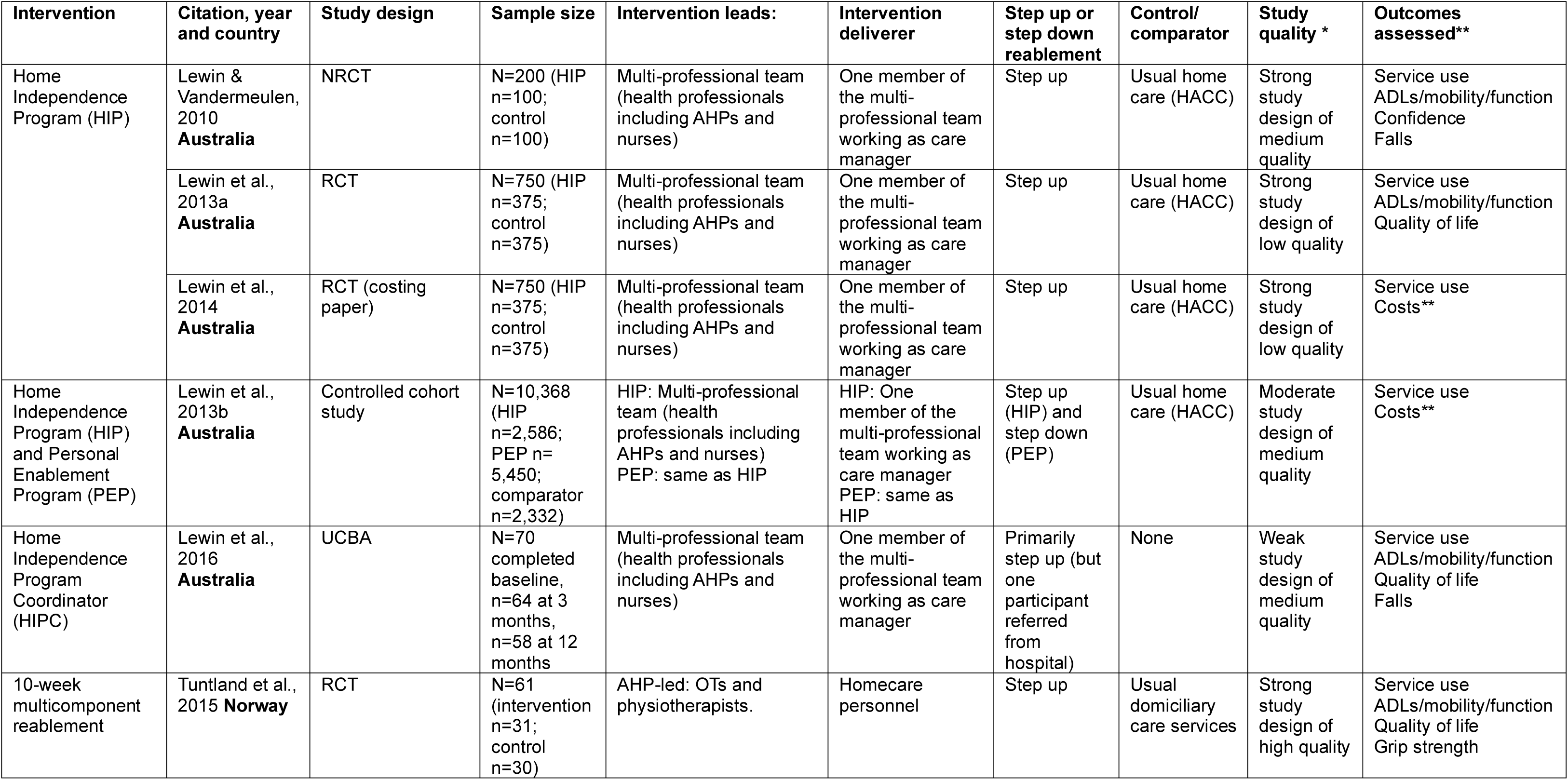
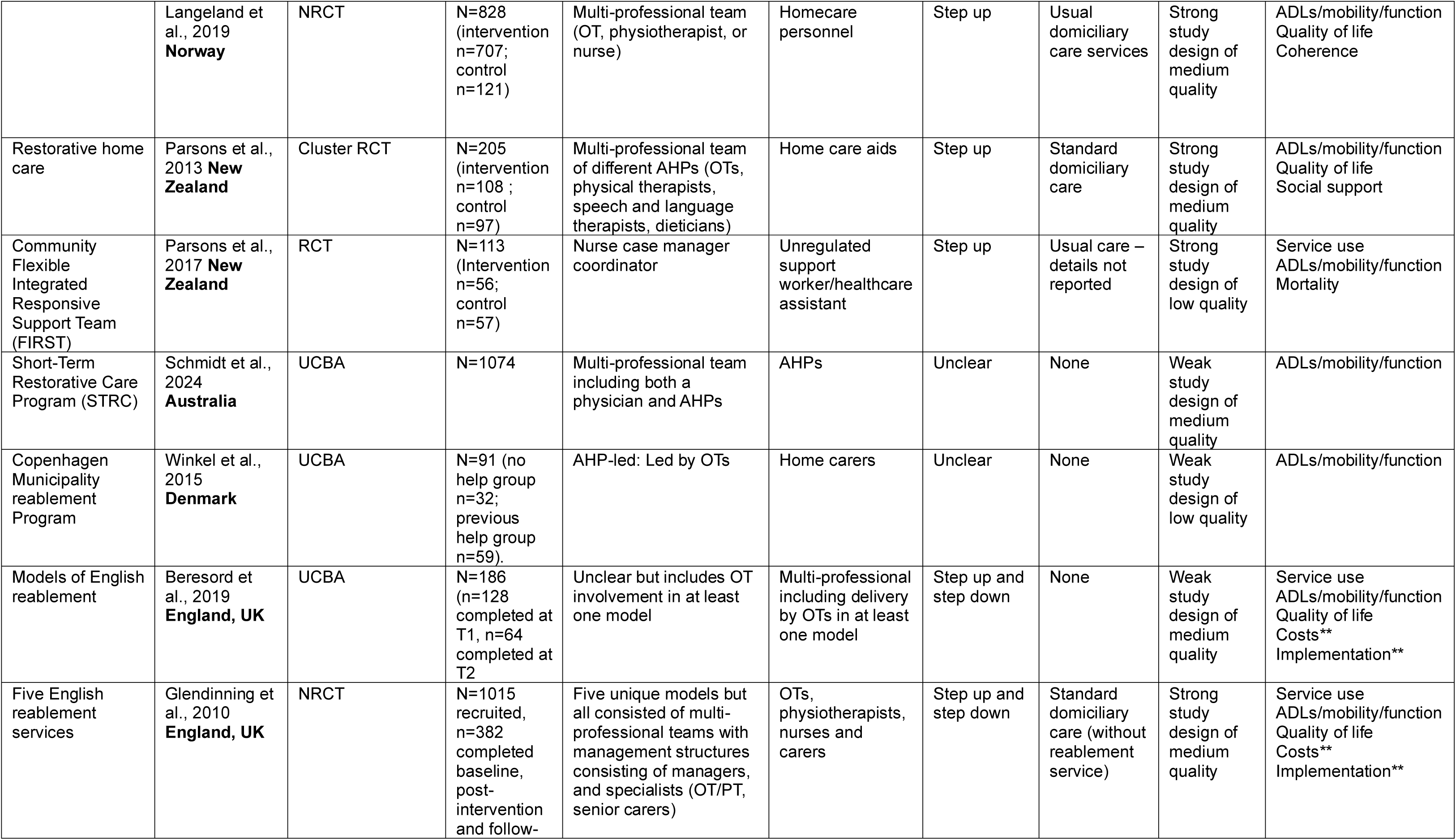
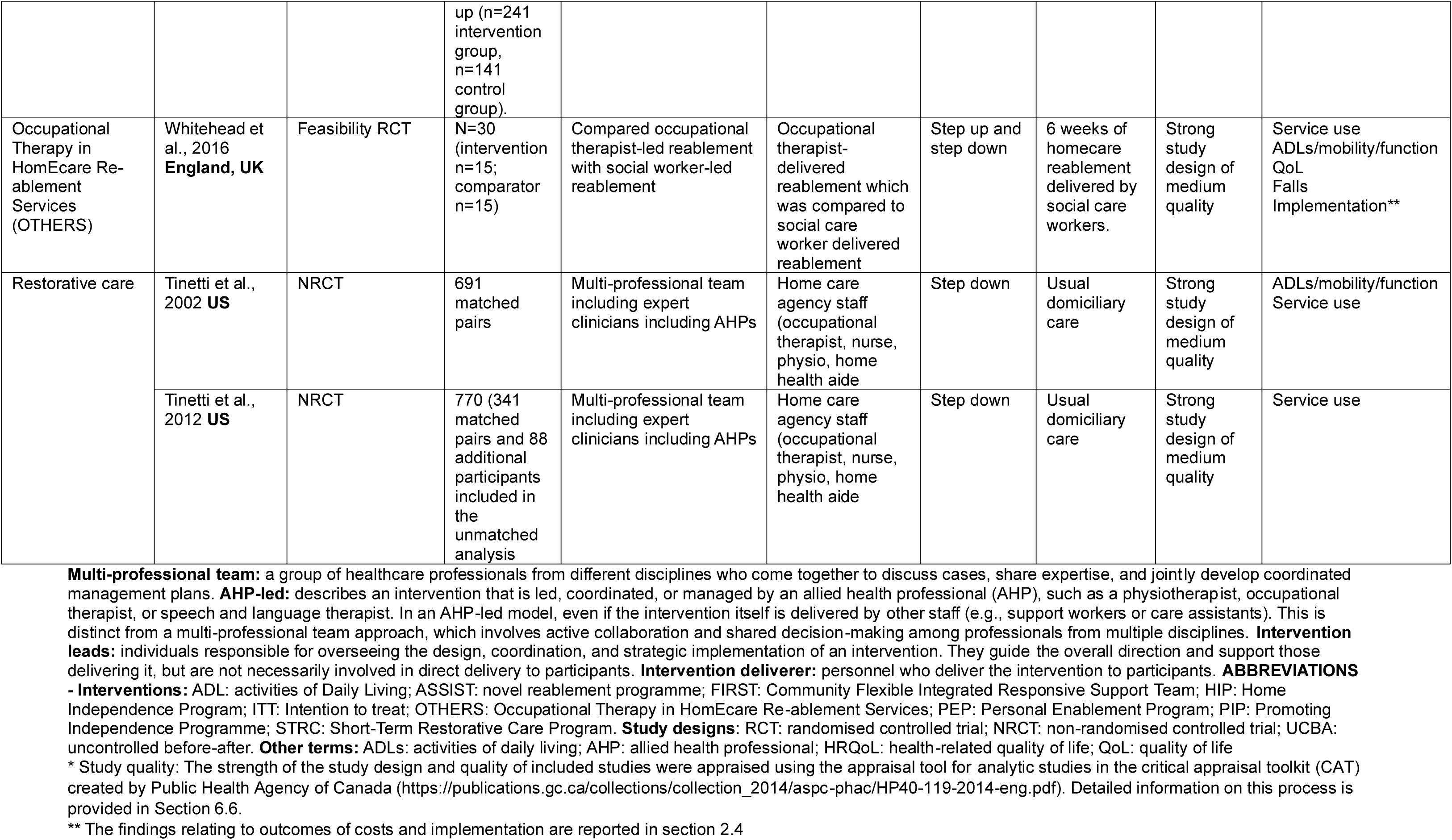
Study characteristics of included primary studies reporting on clinical effectiveness (person-related outcomes and service level outcomes) (n=16).

| Intervention | Citation, year and country | Study design | Sample size | Intervention leads: | Intervention deliverer | Step up or step down reablement | Control/ comparator | Study quality * | Outcomes assessed** |
| --- | --- | --- | --- | --- | --- | --- | --- | --- | --- |
| Home Independence Program (HIP) | Lewin & Vandermeulen, 2010<br>**Australia** | NRCT | N=200 (HIP n=100; control n=100) | Multi-professional team (health professionals including AHPs and nurses) | One member of the multi-professional team working as care manager | Step up | Usual home care (HACC) | Strong study design of medium quality | Service use<br>ADLs/mobility/function<br>Confidence<br>Falls |
|  | Lewin et al., 2013a<br>**Australia** | RCT | N=750 (HIP n=375; control n=375) | Multi-professional team (health professionals including AHPs and nurses) | One member of the multi-professional team working as care manager | Step up | Usual home care (HACC) | Strong study design of low quality | Service use<br>ADLs/mobility/function<br>Quality of life |
|  | Lewin et al., 2014<br>**Australia** | RCT (costing paper) | N=750 (HIP n=375; control n=375) | Multi-professional team (health professionals including AHPs and nurses) | One member of the multi-professional team working as care manager | Step up | Usual home care (HACC) | Strong study design of low quality | Service use<br>Costs** |
| Home Independence Program (HIP) and Personal Enablement Program (PEP) | Lewin et al., 2013b<br>**Australia** | Controlled cohort study | N=10,368 (HIP n=2,586; PEP n=5,450; comparator n=2,332) | HIP: Multi-professional team (health professionals including AHPs and nurses)<br>PEP: same as HIP | HIP: One member of the multi-professional team working as care manager<br>PEP: same as HIP | Step up (HIP) and step down (PEP) | Usual home care (HACC) | Moderate study design of medium quality | Service use<br>Costs** |
| Home Independence Program Coordinator (HIPC) | Lewin et al., 2016<br>**Australia** | UCBA | N=70 completed baseline, n=64 at 3 months, n=58 at 12 months | Multi-professional team (health professionals including AHPs and nurses) | One member of the multi-professional team working as care manager | Primarily step up (but one participant referred from hospital) | None | Weak study design of medium quality | Service use<br>ADLs/mobility/function<br>Quality of life<br>Falls |
| 10-week multicomponent reablement | Tuntland et al., 2015<br>**Norway** | RCT | N=61 (intervention n=31; control n=30) | AHP-led: OTs and physiotherapists. | Homecare personnel | Step up | Usual domiciliary care services | Strong study design of high quality | Service use<br>ADLs/mobility/function<br>Quality of life<br>Grip strength |
|  | Langeland et al., 2019<br>**Norway** | NRCT | N=828 (intervention n=707; control n=121) | Multi-professional team (OT, physiotherapist, or nurse) | Homecare personnel | Step up | Usual domiciliary care services | Strong study design of medium quality | ADLs/mobility/function<br>Quality of life<br>Coherence |
| Restorative home care | Parsons et al., 2013<br>**New Zealand** | Cluster RCT | N=205 (intervention n=108 ; control n=97) | Multi-professional team of different AHPs (OTs, physical therapists, speech and language therapists, dieticians) | Home care aids | Step up | Standard domiciliary care | Strong study design of medium quality | ADLs/mobility/function<br>Quality of life<br>Social support |
| Community Flexible Integrated Responsive Support Team (FIRST) | Parsons et al., 2017<br>**New Zealand** | RCT | N=113 (Intervention n=56; control n=57) | Nurse case manager coordinator | Unregulated support worker/healthcare assistant | Step up | Usual care – details not reported | Strong study design of low quality | Service use<br>ADLs/mobility/function<br>Mortality |
| Short-Term Restorative Care Program (STRC) | Schmidt et al., 2024<br>**Australia** | UCBA | N=1074 | Multi-professional team including both a physician and AHPs | AHPs | Unclear | None | Weak study design of medium quality | ADLs/mobility/function |
| Copenhagen Municipality reablement Program | Winkel et al., 2015<br>**Denmark** | UCBA | N=91 (no help group n=32; previous help group n=59). | AHP-led: Led by OTs | Home carers | Unclear | None | Weak study design of low quality | ADLs/mobility/function |
| Models of English reablement | Beresford et al., 2019<br>**England, UK** | UCBA | N=186 (n=128 completed at T1, n=64 completed at T2) | Unclear but includes OT involvement in at least one model | Multi-professional including delivery by OTs in at least one model | Step up and step down | None | Weak study design of medium quality | Service use<br>ADLs/mobility/function<br>Quality of life<br>Costs**<br>Implementation** |
| Five English reablement services | Glendinning et al., 2010<br>**England, UK** | NRCT | N=1015 recruited, n=382 completed baseline, post-intervention and follow- up (n=241 intervention group, n=141 control group). | Five unique models but all consisted of multi-professional teams with management structures consisting of managers, and specialists (OT/PT, senior carers) | OTs, physiotherapists, nurses and carers | Step up and step down | Standard domiciliary care (without reablement service) | Strong study design of medium quality | Service use<br>ADLs/mobility/function<br>Quality of life<br>Costs**<br>Implementation** |
| Occupational Therapy in HomeCare Re-ablement Services (OTHERS) | Whitehead et al., 2016<br>**England, UK** | Feasibility RCT | N=30 (intervention n=15; comparator n=15) | Compared occupational therapist-led reablement with social worker-led reablement | Occupational therapist-delivered reablement which was compared to social care worker delivered reablement | Step up and step down | 6 weeks of homecare reablement delivered by social care workers. | Strong study design of medium quality | Service use<br>ADLs/mobility/function<br>QoL<br>Falls<br>Implementation** |
| Restorative care | Tinetti et al., 2002 **US** | NRCT | 691 matched pairs | Multi-professional team including expert clinicians including AHPs | Home care agency staff (occupational therapist, nurse, physio, home health aide) | Step down | Usual domiciliary care | Strong study design of medium quality | ADLs/mobility/function<br>Service use |
|  | Tinetti et al., 2012 **US** | NRCT | 770 (341 matched pairs and 88 additional participants included in the unmatched analysis) | Multi-professional team including expert clinicians including AHPs | Home care agency staff (occupational therapist, nurse, physio, home health aide) | Step down | Usual domiciliary care | Strong study design of medium quality | Service use |

### 2.2 Clinical effectiveness of reablement interventions

#### 2.2.1 Overview of the evidence on effectiveness of reablement interventions

A summary of the characteristics of included studies reporting on the clinical effectiveness of reablement interventions is presented in Table 2. Of the 16 studies that reported on clinical effectiveness, six were randomised controlled trials (RCTs), five were non-randomised controlled trials, one was a (controlled) cohort study, and four were uncontrolled before and after studies. Eight studies evaluated a step-up reablement model to support individuals at risk of deteriorating and help prevent hospital admissions or the need to move to a care home. Two studies focused on the step-down reablement model to support someone’s recovery after hospital discharge and help prevent readmission. Four studies evaluated a reablement model that covered both step-up and step-down, whilst the focus was unclear for the remaining two studies. Four studies evaluated a therapist-led reablement model. Two studies evaluated a reablement intervention led by an occupational therapist or physiotherapist (Tuntland et al., 2015; Langeland et al., 2019) (nurse-led was also an option in one of these studies; Langeland et al., 2019), and one was explicitly occupational therapist-led (Winkel et al., 2015). The fourth study compared occupational therapist-led reablement with social care worker-led reablement (Whitehead et al., 2016). Six studies evaluated a reablement model led by a multi-professional teams that included allied health professionals (Tinetti et al., 2002; Tinetti et al., 2012; Parsons et al., 2013; Schmidt et al., 2024; Glendinning et al., 2010; Beresford et al., 2019). Five studies evaluated a reablement model that was led by muti-professional teams that included allied health professionals and nurses. These five studies evaluated the Home Independence Program (HIP) or an adaptation of this (Lewin & Vandermeulen, 2010; Lewin et al., 2013a; Lewin et al., 2014; Lewin et al., 2013b; Lewin et al., 2016). One study evaluated a model led by a nurse case manager coordinator (Parsons et al., 2017). Across the 16 studies, the reablement intervention was delivered by a range of personnel, including: an occupational therapist (1 study); a member of a multi-professional team or multi-professional workforce (9 studies); a multi-professional team consisting of range of different AHPs only (1 study) and non-health professionals such as homecare personnel, healthcare assistants, home care aides, or carers (5 studies).

Six were conducted in Australia (Lewin & Vandermeulen, 2010; Lewin et al., 2013a; Lewin et al., 2013b; Lewin et al., 2014, Lewin et al., 2016; Schmidt et al., 2024), three in the UK (Beresford et al., 2019; Glendinning et al., 2010; Whitehead et al., 2016), two in New Zealand (Parsons et al., 2013; Parsons et al., 2017), two in Norway (Tuntland et al., 2015; Langeland et al., 2019), two in the US (Tinetti et al., 2002; Tinetti et al., 2012), and one was conducted in Denmark (Winkel et al., 2015).

Thirteen studies assessed the effectiveness of reablement interventions on person-related outcomes including mobility and activities of daily living (ADL) (Lewin & Vandermeulen, 2010; Lewin et al., 2013a; Lewin et al., 2016; Tuntland et al., 2015; Langeland et al., 2019; Parsons et al., 2013; Parsons et al., 2017; Schmidt et al., 2024; Winkel et al., 2015; Beresford et al., 2019; Whitehead et al., 2016; Glendinning et al., 2010; Tinetti et al., 2002), quality of life (Lewin et al., 2013a; Lewin et al., 2016; Tuntland et al., 2015; Langeland et al., 2019; Parsons et al., 2013; Beresford et al., 2019; Whitehead et al., 2016; Glendinning et al., 2010), falls outcomes (Lewin & Vandermeulen, 2010; Lewin et al., 2016; Whitehead et al., 2016), grip strength (Tuntland et al., 2015), coherence (Langeland et al., 2019), social support (Parsons et al., 2013), and mortality (Parsons et al., 2017).

Twelve studies assessed the effectiveness of reablement on service-level outcomes including the use of home care services (Lewin & Vandermeulen, 2010; Lewin et al., 2013a; Lewin et al., 2014; Lewin et al., 2013b; Lewin et al., 2016; Beresford et al., 2019; Glendinning et al., 2010; Tinetti et al., 2012), hospital admissions (Lewin et al., 2013a; Lewin et al., 2014; Lewin et al., 2016; Parsons et al., 2017; Beresford et al., 2019; Whitehead et al., 2016; Glendinning et al., 2010; Tinetti et al., 2012), residential care admissions (Lewin et al., 2013a; Lewin et al., 2016; Parsons et al., 2017), and other health and social care service use outcomes (Lewin et al., 2014; Tuntland et al., 2015; Parsons et al., 2017; Beresford et al., 2019; Whitehead et al., 2016; Glendinning et al., 2010; Tinetti et al., 2002).

#### 2.2.2 Quality of studies evaluating the effectiveness of reablement on person-related and service-level outcomes

The studies that evaluated the effectiveness of reablement on person-related and service-level outcomes were critically appraised using the Analytic Study Critical Appraisal Tool, which is part of the Critical Appraisal Toolkit (CAT) for assessing multiple types of evidence (Public Health Agency of Canada 2014). CAT provides a toolkit for evaluating the evidence base, which incorporates the assessment of individual studies and the overall body of evidence to inform practice and policy development (see section 6.6 for further details). A summary rating of the strength of the study design and quality of individual studies is provided in Table 2. Following the results section and results summary tables for person-related outcomes (Table 3) and service level outcomes (Table 4), a summary of the rating of the certainty of the overall evidence for the effectiveness of reablement on person-related outcomes and service-level outcomes is presented at the end of Section 2.2. in Table 5.

**Table 3:**
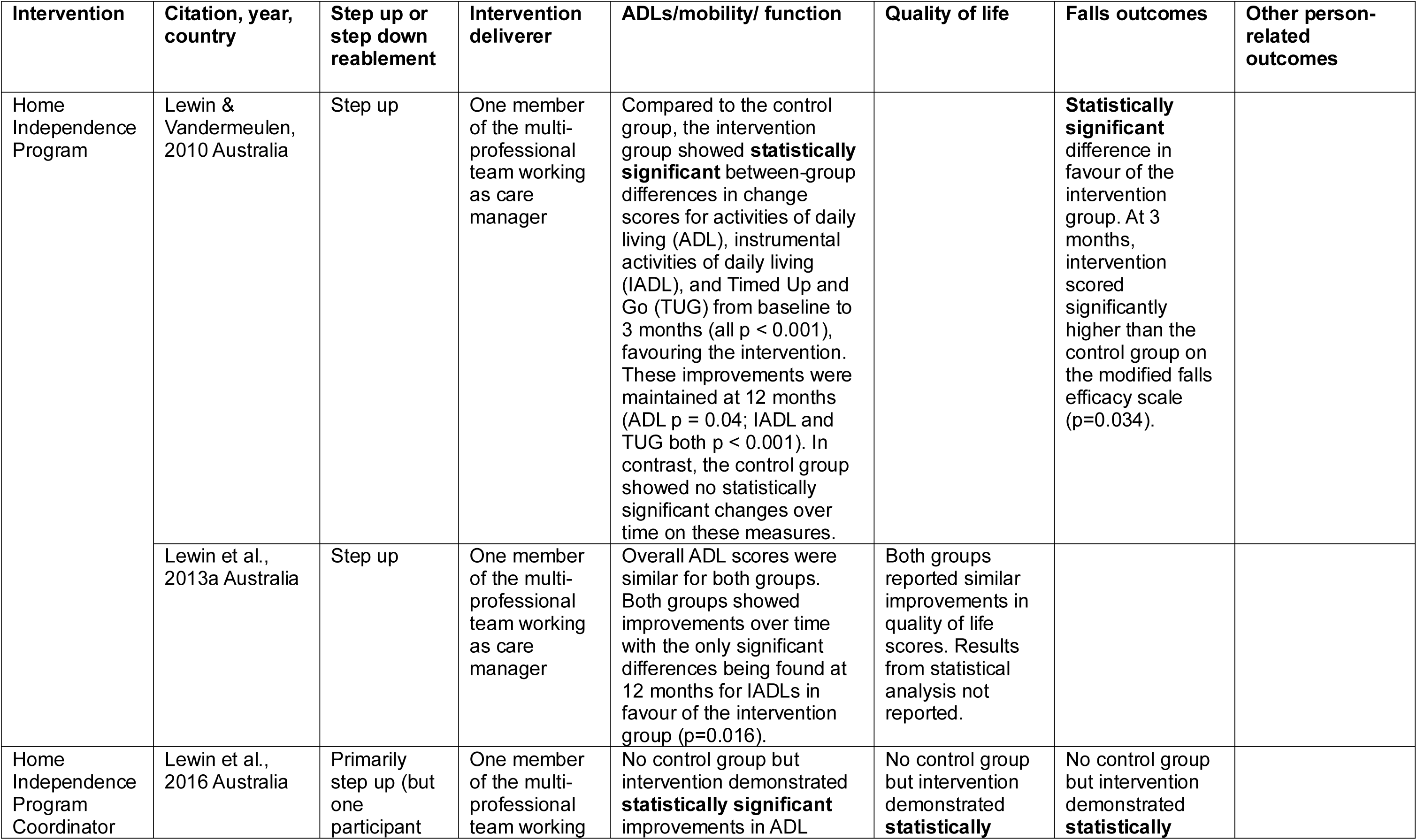
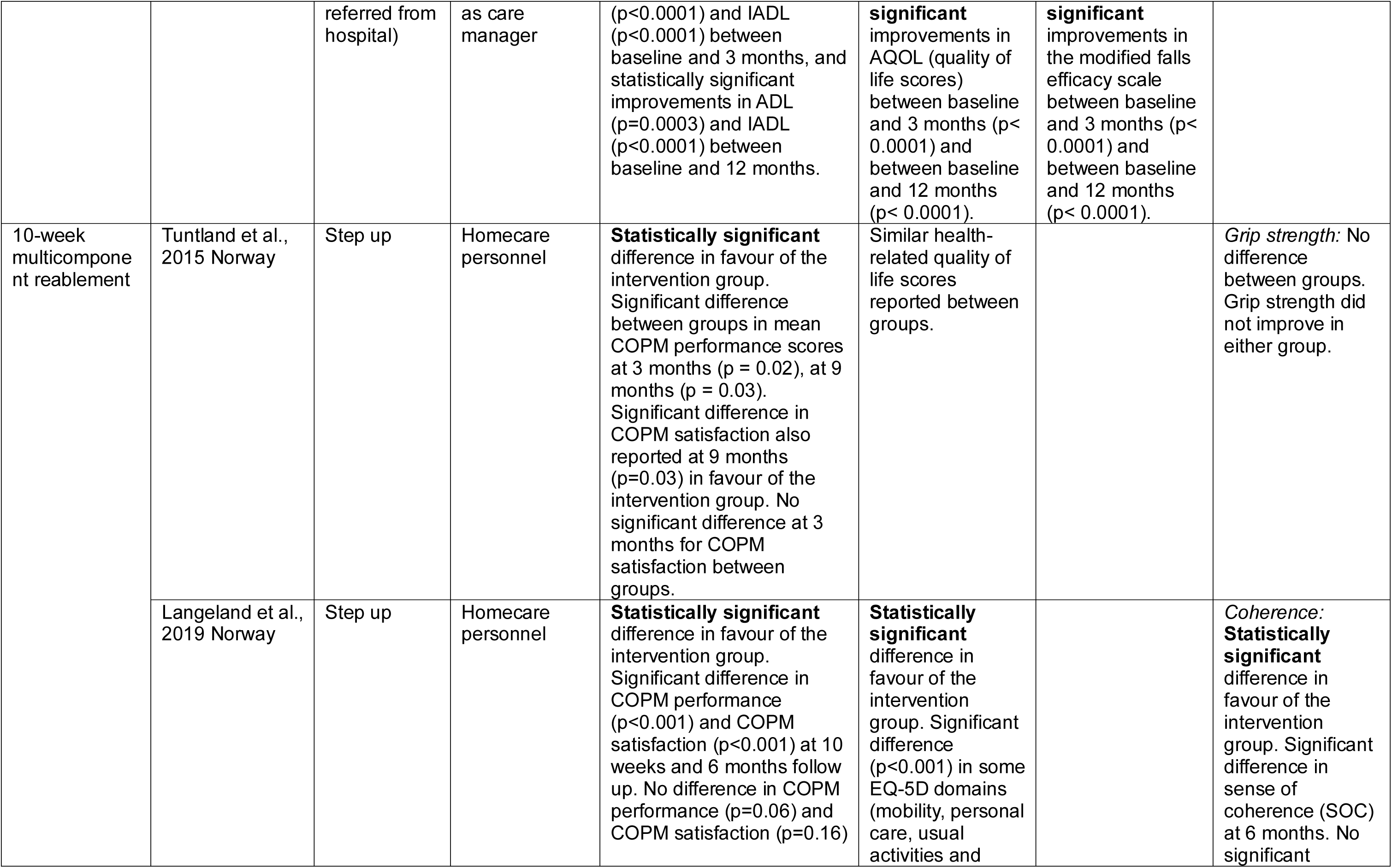
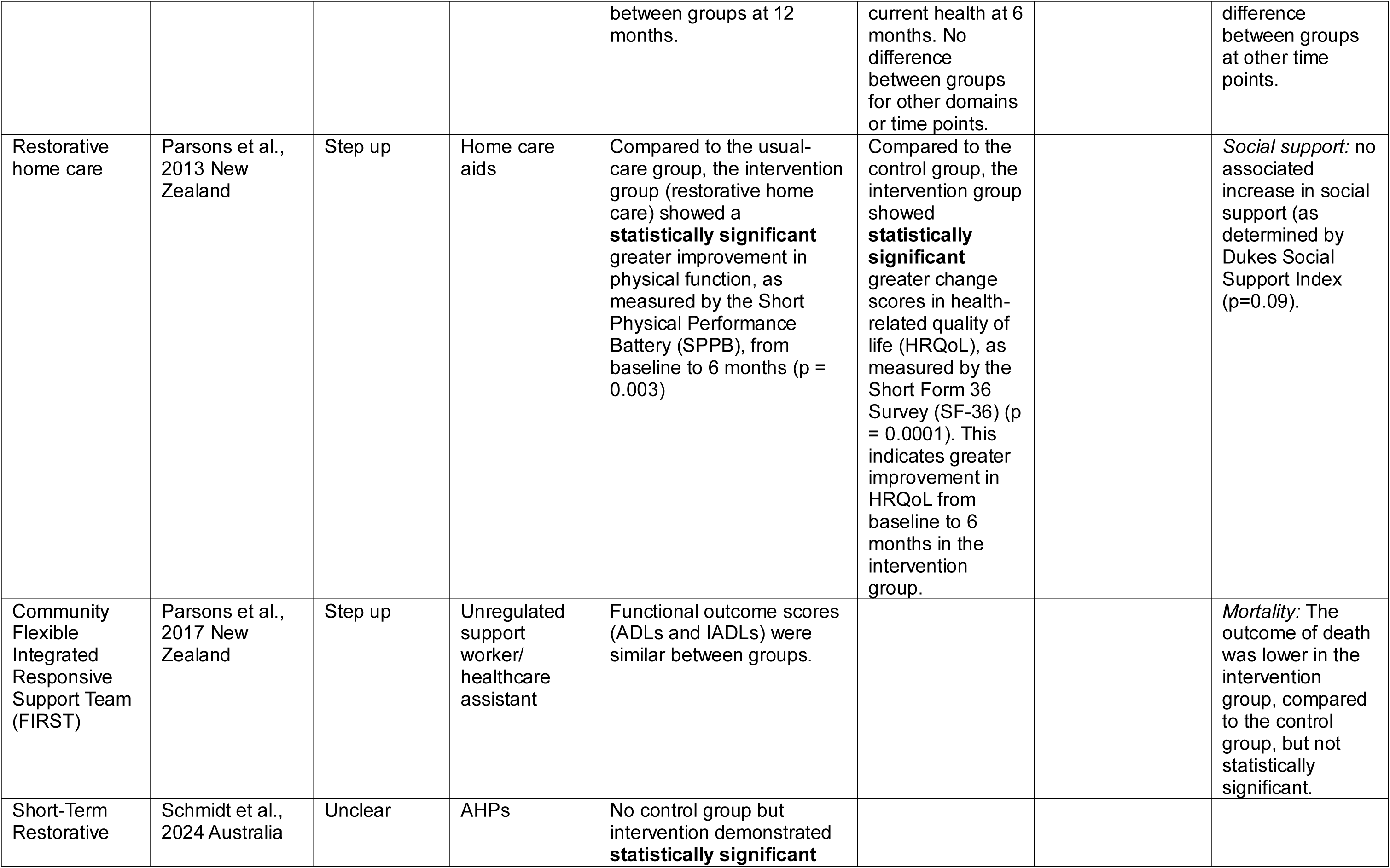
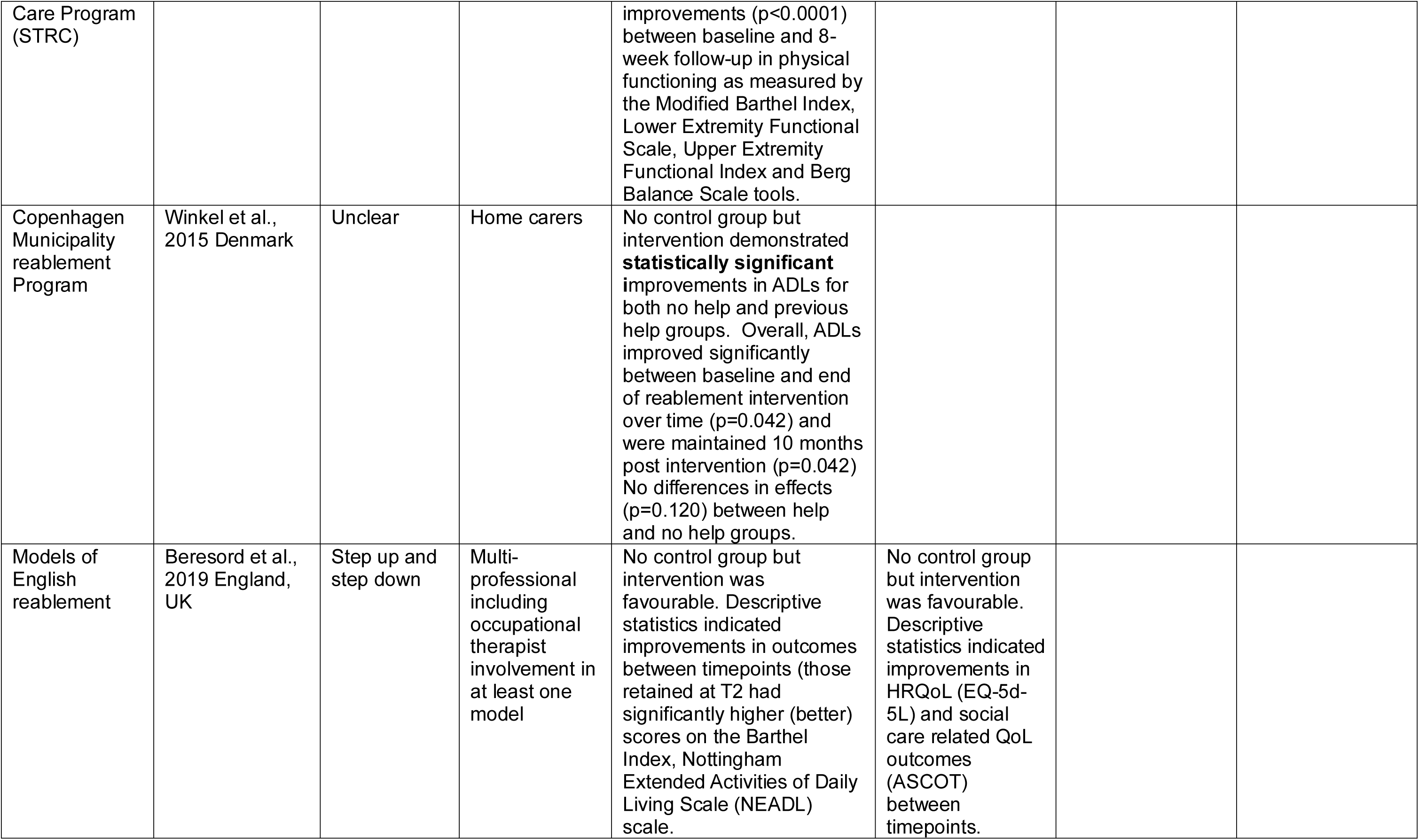
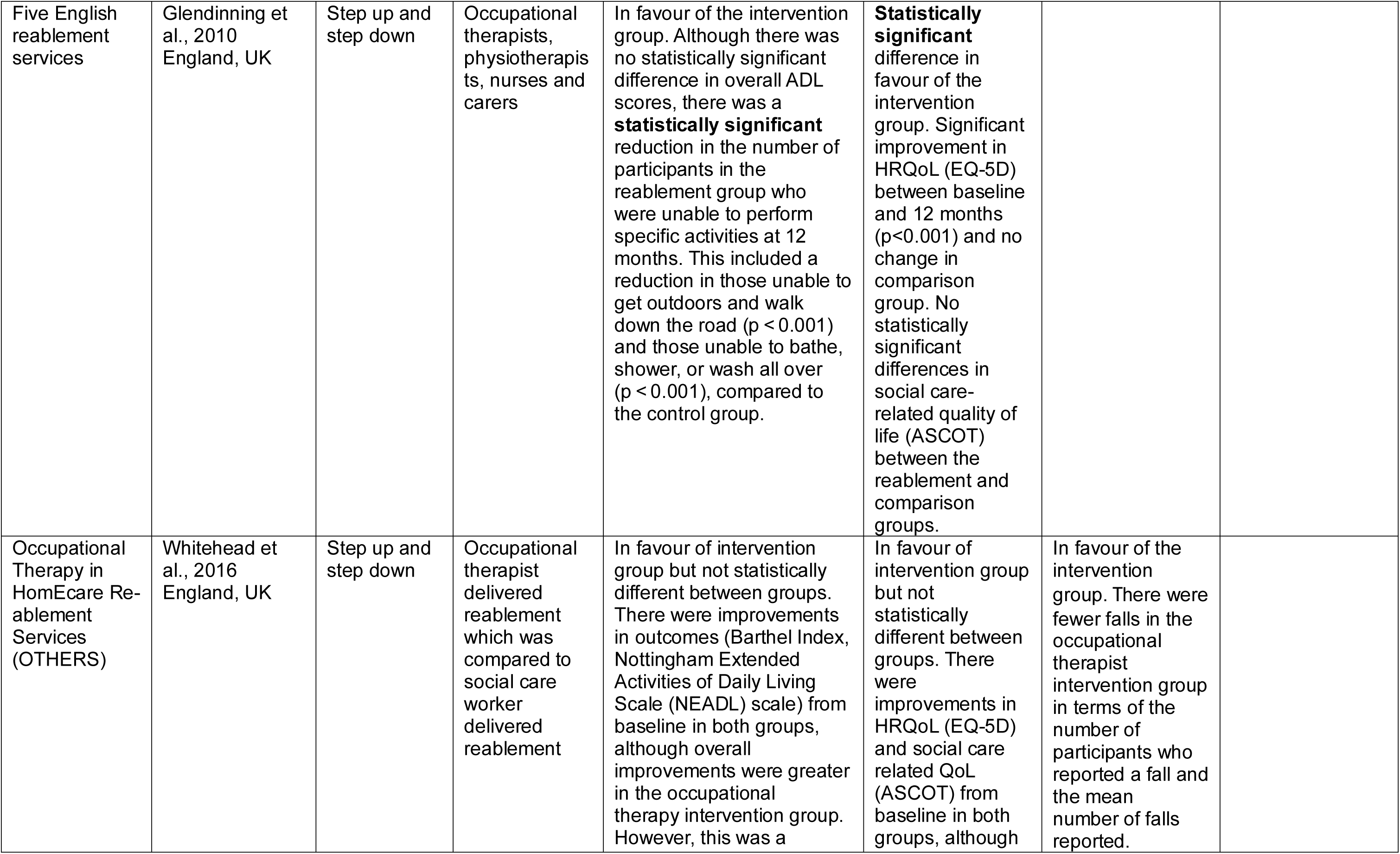
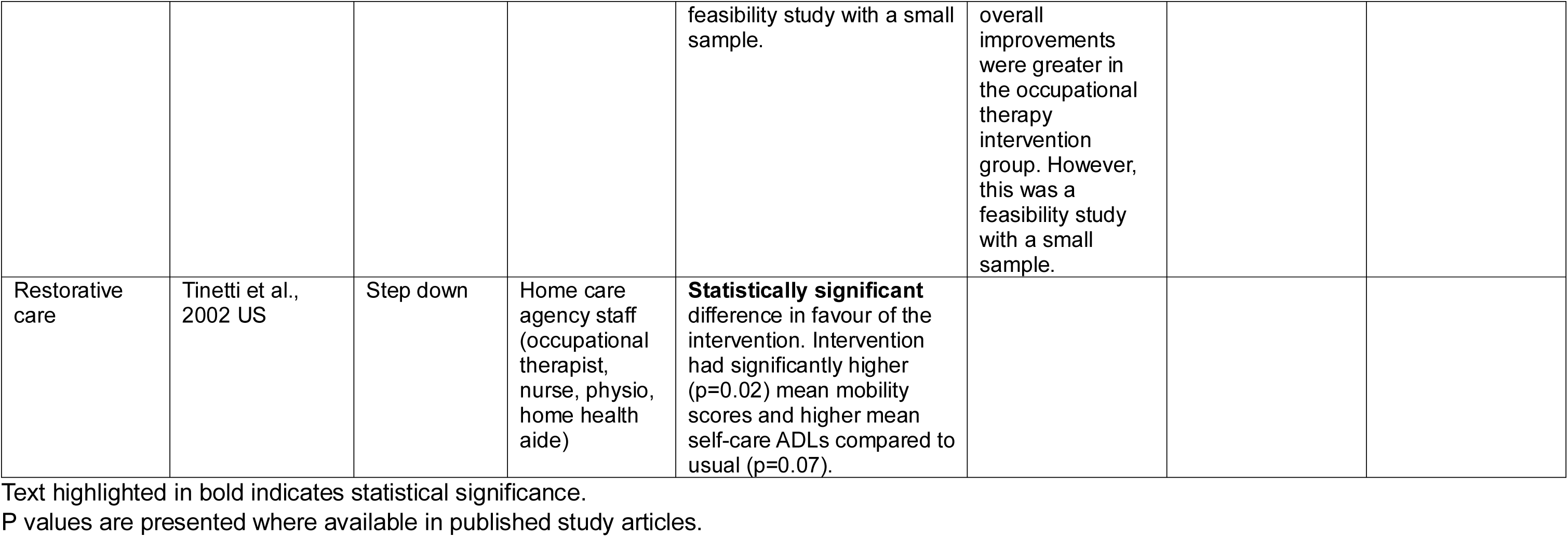
Summary of main findings for person-related outcomes.

| Intervention | Citation, year, country | Step up or step down reablement | Intervention deliverer | ADLs/mobility/ function | Quality of life | Falls outcomes | Other person-related outcomes |
| --- | --- | --- | --- | --- | --- | --- | --- |
| Home Independence Program | Lewin & Vandermeulen, 2010 Australia | Step up | One member of the multi-professional team working as care manager | Compared to the control group, the intervention group showed **statistically significant** between-group differences in change scores for activities of daily living (ADL), instrumental activities of daily living (IADL), and Timed Up and Go (TUG) from baseline to 3 months (all $p < 0.001$), favouring the intervention. These improvements were maintained at 12 months (ADL $p = 0.04$; IADL and TUG both $p < 0.001$). In contrast, the control group showed no statistically significant changes over time on these measures. | | **Statistically significant** difference in favour of the intervention group. At 3 months, intervention scored significantly higher than the control group on the modified falls efficacy scale ($p=0.034$). | |
| | Lewin et al., 2013a Australia | Step up | One member of the multi-professional team working as care manager | Overall ADL scores were similar for both groups. Both groups showed improvements over time with the only significant differences being found at 12 months for IADLs in favour of the intervention group ($p=0.016$). | Both groups reported similar improvements in quality of life scores. Results from statistical analysis not reported. | | |
| Home Independence Program Coordinator | Lewin et al., 2016 Australia | Primarily step up (but one participant | One member of the multi-professional team working | No control group but intervention demonstrated **statistically significant** improvements in ADL | No control group but intervention demonstrated **statistically** | No control group but intervention demonstrated **statistically** |  |
| | | referred from hospital) | as care manager | ($p<0.0001$) and IADL ($p<0.0001$) between baseline and 3 months, and statistically significant improvements in ADL ($p=0.0003$) and IADL ($p<0.0001$) between baseline and 12 months. | **significant** improvements in AQOL (quality of life scores) between baseline and 3 months ($p<0.0001$) and between baseline and 12 months ($p<0.0001$). | **significant** improvements in the modified falls efficacy scale between baseline and 3 months ($p<0.0001$) and between baseline and 12 months ($p<0.0001$). | |
| 10-week multicomponent reablement | Tuntland et al., 2015 Norway | Step up | Homecare personnel | **Statistically significant** difference in favour of the intervention group. Significant difference between groups in mean COPM performance scores at 3 months ($p = 0.02$), at 9 months ($p = 0.03$). Significant difference in COPM satisfaction also reported at 9 months ($p=0.03$) in favour of the intervention group. No significant difference at 3 months for COPM satisfaction between groups. | Similar health-related quality of life scores reported between groups. | | *Grip strength:* No difference between groups. Grip strength did not improve in either group. |
| | Langeland et al., 2019 Norway | Step up | Homecare personnel | **Statistically significant** difference in favour of the intervention group. Significant difference in COPM performance ($p<0.001$) and COPM satisfaction ($p<0.001$) at 10 weeks and 6 months follow up. No difference in COPM performance ($p=0.06$) and COPM satisfaction ($p=0.16$) | **Statistically significant** difference in favour of the intervention group. Significant difference ($p<0.001$) in some EQ-5D domains (mobility, personal care, usual activities and | | *Coherence:* **Statistically significant** difference in favour of the intervention group. Significant difference in sense of coherence (SOC) at 6 months. No significant |
|  |  |  |  | between groups at 12 months. | current health at 6 months. No difference between groups for other domains or time points. |  | difference between groups at other time points. |
| Restorative home care | Parsons et al., 2013 New Zealand | Step up | Home care aids | Compared to the usual-care group, the intervention group (restorative home care) showed a **statistically significant** greater improvement in physical function, as measured by the Short Physical Performance Battery (SPPB), from baseline to 6 months ($p = 0.003$) | Compared to the control group, the intervention group showed **statistically significant** greater change scores in health-related quality of life (HRQoL), as measured by the Short Form 36 Survey (SF-36) ($p = 0.0001$). This indicates greater improvement in HRQoL from baseline to 6 months in the intervention group. | | *Social support:* no associated increase in social support (as determined by Dukes Social Support Index ($p=0.09$)). |
| Community Flexible Integrated Responsive Support Team (FIRST) | Parsons et al., 2017 New Zealand | Step up | Unregulated support worker/ healthcare assistant | Functional outcome scores (ADLs and IADLs) were similar between groups. |  |  | *Mortality:* The outcome of death was lower in the intervention group, compared to the control group, but not statistically significant. |
| Short-Term Restorative | Schmidt et al., 2024 Australia | Unclear | AHPs | No control group but intervention demonstrated **statistically significant** |  |  |  |
| Care Program (STRC) | | | | improvements ($p<0.0001$) between baseline and 8-week follow-up in physical functioning as measured by the Modified Barthel Index, Lower Extremity Functional Scale, Upper Extremity Functional Index and Berg Balance Scale tools. | | | |
| Copenhagen Municipality reablement Program | Winkel et al., 2015 Denmark | Unclear | Home carers | No control group but intervention demonstrated **statistically significant** improvements in ADLs for both no help and previous help groups. Overall, ADLs improved significantly between baseline and end of reablement intervention over time ($p=0.042$) and were maintained 10 months post intervention ($p=0.042$) No differences in effects ($p=0.120$) between help and no help groups. | | | |
| Models of English reablement | Beresford et al., 2019 England, UK | Step up and step down | Multi-professional including occupational therapist involvement in at least one model | No control group but intervention was favourable. Descriptive statistics indicated improvements in outcomes between timepoints (those retained at T2 had significantly higher (better) scores on the Barthel Index, Nottingham Extended Activities of Daily Living Scale (NEADL) scale. | No control group but intervention was favourable. Descriptive statistics indicated improvements in HRQoL (EQ-5d-5L) and social care related QoL outcomes (ASCOT) between timepoints. |  |  |

|  |  |  |  |  |  |  |
| --- | --- | --- | --- | --- | --- | --- |
| Five English reablement services | Glendinning et al., 2010<br>England, UK | Step up and step down | Occupational therapists, physiotherapists, nurses and carers | In favour of the intervention group. Although there was no statistically significant difference in overall ADL scores, there was a **statistically significant** reduction in the number of participants in the reablement group who were unable to perform specific activities at 12 months. This included a reduction in those unable to get outdoors and walk down the road ($p < 0.001$) and those unable to bathe, shower, or wash all over ($p < 0.001$), compared to the control group. | **Statistically significant** difference in favour of the intervention group. Significant improvement in HRQoL (EQ-5D) between baseline and 12 months ($p < 0.001$) and no change in comparison group. No statistically significant differences in social care-related quality of life (ASCOT) between the reablement and comparison groups. | |
| Occupational Therapy in HomeCare Reablement Services (OTHERS) | Whitehead et al., 2016<br>England, UK | Step up and step down | Occupational therapist delivered reablement which was compared to social care worker delivered reablement | In favour of intervention group but not statistically different between groups. There were improvements in outcomes (Barthel Index, Nottingham Extended Activities of Daily Living Scale (NEADL) scale) from baseline in both groups, although overall improvements were greater in the occupational therapy intervention group. However, this was a | In favour of intervention group but not statistically different between groups. There were improvements in HRQoL (EQ-5D) and social care related QoL (ASCOT) from baseline in both groups, although | In favour of the intervention group. There were fewer falls in the occupational therapist intervention group in terms of the number of participants who reported a fall and the mean number of falls reported. |

|  |  |  |  |  |  |
| --- | --- | --- | --- | --- | --- |
|  |  |  |  | feasibility study with a small sample. | overall improvements were greater in the occupational therapy intervention group. However, this was a feasibility study with a small sample. |
| Restorative care | Tinetti et al., 2002 US | Step down | Home care agency staff (occupational therapist, nurse, physio, home health aide) | **Statistically significant** difference in favour of the intervention. Intervention had significantly higher (p=0.02) mean mobility scores and higher mean self-care ADLs compared to usual (p=0.07). |  |

Text highlighted in bold indicates statistical significance.
P values are presented where available in published study articles.

**Table 4:** Summary of main findings for service level outcomes.

| Intervention name | Citation, year, country | Step up or step down reablement | Intervention deliverer | Hospital admissions | Home care service use | Residential care | Other health and social care service use |
| --- | --- | --- | --- | --- | --- | --- | --- |
| Home Independence Program (HIP) | Lewin & Vandermeulen, 2010 Australia | Step up | One member of the multi-professional team working as care manager | | **Statistically significant**<br>difference in favour of the intervention. Individuals who received HIP had significantly lower odds of requiring services, 0.07 times at 3 months and 0.14 times at 12 months, compared to the control group ($p < 0.001$ for both). | | |
| | Lewin et al., 2013a Australia | Step up | One member of the multi-professional team working as care manager | In favour of the control group, but not statistically significant. At 3 months, there were 6 and 24 hospital admission in the control and intervention groups, respectively. At 12 months however, people receiving usual domiciliary care were more likely to be admitted to hospital, with 3 hospital admissions recorded in that group and 1 | **Statistically significant**<br>difference in favour of the intervention. HIP was found to substantially and significantly decrease the odds of having an ongoing service at both 3 and 12 months ($p < 0.001$ for both). | In favour of the intervention group, but not statistically significant. At 3 months, there were 21 and 16 admissions to residential care in the control and intervention groups, respectively. At 12 months, there were 48 and 44 admissions in the control and intervention groups, respectively. | |
|  |  |  |  | recorded in the HIP group. |  |  |  |
| | Lewin et al., 2014 Australia | Step up | One member of the multi-professional team working as care manager | In favour of the intervention group, but not statistically significant. There were fewer hospitalisations in the intervention group compared with the control group during the first and second year, and overall at 24 months. | **Statistically significant** difference in favour of the intervention. The intervention used significantly less home-care hours compared to the control group ($p < 0.001$) during the first and second year, and overall after 24 months. | | *Emergency department visits:* In favour of the intervention group, but not statistically significant. There were fewer emergency department presentations in the intervention group compared with the control group during the first and second year, and overall at 24 months. |
| Home Independence Program (HIP) and Personal Enablement Program (PEP) | Lewin et al., 2013b Australia | Step up (HIP) and step down (PEP) | HIP: One member of the multi-professional team working as care manager<br>PEP: same as HIP |  | In favour of the intervention group, but not statistically significant. Compared to standard home care, both HIP and PEP groups used fewer home care services in the first 3 years, with HIP remaining lower in year 3. Both HIP and PEP used less services over 57 months, with HIP still lower in the final year. |  |  |
| Home Independence | Lewin et al., 2016 Australia | Primarily step up (but | One member of the multi- | In favour of the HIPC intervention, | In favour of the intervention but not | In favour of the intervention, but |  |
| Program Coordinator (HIPC) |  | one participant referred from hospital) | professional team working as care manager | but not statistically significant. Secondary comparison reported slightly less hospitalisations than previous RCT (Lewin et al., 2013a). | statistically significant. Secondary analysis found that both groups reported a similar reduced need for ongoing home care services at 3 and 12 months. Outcomes for the HIPC group at 3 months and 12 months were somewhat better than the HIP group with more participants no longer requiring home care services. | not statistically significant. Secondary comparison reported slightly less transfers to residential care than previous RCT (Lewin et al., 2013a). |  |
| 10-week multicomponent reablement | Tuntland et al., 2015 Norway | Step up | Homecare personnel |  |  |  | *Outpatient treatments:*<br>**Statistically significant** in favour of the intervention group. Statistically significant difference (p=0.007) in outpatient treatments between the groups at 3-month follow-up: control group (12 treatments) vs intervention (3 treatments). |
| Community Flexible Integrated Responsive Support Team (FIRST) | Parsons et al., 2017 New Zealand | Step up | Unregulated support worker/ healthcare assistant | Both groups reported similar hospital admissions over 12 months. No data is shown. |  | In favour of the intervention group but not statistically significant. Permanent institutional care was lower in the intervention group compared to the control group, but not statistically significant. | *Other resource use including emergency department visits:* In favour of the control but not statistically significant. Over 12 months, the intervention used more day care, day centre, meals on wheels and transport and slightly more GP visits. Emergency department presentations were similar for both groups. No data is shown to report frequencies or means. |
| Models of English reablement | Beresford et al., 2019 England, UK | Step up and step down | Multi-professional including occupational therapist involvement in at least one model | No comparator but intervention produced positive outcomes but not statistically significant. Lower number of hospital admissions between baseline and time point 1 and 2. | No comparator but intervention produced positive outcomes but not statistically significant differences. Lower number of home care services between baseline and time point 1 and 2. |  | *Community and social care resource use:* No comparator but intervention produced positive outcomes but not statistically significant. Compared to baseline, there was reduced use of community care and social care services at time point 1 and 2. |
| Five English reablement services | Glendinning et al., 2010<br>England, UK | Step up and step down | Occupational therapists, physiotherapists, nurses and carers | In favour of the comparator group but not statistically significant. At the 10-month follow-up, the intervention had higher hospital inpatient stays (n=237) compared with the comparison group (n=139) | In favour of the intervention group but not statistically significant. The percentage of people receiving independent home care was lower in the reablement than the comparison group both within the first eight weeks (2% and 65% respectively) and the remaining ten months (18% and 50% respectively). |  | *Social care resource use:* In favour of the intervention but not statistically significant. Only six people in the comparison group and none in the reablement group used day care services in the 10-month follow-up period. For other social care services other than meals on wheels, more people receiving conventional home care services were in receipt of each service compared with those in the reablement group. |
| Occupational Therapy in HomEcare Reablement Services (OTHERS) | Whitehead et al., 2016<br>England, UK | Step up and step down | Occupational therapist delivered reablement which was compared to social care worker delivered reablement | In favour of control group but not statistically significant. Higher hospital admissions in intervention group (27%) compared with the control group (17%) at 3 months, and higher admissions in the intervention group |  |  | *Community and social care resource use:* Total resource use at 6 months similar between groups. Intervention group less likely to use health services (including GP), an occupational therapist and |
|  |  |  |  | (40%) compared with the control group (17%) at 6 months. |  |  | access meals at home. |
| Restorative care | Tinetti et al., 2002 US | Step down | Home care agency staff (occupational therapist nurse, physio, home health aide) | | | | *Emergency department visits:*<br>**Statistically significant** difference in favour of the intervention group. There was a significant difference ($p < 0.001$) in emergency department visits between the intervention (10%) and control (20%) groups. |
| | Tinetti et al., 2012 US | Step down | Home care agency staff (occupational therapist, nurse, physio, home health aide) | In favour of the intervention group but not statistically significant. 13.2% of restorative model participants were readmitted to an acute hospital during the episode of home care, versus 17.6% of usual domiciliary care participants. According to the conditional regression analysis, participants receiving the restorative model of home care were<br>32% less likely to be<br>readmitted than<br>those receiving<br>usual domiciliary<br>care. | **Statistically significant** difference in favour of the intervention group. The mean length of home care episodes was $20.3 \pm 14.8$ days in the restorative care group and $29.1 \pm 31.7$ days in the usual domiciliary care group ($p < 0.001$). | | |

Text highlighted in bold indicates statistical significance P values are presented where available in published study articles.

**Table 5.**
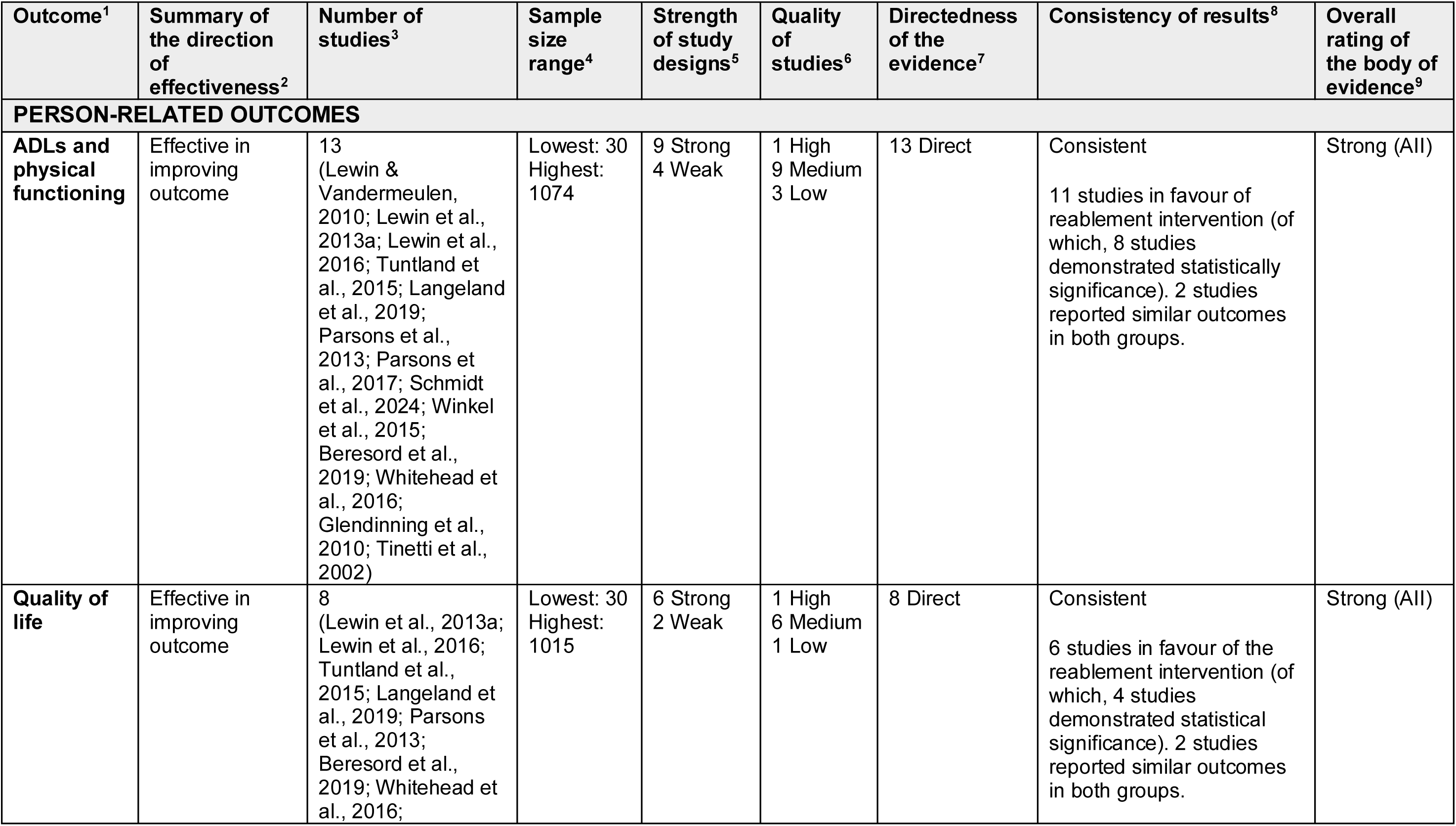
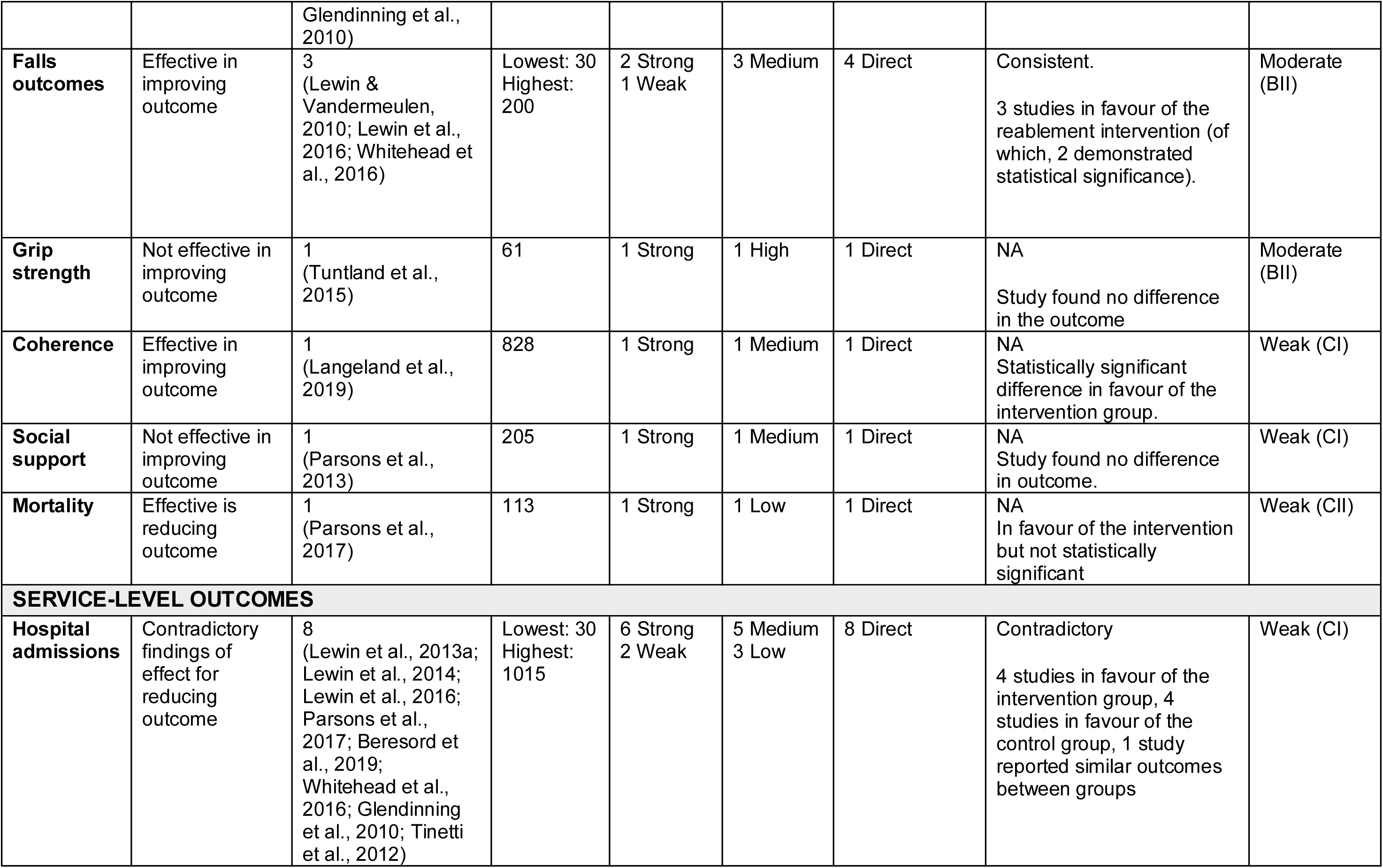
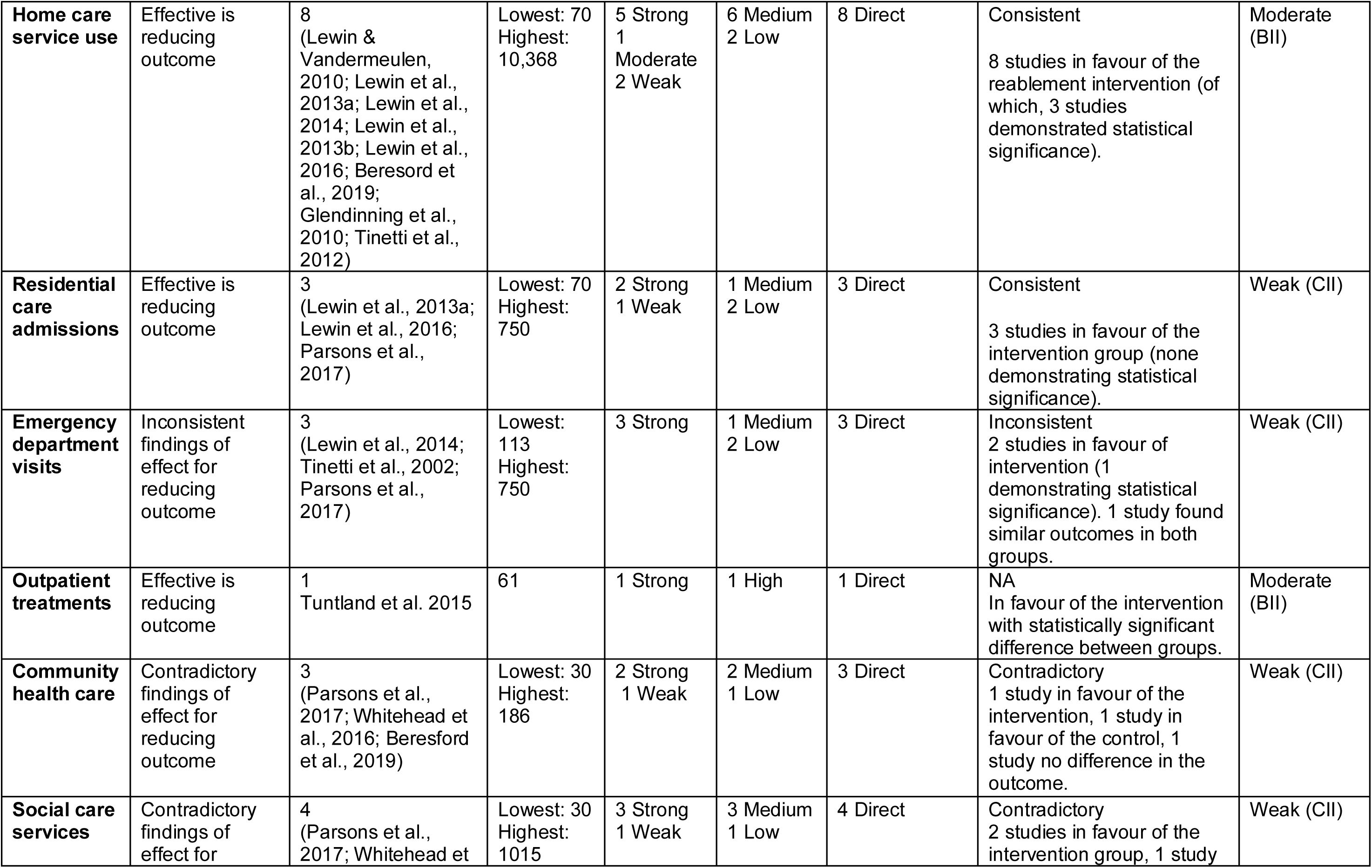
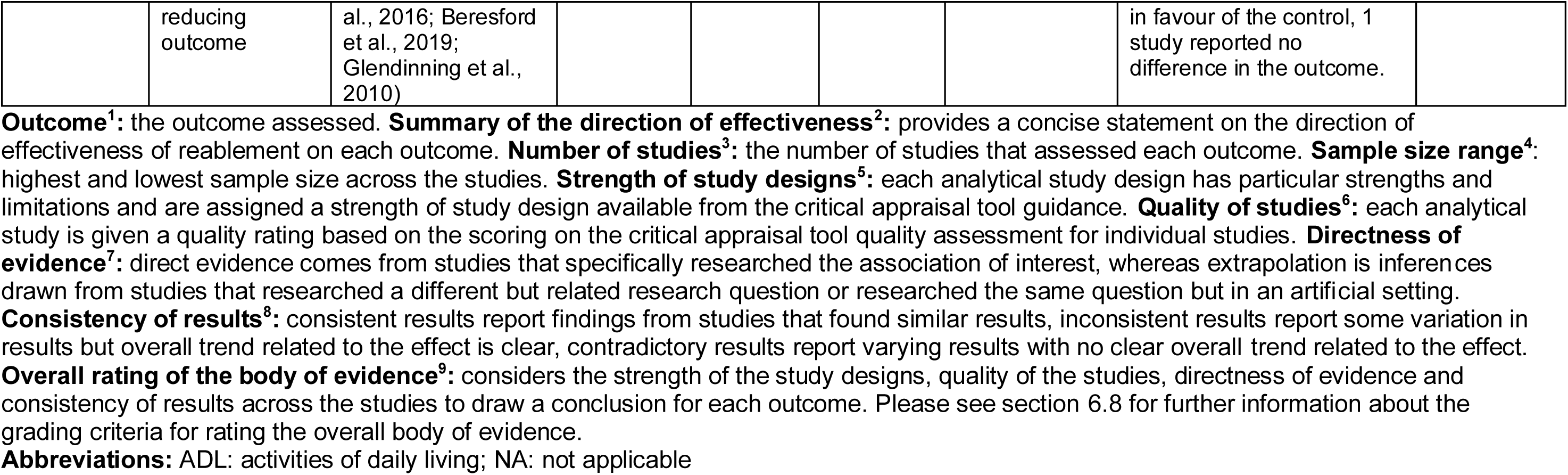
Summary of the rating of the certainty of the overall body of evidence for the effectiveness of reablement on person-related outcomes and service-level outcomes.

The quality of included studies varied. All studies had a clear, focused research question. Most studies (fifteen) evidenced a study population representative of the target population, however one study did not describe recruitment sufficiently to conclude whether its study population was representative. Most studies (ten) did not use a random sampling methodology, contributing to selection bias. Five studies allocated participants to groups systematically, of which four used randomisation processes for group allocation, which allowed for the adjustment and control of potential confounders. Twelve studies went some way to controlling for misclassification bias, while three studies were at high risk of misclassification. Eight studies reported a lack of a control group or major differences between control and intervention groups.

Five studies failed to adequately blind assessors and participants, thus were at risk of information bias. Most studies (nine) used validated and reliable data collection instruments and methods. Follow-up was poorly conducted across the studies, with seven studies that reported less than 80% completion rate at follow-up, which lead to significant missing data. However, the observed low completion rate at follow-up can likely be attributed to the sample characteristics, comprised of older individuals and consequently experienced higher levels of mortality. Follow-up periods were slightly different across the studies, with a mixture of short- and long-term follow-ups reported. Statistical testing was appropriate for the data used and hypotheses tested. Thirteen studies included sufficiently powered sample sizes to reliably detect statistically significant differences, while three studies failed to do so.

This review rated the overall quality of studies evaluating the effectiveness of reablement on person-related and service-level outcomes, and a significant variation was observed. Only one study (Tuntland et al., 2015) was rated as high quality. This study had a rigorous methodology and its results can be viewed as reliable. Most studies were rated as medium quality (11 studies), each with some limitations in their study designs. Four studies were rated as low quality, showing significant methodological weaknesses that failed to account for potential biases that could impact the reliability of the results. Detailed results from the critical appraisal of individual studies can be found in the appendices (section 9.2.1).

#### 2.2.3 The clinical effectiveness of reablement interventions on person-related outcomes

Table 3 summarises the main findings of clinical effectiveness on person-related outcomes. The following sections present the evidence under respective outcome categories of person-related outcomes.

##### Mobility including activities of daily living

Thirteen studies assessed the effectiveness of reablement on improving mobility and ADLs (Lewin & Vandermeulen, 2010; Lewin et al., 2013a; Lewin et al., 2016; Tuntland et al., 2015; Langeland et al., 2019; Parsons et al., 2013; Parsons et al., 2017; Schmidt et al., 2024; Winkel et al., 2015; Beresford et al., 2019; Whitehead et al., 2016; Glendinning et al., 2010; Tinetti et al., 2002). The feasibility RCT (referred to as Occupational Therapy in HomEcare Re-ablement Services (OTHERS)) compared a model of reablement led and delivered by occupational therapists with a model of reablement led and delivered by social care workers (Whitehead et al., 2016).

Three Australian studies assessed the effectiveness of the Home Independence Program (HIP) on improving mobility and ADLs (Lewin & Vandermeulen, 2010; Lewin et al., 2013a; Lewin et al., 2016). HIP is an intensive reablement intervention including multiple home visits delivered for up to 12 weeks. Firstly, a small non-randomised controlled trial (NRCT) assessed the effectiveness of HIP delivered in one metropolitan area of Perth, Australia.

Participants who received HIP included people first referred for home care or people in need of further home care assistance (Lewin & Vandermeulen, 2010). The intervention was led by a multi-professional team of health professionals, including occupational therapists, physiotherapists and nurses, with one member of the team working as the care manager to implement assessments, goal setting and strategies relevant to the clients’ needs and goals. ADLs (basic activities required for survival), instrumental ADLs (IADLS) (complex activities necessary for independent living) and the Timed Sit Up and Go test score (measure of mobility) were compared between the intervention and the control group who received home and community care (HACC) services. Significant difference between the groups in terms of change in ADL (p<0.001), IADL (p<0.001) and Timed Sit Up and Go (TUG) (p<0.001) scores between baseline and 3 months, and at 12 months: ADL (p=0.04), IADL (p<0.001), and TUG (p<0.001) were reported in favour of the intervention group (Lewin & Vandermeulen, 2010).

The second study of HIP, a large randomised controlled trial (RCT) conducted across different areas of Perth, assessed the effectiveness of the HIP intervention for people who had no prior receipt of home care services before HIP, compared with usual domiciliary care services (Lewin et al., 2013a). Overall, ADL scores were similar for both groups with both groups reporting improvements over time. At 3-month follow-up, there was no difference in ADL scores between the HIP and usual domiciliary care groups, with both groups demonstrating improvements in ADLs and IADLs from baseline to 3-month follow-up. At 12-month follow-up, there was a significant difference (p=0.016) in IADLs between groups, favouring the intervention (Lewin et al., 2013a).

The third study assessed the effectiveness of a new model of HIP whereby non-health professionals replaced health professionals as care managers to deliver the intervention under the new HIP-coordinator (HIPC) service model (Lewin et al., 2016). This study was an uncontrolled before and after study and therefore did not include a comparator group.

Between baseline and 3-months follow-up, HIPC was associated with statistically significant improvements in ADLs (p<0.0001) and IADLs (p<0.0001). There were also statistically significant improvements in ADLs (p=0.0003) and IADLs (p<0.0001) between baseline and 12 months, demonstrating a sustained improvement. A secondary analysis compared the results from HIPC with the results from the earlier RCT of HIP, where health professionals undertook the care manager role (Lewin et al., 2013a). The secondary analysis reported statistically significant differences in improvements in ADLs (p=0.0237) and IADLs (p=0.0159) between HIPC and HIP between baseline and 12 months, favouring HIPC when non-health professionals take on the role of care manager (Lewin et al., 2016).

Another study of a separate reablement intervention delivered in Australia assessed the effectiveness of a short-term restorative care programme (Schmidt et al., 2024). The intervention was delivered by allied health professionals for 8 weeks and included multi-professional team involvement. This was a moderate quality uncontrolled before and after study (UCBA) that assessed the difference in ADL and functional outcomes between baseline and following the intervention. The study did not include a comparator group, but intervention demonstrated statistically significant improvements (p<0.0001) between baseline and 8-week follow-up in ADLs and functioning as measured by the Modified Barthel Index, Lower Extremity Functional Scale, Upper Extremity Functional Index and Berg Balance Scale tools (Schmidt et al., 2024).

In Norway, two studies assessed the clinical effectiveness of a 10-week multicomponent reablement programme on self-perceived activity performance and satisfaction with performance (Tuntland et al., 2015; Langeland et al., 2019). Both studies used the Canadian Occupational Performance Measure (COPM) to assess self-perceived activity performance and satisfaction with performance. Firstly, Tuntland and colleagues conducted the first RCT of home-based reablement in Europe, conducted in one rural municipality in Norway. This was a high quality study, comparing a reablement programme where occupational therapists and physical therapists supervised home care personnel to deliver the intervention, to usual domiciliary care services. The reablement groups experienced statistically significant improvements in mean COPM performance scores at both 3 months (p=0.02) and 9 months (p=0.03). The reablement group also reported significantly higher COPM satisfaction scores at 9 months (p=0.03); however, no statistically significant differences in COPM satisfaction were found between groups at 3 months (Tuntland et al., 2015). A large non-randomised controlled trial (NRCT) also compared a 10-week multicomponent reablement programme with usual domiciliary care services (Langeland et al., 2019). This was a medium quality study conducted across 47 Norwegian municipalities. The intervention was delivered by homecare personnel and involved a multi-professional team who collaborated with the participants for the whole intervention period. The results reported significantly higher COPM performance (p<0.001) and COPM satisfaction (p<0.001) at 10 weeks and 6 months follow-up, compared to usual domiciliary care. There was no significant difference between groups for COPM performance and COPM satisfaction at 12 months.

A medium quality cluster RCT compared a restorative home care intervention with standard home care in New Zealand (Parsons et al., 2013). The intervention included allied health professional (AHP) involvement and was delivered by home care aids, but the study did not report on the duration of the intervention. A greater improvement in physical function scores between baseline and 6 months were reported for the intervention group (p=0.003). A second study conducted in New Zealand, a low quality RCT, compared the effectiveness of a Community Flexible Integrated Responsive Support Team (FIRST) intervention with usual domiciliary care (Parsons et al., 2017). The intervention was coordinated by a nurse case manager and was delivered by unregulated support worker/healthcare assistant. The study did not report on the involvement of other healthcare professionals such as AHPs, nor did it provide information on the duration of the reablement, or any details about the comparator group. The findings reported similar functional outcome scores, ADLs or IADLs in both groups (Parsons et al., 2017).

In Denmark, a low quality UCBA study assessed the effectiveness of the Copenhagen Municipality Reablement Program for improving participants’ ability to perform ADLs (Winkel et al., 2015). The study also explored whether the outcomes differed between participants who had already received home care services and had applied for more help (help group) and those who had not received home care services prior to the reablement program and had applied for home help (no help group). The results reported significant improvements in ADLs between baseline and the end of the reablement intervention (p=0.042) with improvements being maintained at 10 months post-intervention (p=0.042). There were no significant differences in ADL scores between the ‘help’ and ‘no help’ groups (p=0.120) (Winkel et al., 2015).

Two UK studies assessed the effectiveness of reablement on ADLs and mobility (Beresford et al., 2019; Glendinning et al., 2010). Another UK study reporting on ADLs compared a model of reablement led and delivered by occupational therapists with a model of reablement led and delivered by social care workers (Whitehead et al., 2016). A medium quality non-randomised controlled trial evaluated five models of reablement services in England (Glendinning et al., 2015). Each model included the involvement of multi-professional teams with management structures consisting of managers, multi-professional specialists (occupational therapists, physiotherapists and senior carers). Differences in effectiveness between the reablement models were not reported. At 12-month follow-up, participants in the standard care comparison group were less able to undertake all ADLs compared to participants in the reablement group. Results found statistically significant differences between the groups (in favour of the intervention group) for some outcomes, including being unable to get outdoors and walk down the road (p<0.001), and unable to bath, shower or wash all over (p<0.001) (Glendinning et al., 2015).

Another study, a medium quality uncontrolled before and after (UCBA) study, assessed the effectiveness of models of English reablement (Beresford et al., 2019). Descriptive statistics indicated in terms of improvements in ADL outcomes between baseline and 6 months follow-up, as measured by the Barthel Index and the Nottingham Extended Activities of Daily Living Scale (NEADL) scale (Beresford et al., 2019).

A medium quality feasibility RCT (referred to as OTHERS) conducted in England compared a model of reablement led and delivered by occupational therapists with a model of reablement led and delivered by social care workers under the direction of a reablement care team leader (Whitehead et al., 2016). There were improvements in ADL outcomes (Barthel Index, Nottingham Extended Activities of Daily Living Scale (NEADL) scale) from baseline in both groups, although overall improvements were greater in the occupational therapist intervention group. This was a feasibility study with a small sample (Whitehead et al., 2016).

A non-randomised controlled study assessed the effectiveness of step-down restorative care in the US (Tinetti et al., 2002). This study was of medium quality and compared restorative care delivered by home care agency staff (occupational therapists, nurses, physiotherapists and home health aides) with usual domiciliary care. The intervention group reported significantly higher mean mobility scores (p=0.02) and higher mean self-care ADLs (p=0.07) compared to usual domiciliary care) (Tinetti et al., 2002).

##### Quality of life

Eight studies reported on the effectiveness of reablement services on Quality of Life (QoL) outcomes (Glendinning et al., 2010; Lewin et al,. 2013a; Parsons et al., 2013; Tuntland et al., 2015; Whitehead et al., 2016; Lewin et al., 2016; Langelend et al., 2019 and Beresford et al., 2019). Specifically, the study by Whitehead and colleagues compared a model of reablement led and delivered by occupational therapists with a model of reablement led and delivered by social care workers.

A high-quality comparative before-after study assessing five models of reablement services provided by local authorities in England compared with standard home care (Glendinning et al., 2010) found significant improvement in HRQoL (derived from the EQ-5D) between baseline and 12 months (p<0.001) in the intervention group and found no change within the comparison group. No statistically significant differences were identified in social care-related quality of life (derived through the ASCOT tool) between the reablement and comparison groups.

The low quality RCT conducted in Australia (Lewin et al., 2013a) comparing the HIP reablement programme with HACC usual domiciliary care found similar QoL outcomes between groups. Both groups reported improvements in QoL, however no figures or statistical analysis are presented in the article.

A medium quality cluster RCT compared a restorative home care intervention with standard home care in New Zealand (Parsons et al., 2013). This study found improved QoL outcomes amongst individuals receiving the restorative home care intervention. Between baseline and 6 months, there was greater change in HRQoL (as measured by the SF-36) in the intervention group (p=0.0001) than the control group.

A high quality RCT analysing a multicomponent home-based reablement programme in Norway (Tuntland et al., 2015) reported similar HRQoL outcomes between intervention and control groups (as measured by the COOP/Wonka). People in both groups evidenced improvement in HRQoL across most domains up to 9-month follow-up.

A medium quality feasibility RCT of OTHERS conducted in England compared a model of reablement led and delivered by occupational therapists with a model of reablement led and delivered by social care workers (Whitehead et al., 2016) found non-statistically significant improvements in HRQoL (measured by the EQ-5D) and social care related QoL (measured by the ASCOT tool) from baseline in both groups, although overall improvements were greater in the occupational therapy intervention group (care provided by an occupational therapist).

A moderate quality uncontrolled before-after study (Lewin et al., 2016) assessing the impact of the HIPC and HIP services in Australia reported statistically significant improvements in QoL (as measured by the Assessment of Quality of Life outcome measure) in the single cohort group between baseline and 3 months (p< 0.0001) and between baseline and 12 months (p< 0.0001). This increase however was not statistically significant between the 3 and 12-month follow-up periods.

A medium quality non-randomised controlled trial comparing a 10-week multicomponent reablement programme with usual domiciliary care services in Norway (Langeland et al., 2019) found a positive significant difference (p<0.001) in some domains of the EQ-5D (mobility, personal care, usual activities and current health) at 6-month follow-up. No difference between groups for other domains or at other time points were evidenced.

##### Falls outcomes

Three studies reported on the effectiveness of reablement on falls outcomes (Lewin & Vandermeulen, 2010; Lewin et al., 2016; Whitehead et al., 2016). Specifically, the study by Whitehead and colleagues compared a model of reablement led and delivered by occupational therapists with a model of reablement led and delivered by social care workers.

In Australia, a medium quality non-randomised controlled trial reported the HIP programme, when delivered by health care professionals, had a positive impact on falls efficacy, defined as the perceived ability of individuals to perform activities without losing balance or falling. At 3 months, HIP scored significantly higher than the usual domiciliary care services control group on the modified falls efficacy scale (p=0.034) (Lewin & Vandermeulen, 2010). In a later medium quality study assessing the effectiveness of HIPC, where non-health professionals acted as care managers to deliver the reablement instead of health care professionals, HIPC demonstrated statistically significant improvements in the modified falls efficacy scale between baseline and 3 months (p< 0.0001) and between baseline and 12 months (p< 0.0001) (Lewin et al., 2016).

In the UK, a medium quality feasibility RCT of OTHERS reported fewer falls in the occupational therapist reablement group compared to the social care worker reablement group in terms of the number of participants who reported a fall and the mean number of falls reported at both 3 and 6 month follow-up periods (Whitehead et al., 2016).

##### Other person-related outcomes

Other person-related outcomes reported in the identified studies included grip strength (Tuntland et al., 2015), sense of coherence (Langeland et al., 2019), social support (Parsons et al., 2013), and mortality (Parsons et al., 2017).

The first RCT of home-based reablement in Europe (a high quality RCT) conducted in one rural municipality in Norway compared grip strength between a 10-week reablement programme where occupational therapists and physical therapists supervised home care personnel to deliver the intervention, with usual domiciliary care services. Neither group reported any improvements in grip strength at 3 or 9 months follow-up (Tuntland et al., 2015).

Langeland and colleagues conducted a large non-randomised controlled trial (medium quality) comparing a 10-week multicomponent reablement programme with usual domiciliary care services in Norway. Coping related to experiences of coherence (comprehensibility, manageability and meaning) was assessed using the Sense of Coherence Questionnaire (SOC-13). This paper reported a statistically significant difference, in favour of the intervention group at 3-months (p=0.04), but no statistically significant differences between groups at 12-month follow-up (p=0.72) (Langeland et al., 2019).

In New Zealand, a medium quality cluster RCT compared a restorative home care intervention with standard home care (Parsons et al., 2013). The intervention included allied health professional (AHP) involvement and was delivered by home care aides. No significant differences (p=0.09) in social support (measured using the Dukes Social Support Index) was reported between groups at 6-months follow-up (Parsons et al, 2013).

A second study conducted in New Zealand, a low quality RCT, compared the effectiveness of a Community Flexible Integrated Responsive Support Team (FIRST) intervention with usual domiciliary care (Parsons et al., 2017). The intervention was coordinated by a nurse case manager and was delivered by an unregulated support worker/healthcare assistant. The outcome of death was lower in this intervention group, compared to the control group, but this was not statistically significant (Parsons et al., 2017).

A high quality prospective cohort study assessing models of reablement in England (Beresford et al., 2019) found HRQoL improvements amongst those receiving reablement care. Outcomes were assessed at entry to the reablement services (T0), at discharge from the reablement service (T1), and at 6 months post discharge (T2). Descriptive statistics indicated improvements in HRQoL and social care related QoL outcomes (measured by the EQ-5D-5L and ASCOT, respectively) between timepoints. The differences in mean EQ-5D-5L and EQ-5D Visual Analogue Scale between baseline and 6 months follow up were 0.15 and 17.77, respectively.

#### 2.2.4 Bottom line results for the effectiveness of reablement on person-related outcomes

The identified evidence was conducted in six different countries, with Australia contributing the highest number of research studies (four). The evidence was mainly focused on mobility, including ADLs (13 studies). The second most commonly reported outcome assessed was QoL (8 studies). Falls outcomes, assessed in 3 studies, indicated a lack of evidence on the effectiveness of reablement to prevent falls. Of the 13 studies reporting person-related outcomes, 7 studies assessed step-up reablement interventions, 3 studies assessed step-up and step-down reablement interventions, 1 study assessed step-down only, and 2 studies were unclear due to insufficient information on recruitment strategies and participant characteristics reported. Findings for each person-related outcome were graded and are summarised below and shown in Table 5. Grading was based on guidance by the Public Health Agency of Canada (2014) using the Critical Appraisal Toolkit (CAT). For more information see section 6.8.

- Strong international evidence (grade AII) from 13 studies indicated that reablement was effective in improving **mobility and activities of daily living**. Eleven of the studies reported improvements in mobility and ADLs, of which, eight studies found statistically significant improvements. Two of the studies reported similar outcomes between groups.
- Strong international evidence (grade AII) from eight studies suggested that reablement interventions were effective for increasing quality of life among clients. Six of the studies reported improvements in **quality of life** outcomes, and of these, six studies found statistically significant improvements. Two studies reported similar quality of life scores between groups.
- Moderate evidence (grade BII) from three studies from Australia and the UK suggested that reablement may improve outcomes to prevent **falls**. All three studies reported improvements in outcomes, with two studies reporting statistically significant findings.
- Moderate evidence (grade BII) from one high quality Norwegian study reported no difference in **grip strength** between the reablement intervention and the usual domiciliary care group.
- Weak evidence (grade CI) from a study conducted in Norway reported statistically significant improvements in sense of **coherence** in the reablement intervention group.
- Weak evidence (grade CI) from one study conducted in New Zealand found that reablement had no effect on increasing clients’ **social support**.
- Weak evidence (grade CII) from one study from New Zealand reported that **mortality** was lower in the reablement intervention group compared with usual domiciliary care, but the results were not statistically significant.

#### 2.2.5 Effectiveness of reablement on service-level outcomes

Table 4 summarises the main findings of clinical effectiveness on service-related outcomes. The following sections present the evidence under respective categories of service-related outcomes.

##### Admissions to residential care

Three identified studies assessed the impact of reablement services on admissions to residential care. Two studies were RCTs, one conducted in Australia (Lewin et al., 2013a) and the other in New Zealand (Parsons et al., 2017). The third was an uncontrolled before-after study conducted in Australia (Lewin et al., 2016).

One low quality RCT conducted in Australia compared the HIP reablement programme with home and community care (HACC) services (Lewin et al., 2013a). People receiving the HIP reablement programme were found to have a reduced rate of admissions to residential care compared to HACC. There was an equal number of participants in the reablement and HACC groups (Table 2). At 3 months, there were 21 and 16 admissions to residential care in the control and intervention groups, respectively. At 12 months, there were 48 and 44 admissions in the control and intervention groups, respectively. Nevertheless, the paper did not provide information to determine whether the admissions reported related to multiple admissions for the same individual.

A low quality RCT conducted in New Zealand compared the FIRST reablement intervention with usual domiciliary care (Parsons et a., 2017). People receiving the FIRST reablement intervention had a lower rate of placement in residential care than those receiving usual domiciliary care (17 in the FIRST intervention group, 22 in the usual domiciliary care group). However, this lower rate was not statistically significant.

A moderate quality uncontrolled before-after study assessed the impact of a Home Independence Program Coordinator (HIPC) service in Australia compared to HIP (Lewin et al., 2016). People who received the HIPC service were less likely to be admitted to residential care, however this was not statistically significant. A secondary comparison with the group of people who received the HIP programme in the RCT (Lewin et al., 2013a) reported slightly less transfers to residential care amongst people receiving HIPC.

##### Long-term home care needs

Eight identified studies evaluated the impact reablement services had on the requirement for future home care services: one RCT (Lewin et al., 2013a), one longitudinal study based on RCT data (Lewin et al., 2014), two non-randomised RCTs (Lewin and Vendermeulen, 2010; Tinetti et al., 2012), one retrospective cohort study (Lewin et al., 2013b), one uncontrolled before-after study (Lewin et al., 2016), one comparative before-after study (Glendinning et al., 2010), and one prospective cohort study (Beresford et al., 2019).

A moderate quality non-randomised controlled trial conducted in Australia compared the HIP reablement programme with usual home-care services (Lewin and Vendermeulen, 2010).

People who received HIP had statistically significantly lower odds of requiring home care services after the programme compared to people who received usual domiciliary care. Those who received HIP were 0.07 (p<0.001) times as likely to require home care services at 3 months and 0.14 (p<0.001) times at 12 months follow-up, compared to the control group.

A low quality RCT conducted in Australia (Lewin et al., 2013a) also compared the HIP reablement programme, but in their study usual domiciliary care was represented by HACC usual domiciliary care. The HIP programme was found to substantially and significantly decrease the odds of having an ongoing home care service at both 3 months (Odds Ratio: 0.18, 95% CI: 0.13-0.26) and 12 months (Odds Ratio: 0.22, 95% CI: 0.15-0.32) (p<0.001 for both).

Lewin et al. (2014) presented the long-term outcomes (2 year follow-up) of the low quality RCT conducted in Australia comparing the HIP reablement programme with HACC usual domiciliary care (Lewin et al., 2013a). Those in receipt of the HIP programme used significantly less home-care hours compared to the control group (p<0.001) during the first year (19.1 compared to 45.6, p<0.001) and second year (13.4 compared to 36.2, p<0.001), and overall after 24 months (29.8 compared to 74.4, p<0.001).

A moderate quality retrospective cohort study conducted in Australia assessed the outcomes of people who received two models of reablement support: HIP and Personal Enablement Program (PEP) (Lewin et al., 2013b). HIP is a multi-component reablement service targeted at elderly people referred from the community, while PEP uses the HIP service model but is targeted at elderly people discharged from hospital. Compared to standard home care, both HIP and PEP groups used fewer home care services in the first 3 years, with HIP remaining lower in year 3. Both HIP and PEP used less services over 57 months, with HIP still lower in the final year.

The moderate quality uncontrolled before-after study by Lewin et al. (2016) assessed the impact of the HIPC and HIP services in Australia. Secondary analysis found that both groups reported a reduction in people who required ongoing home care services at 3 and 12 months. However, outcomes for the HIPC groups were somewhat better than the HIP group with more people in the HIPC group no longer required home care services at 3 months (51.43% vs. 36.45%) and 12 months (65.71% vs. 39.68%).

A high quality prospective cohort study assessed models of reablement in England (Beresford et al., 2019). The study contained no comparator, but reablement services produced positive, but not statistically significant differences in outcome over the study period. A lower number of home care service hours required was reported between baseline (mean hours: 3.09) and 6 month follow-up (mean hours: 0.5).

A high quality comparative before-after study assessed five models of reablement services provided by local authorities in England compared with standard home care (Glendinning et al., 2010). The percentage of people who received independent home care was lower in the reablement group than the comparison group both within the first eight weeks (2% and 65%, respectively) and the remaining ten months (18% and 50%, respectively).

A moderate quality non-randomised controlled trial from the US assessed restorative care models and usual domiciliary care (Tinetti et al., 2012). The restorative care model that was evaluated was developed based on principles adapted from geriatric medicine, nursing, rehabilitation, goal attainment, chronic care management and behavioural change theory.

The intervention team consisted of a multi-professional team of home care clinicians. Home care episodes were significantly lower for people in receipt of the restorative care group compared to usual domiciliary care. The mean length of home care episodes was 20.3 ± 14.8 days in the restorative care group and 29.1 ± 31.7 days in the usual domiciliary care group (p< 0.001).

##### Hospital admissions

Eight identified studies assessed the impact reablement services had on hospital admissions: two RCTs (Parsons et al., 2017; Lewin et al., 2013a), one longitudinal study based on RCT data (Lewin et al., 2014), one uncontrolled before-after study (Lewin et al., 2016), one prospective cohort study (Beresford et al., 2019), one feasibility parallel group randomised controlled trial (Whitehead et al., 2016), one comparative before-after study (Glendinning et al., 2010), and one non-randomised controlled trial (Tinetti et al., 2012). The study by Whitehead and colleagues compared a model of reablement led and delivered by an occupational therapist compared with a model of reablement delivered by social care workers.

One low quality RCT conducted in Australia compared the HIP reablement programme with HACC usual domiciliary care (Lewin et al., 2013a). The study found an increased, but not statistically significant rate of hospital admission at 3 months for people who received the HIP programme when compared to usual domiciliary care. At 3 months, there were 6 and 24 hospital admissions in the control and intervention groups, respectively. At 12 months people who received usual domiciliary care were more likely to be admitted to hospital, with 3 hospital admissions recorded in the usual domiciliary care group and 1 recorded in the HIP group.

Lewin et al. (2014) presented the long-term outcomes of the low quality RCT conducted in Australia comparing the HIP reablement programme with HACC usual domiciliary care (Lewin et al., 2013a). There were fewer hospitalisations in the intervention group compared with the control group during the first and second year, and overall at 24 months.

A moderate quality uncontrolled before-after study (Lewin et al., 2016) assessed the impact of the HIPC and HIP services in Australia. The study found lower hospitalisations in the HIPC group compared to HIP at 3 months (1 (1%) compared to 21 (7%)). As a proportion of each group, admissions increased in the HIPC group compared to HIP at 12 months (4 (6%) compared to 12 (4%)). These differences were not statistically significant. Secondary comparison reported slightly less hospitalisations than results from the previous RCT (Lewin et al., 2013a).

A low quality RCT conducted in New Zealand compared the FIRST reablement intervention with usual domiciliary care (Parsons et al., 2017). The RCT found no difference in hospitalisations between people who received the FIRST intervention and people who received usual domiciliary care. Both groups reported similar hospital admissions over 12 months. No numerical figures are offered in the paper.

A high quality prospective cohort study assessed models of reablement in England (Beresford et al., 2019). The study contained no comparator, but reablement services produced positive, but not statistically significant differences in the number of hospital admissions and hospital length of stay. Mean number of hospital visits without overnight stay fell between baseline (0.31) and 6-month follow up (0.18). Mean hospital length of stay at baseline was 2.32 days and reduced to 0.16 days at 6-month follow-up.

A moderate quality feasibility parallel group randomised controlled trial compared a home care reablement service led and delivered by occupational therapists with reablement led and delivered by social care workers (control group) in England (Whitehead et al., 2016). The trial found a higher number of hospital admissions in the group that received the OTHERS reablement service (27%) than in the control group (17%) at 3 months follow-up. The same trend held at 6-month follow-up, with (40%) of those in the intervention group admitted to hospital, compared to (17%) in the control group.

A high quality comparative before-after study assessed five models of reablement services provided by local authorities in England compared with standard home care (Glendinning et al., 2010) found higher hospital inpatient stays amongst people who received reablement services. At 10-month follow-up, the intervention group had higher hospital inpatient stays (237 days) compared with the comparison group (139 days). This difference was not statistically significant.

A moderate quality non-randomised controlled trial from the US assessed restorative care models and usual domiciliary care (Tinetti et al., 2012). The study found people who received restorative care were less likely to be readmitted to an acute hospital during their episode of home care. 13.2% of restorative model participants were readmitted to an acute hospital during the episode of home care, versus 17.6% of usual domiciliary care participants. Conditional regression analysis suggested participants who received the restorative model of home care were 32% less likely to be readmitted than those who received usual domiciliary care.

##### Other health and social care resource use

Seven identified studies assessed the impact reablement services had on other forms of health and social care resource use: one longitudinal study based on RCT data (Lewin et al., 2014), two RCTs (Parsons et al., 2017; Tuntland et al., 2015), one prospective cohort study (Beresford et al., 2019), one feasibility parallel group randomised controlled trial (Whitehead et al. 2016), and one comparative before-after study (Tinetti et al., 2002).

Lewin et al. (2014) presented the long-term outcomes of the low quality RCT conducted in Australia that compared the HIP reablement programme with HACC usual domiciliary care (Lewin et al., 2013a). The longer-term outcomes of the RCT were that fewer emergency department presentations were recorded in the intervention group (188 visits, 50% of group) compared to the usual domiciliary care group (208 visits, 58% of group) during the first year. There were also a lower number of emergency department visits among the HIP group (239 visits, 64% of the group) compared to the usual domiciliary care group (257 visits, 69% of group). However, these differences were not statistically significant.

A high quality RCT analysed a multicomponent home-based reablement programme in Norway (Tuntland et al., 2015). The study found a statistically significant difference (p=0.007) in outpatient treatments between the groups at 3-month follow-up: control group (12 treatments) vs intervention (3 treatments).

A low quality RCT conducted in New Zealand that compared the FIRST reablement intervention with usual domiciliary care (Parsons et al., 2017) found a non-significant difference in other social care resource use between control and intervention groups. Over 12 months, those in the intervention group used more day care, day centre, meals on wheels and transport and slightly more general practitioner (GP) visits. The number of emergency department presentations were similar in both groups. No data is shown in the paper to report figures.

A high quality prospective cohort study assessed models of reablement in England (Beresford et al., 2019). As a cohort study there was no comparator, but the intervention produced positive, but non-statistically significant differences in service outcomes between timepoints. There was a reduction in mean community care visits from entry to the reablement service (2.08 visits) to 6 month follow up (0.9 visits). There was also a reduction in mean social care service visits between entry to the reablement service (0.92 visits) and 6-month follow up (0.72 visits).

A moderate quality feasibility parallel group randomised controlled trial compared a home care reablement service led and delivered by occupational therapists with a control group who received reablement which was led and delivered by social care workers (Whitehead et al., 2016). Total resource use at 6 months follow-up was similar between groups. Those in the OTHERS group were less likely to use community and social care services (including GP services) (60% of participants in the intervention group used health services, compared to 100% of participants in the control group), an occupational therapist (0% of participants in the OTHERS group compared to 17% of participants in the control group) and accessed meals at home (10% of participants in the OTHERS group compared to 25% of participants in the control group).

A high quality comparative before-after study assessed five models of reablement services in England compared with usual domiciliary care (Glendinning et al., 2010). This study found those in receipt of reablement services used less other social care services than those receiving standard home care. Of 241 participants in the reablement group and 141 participants in the control group that completed follow-up assessments, six people in the comparison group and none in the reablement group used day care services during the 10-month follow-up period. For other social care services other than meals on wheels, more people receiving conventional home care services were in receipt of each service compared with those in the reablement group.

A high quality non-randomised controlled trial compared restorative home care reablement with usual domiciliary care in the US (Tinetti et al., 2002). Those in receipt of the reablement intervention evidenced a statistically significant difference (p<0.001) in emergency department visits between the intervention (10% utilisation rate) and control (20% utilisation rate) groups.

#### 2.2.6 Bottom line results for the effectiveness of reablement on service level outcomes

The identified evidence was conducted in five different countries, with Australia contributing the highest number of research studies (five studies). The evidence of service level outcomes was mainly focused on long-term home care needs (8 studies) and hospital admissions (8 studies). Other health and social care resource use (7 studies) and admissions to residential care (3 studies) were also reported. A lack of evidence in reporting the effectiveness of reablement services on reducing admissions to residential care was identified. Of the 12 studies that reported service level outcomes, 6 studies assessed step-up reablement interventions, 4 studies assessed step-up and step-down reablement interventions and 2 studies assessed step-down only. Findings for each person-related outcome were graded and are summarised below and are shown in Table 5.

- Weak international evidence (grade CI) from eight studies reported contradictory evidence (see glossary of terms) on the effectiveness of reablement to reduce **hospital admissions**.
- Moderate international evidence (grade BII) from eight studies reported consistent findings demonstrating the effectiveness of reablement to reduce the use of ongoing **home care services**, with three studies reporting statistically significant findings.
- Weak international evidence (grade CII) from three studies from Australia (n=2) and New Zealand (n=1) suggested that reablement was effective in reducing **residential care admissions**, but none of the studies reported statistically significant findings.
- Weak international evidence (grade CII) from three studies reported inconsistent findings on the effectiveness of reablement to reduce **emergency department visits**, with two studies reporting favourable outcomes for the reablement group (one demonstrating statistically significant findings), and one study reporting similar outcomes in both groups.
- Moderate evidence (grade BII) from one Norwegian study found that reablement was effective in reducing the number of **outpatient treatments** compared with usual domiciliary care with statistically significant differences between groups.
- Weak evidence (grade CII) from three studies conducted in England (n=2) and New Zealand (n=1) reported contradictory findings on the effectiveness of reablement to reduce **community care service use**.
- Weak evidence (grade CII) from four studies conducted in England (n=3) and New Zealand (n=1) reported contradictory findings on the effectiveness of reablement to reduce **social care service use**.

### 2.3 Cost-effectiveness of reablement interventions

#### 2.3.1 Overview of the evidence on the cost-effectiveness of reablement interventions

Three economic evaluations reporting on the cost-effectiveness of reablement were included in this review. The study characteristics of the included economic evaluations are presented in Table 6. The first economic evaluation was a cost-effectiveness analysis (CEA) of five English reablement services (Glendinning et al., 2010). The second economic evaluation was a model-based cost-minimisation analysis (CMA) conducted in England (Bauer et al., 218) and was based on differences in service-use outcomes reported in a RCT of the Home Independence Program in Australia (Lewin et al., 2013a). The final economic evaluation, a trial-based CEA, assessed the cost-effectiveness of a 10-week multicomponent reablement intervention in Norway (Kjerstad and Tuntland, 2016).

**Table 6:**
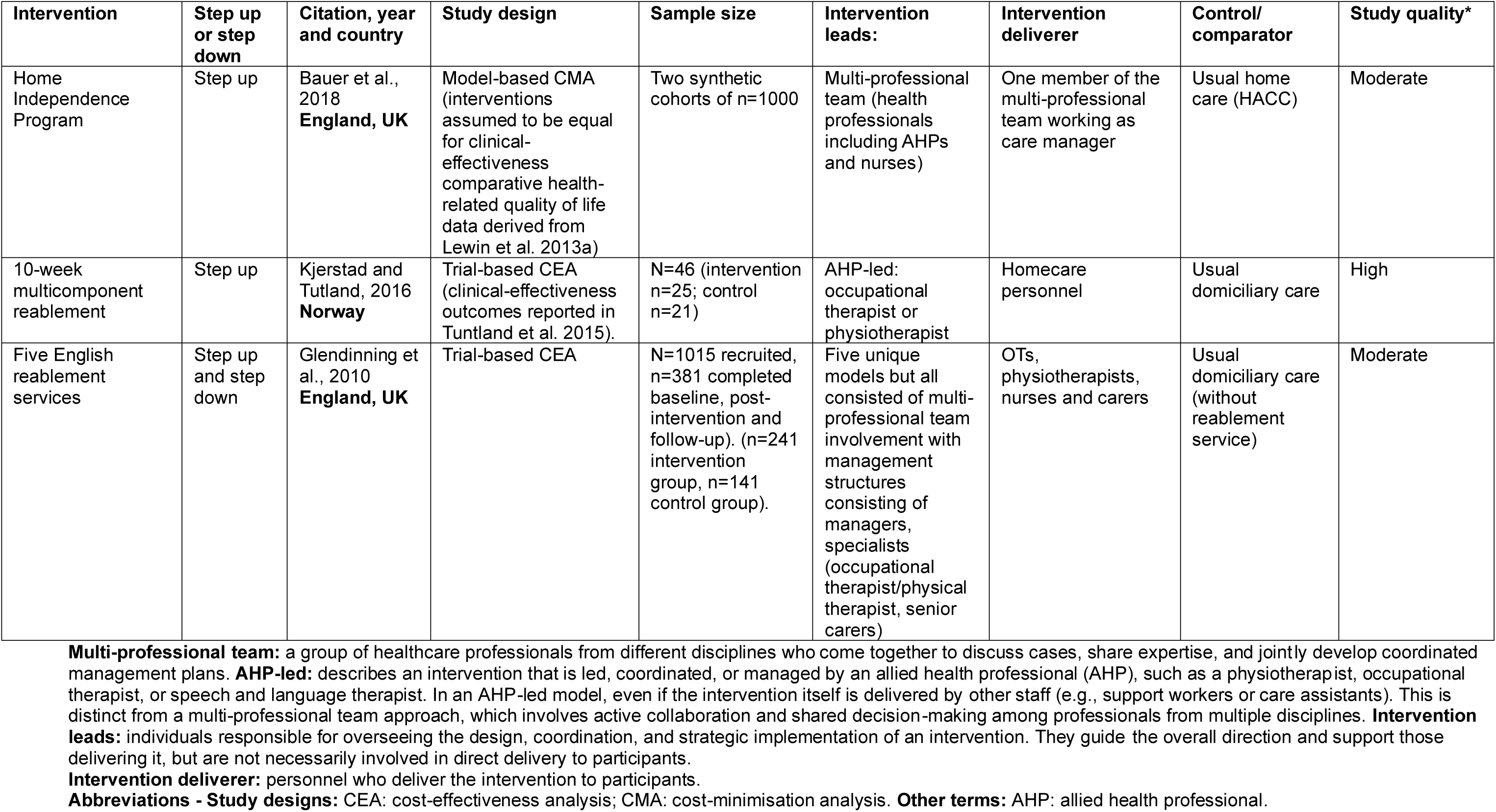
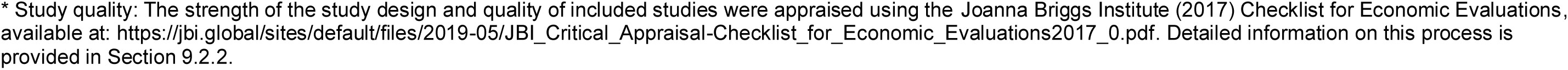
Study characteristics of economic evaluation studies assessing cost-effectiveness (n=3)

| Intervention | Step up or step down | Citation, year and country | Study design | Sample size | Intervention leads: | Intervention deliverer | Control/ comparator | Study quality* |
| --- | --- | --- | --- | --- | --- | --- | --- | --- |
| Home Independence Program | Step up | Bauer et al., 2018<br>**England, UK** | Model-based CMA (interventions assumed to be equal for clinical-effectiveness comparative health-related quality of life data derived from Lewin et al. 2013a) | Two synthetic cohorts of n=1000 | Multi-professional team (health professionals including AHPs and nurses) | One member of the multi-professional team working as care manager | Usual home care (HACC) | Moderate |
| 10-week multicomponent reablement | Step up | Kjerstad and Tutland, 2016<br>**Norway** | Trial-based CEA (clinical-effectiveness outcomes reported in Tuntland et al. 2015). | N=46 (intervention n=25; control n=21) | AHP-led: occupational therapist or physiotherapist | Homecare personnel | Usual domiciliary care | High |
| Five English reablement services | Step up and step down | Glendinning et al., 2010<br>**England, UK** | Trial-based CEA | N=1015 recruited, n=381 completed baseline, post-intervention and follow-up). (n=241 intervention group, n=141 control group). | Five unique models but all consisted of multi-professional team involvement with management structures consisting of managers, specialists (occupational therapist/physical therapist, senior carers) | OTs, physiotherapists, nurses and carers | Usual domiciliary care (without reablement service) | Moderate |

**Intervention deliverer:** personnel who deliver the intervention to participants.
**Abbreviations - Study designs:** CEA: cost-effectiveness analysis; CMA: cost-minimisation analysis. **Other terms:** AHP: allied health professional.
\* Study quality: The strength of the study design and quality of included studies were appraised using the Joanna Briggs Institute (2017) Checklist for Economic Evaluations, available at: https://jbi.global/sites/default/files/2019-05/JBI_Critical_Appraisal-Checklist_for_Economic_Evaluations2017_0.pdf. Detailed information on this process is provided in Section 9.2.2.

#### 2.3.2 Quality of studies evaluating the cost-effectiveness of reablement interventions

Two of the included economic evaluation studies were deemed to be of moderate quality when critically appraised by the Joanna Briggs Institute (JBI) checklist for economic evaluations (Bauer et al., 2018; Glendinning et al., 2010), see the appendices (Section 9.2.2). The economic evaluation conducted by Kjerstad and Tuntland (2016) was deemed to be of strong quality.

The CEA conducted by Glendinning et al. (2010) contained key methodological flaws as it included improper application of the NICE threshold (using non-parameterised measures) and did not calculate EQ-5D utility values or QALYs.

The CMA by Bauer et al. (2018) used evidence from the moderate quality analyses by Glendinning et al. (2010) and Australian data from Lewin et al. (2013a). The moderate quality evidence underpinning the model-based economic analysis by Bauer et al. (2018) restricted the quality and representativeness of the results. The authors did acknowledge the lack of quality evidence on which to base their model.

The CEA undertaken by Kjerstad and Tuntland (2016) was conducted more rigorously and contained fewer methodological flaws. However, a key methodological flaw in this study was the lack of sensitivity analysis conducted in order to investigate uncertainty in their estimates.

#### 2.3.3 The cost-effectiveness of reablement interventions

Three studies conducted economic evaluations of home-based reablement services compared to standard home care only. Two evaluations took the form of trial-based cost-effectiveness analyses (Kjerstad and Tuntland, 2016; Glendinning et al., 2010) and the other, a model-based cost-minimisation analysis (Bauer et al., 2018). Table 7 summarises the main findings on the cost-effectiveness of reablement interventions.

**Table 7:** Summary of main cost-effectiveness findings.

| Cost-effectiveness |  |  |  |  |  |
| --- | --- | --- | --- | --- | --- |
| Intervention name | Citation, year, country | Outcomes | Effectiveness | Costs | Cost-effective? |
| 10-week multicomponent reablement | Kjerstad and Tutland, 2016 Norway | Performance and satisfaction of ADLs | Intervention group more effective than comparator group | Lower costs reported for intervention group compared to comparator group. | Yes - The incremental cost-effectiveness ratio (ICER) reported that the cost per one-point improvement in the COPM-performance was 868.18 NOK lower on average for the intervention group compared with the control group. The cost per one-point improvement in the COPM-satisfaction was 666.30 NOK lower on average for the intervention group compared with the control group. |
| Home Independence Program | Bauer et al., 2018 England, UK | Model parameters for the relative effects of difference in service use were taken from the Australian randomised controlled trial (Lewin et al., 2013a). | No difference in HRQoL between groups (based on outcomes from Lewin et al., 2013a). | Lower costs reported for intervention group | Yes – when effectiveness is assumed as equal between groups. The overall probability that reablement was cost-reducing (i.e. leading to potential cost savings compared to standard home care) was 94.5% (95% CI 93.1 to 95.9). High probability that reablement is cost saving compared to |
| Five English reablement services | Glendinning et al., 2010 England, UK | HRQoL (EQ-5D) | Intervention group more effective | Intervention associated with higher costs from NHS and social perspective, and lower costs from social care perspective only | Yes - at the £30,000 per QALY gained threshold, the probability of reablement being cost-effective is 99% for total costs and just under 100% for social care costs only. At the more stringent £20,000 threshold, in the total costs case, the probability of cost-effectiveness falls to around 98%, and is still over 99% for the social care costs alternative. |

In their cost-effectiveness analysis, Kjerstad and Tuntland used the results from a high quality RCT that compared a multicomponent home-based reablement programme in Norway to usual domiciliary care (Tuntland et al., 2015). The intervention group received the reablement programme for 10 weeks and were followed-up at 3- and 9-months post-intervention. The primary outcome measure used in the cost-effectiveness analysis was the Canadian Occupational Performance Measure (COPM). The COPM was used to measure self-perceived activity performance and satisfaction with performance of the programme.

Costs in this evaluation were based on recorded patient contacts including the number and duration of home visits to each participant, the type of professional delivering the care and their associated wage costs. The reablement programme was found to be more cost-effective than usual domiciliary care. The incremental cost-effectiveness ratio (ICER) reported that the cost per one-point improvement in COPM-performance was 868.18 NOK lower on average for the intervention group compared to the control group. The cost per one-point improvement in the COPM-satisfaction was 666.30 NOK lower on average for the intervention group compared with the control group. The key driver of the lower costs amongst people who received the reablement programme was a lower average number of home visits in that group (88 visits compared to 158 in control group). The economic evaluation was deemed to be of high/strong quality when assessed by the JBI checklist for economic evaluations. The study did not conduct sensitivity analysis, which would further strengthen the findings.

The cost-effectiveness analysis by Glendinning et al., 2010 used the findings from a large-scale non-randomised controlled trial (NRCT) that compared five unique home care reablement services provided by local councils in England with standard home care only. Each of the reablement services differed slightly but they each consisted of a multi-professional care team with a management team overseeing their delivery. The services ran for between 6 and 8 weeks. Health-related quality of life was assessed using the EQ-5D and social care-related quality of life was assessed using the Adult Social Care Outcomes Toolkit (ASCOT). Social care service use was captured by participants and care providers, while NHS and voluntary organisation service use was self-reported only. Relevant Personal Social Services Research Unit (PSSRU) and NHS unit costs were assigned to service utilisation figures to generate costs for the cost-effectiveness analysis from both NHS and social care and wider societal perspectives. All outcomes were assessed between 9- and 12-month follow-up. At a threshold of £30,000 per EQ-5D outcome gain, there was a 99% probability that reablement was cost-effective when both health and social care costs were included and just under 100% if social care costs only were included. At a more stringent threshold of £20,000 per EQ-5D outcome gain, in the case of health and social care costs, the probability of cost-effectiveness fell to around 98% but was still over 99% when only social care costs were considered. The probability that reablement was cost-effective when reablement health care costs were between 5% and 25% higher than reported, and where health care costs for the comparison group were unchanged, were 98.1% and 89.3% respectively. This study was deemed to be of moderate quality, with some key methodological issues. EQ-5D outcome gain was not clearly defined in this study. The authors did not calculate index values or QALYs from EQ-5D responses. Further, the authors compared changes in ASCOT score to the NICE threshold. Comparison with NICE threshold should be restricted to QALY outcomes only as other outcomes have not been parameterised to the threshold. Sensitivity analysis was completed, but the issues surrounding outcome measurement influenced these results also.

The model-based cost-minimisation analysis by Bauer et al., 2018 examined whether home care reablement schemes for older people at home had the potential to reduce costs from NHS and Personal Social Services (PSS) perspectives in England. The authors developed a decision-analytic Markov model using evidence from previously published studies to run two synthetic cohorts of patients (n=1,000 in each) on which the cost-minimisation analysis was conducted. Cost-minimisation studies typically assume equivalence of outcomes, but this wasn’t stated by the authors, however the authors did acknowledge and arrive at a similar conclusion that the study by Glendinning et al., 2010 did not provide clear evidence on health outcomes from home care reablement services. Differences in service use between home care reablement services and standard home care were obtained from Lewin et al., 2013; 2014. Lewin et al., 2013 was identified in our literature search and deemed to be of low quality and at risk of bias when assessed using the CAT. Costs of home care reablement service provision were based on Glendinning et al., 2010; Lewin 2014; and national audit estimates. Costs of hospital care were generated through assigning unit costs to resource use estimates. Outcomes used in the model included relative risk of hospital admission, probabilities of hospital admission in reablement during the first year, probabilities of hospital admission in reablement during the second year, relative risk of ongoing home care and probabilities of ongoing home care for each synthetic cohort. Mean cost in the reablement group was £56,499 per person, ranging from £55,690 to £57,307 at a 95% confidence interval (CI). Mean cost in the standard care group was £58,560 (95% CI 57,800 to 59,319). The mean difference was -£2,061 (95% CI 1,933 to 2,129). In both groups, most of the total costs (over 80%) were social care related: If reablement was counted as a cost to local authorities, mean costs to this sector were £46,894 in the reablement group and £48,887 in the standard care group. On the same basis, 97% (£1,993) of the net benefit was due to reductions in costs to local authorities, and only 3% (£68) was due to reductions in costs to the NHS. Costs in both reablement and standard care groups decreased with higher starting ages of cohorts, mean differences increased to - £3,244 (starting age 75) and -£3,844 (starting age 85). The decrease in costs was explained by the overall shorter period in which costs occurred, and the increase in mean differences reflected the relatively greater impact on hospital admission rates for older age groups (who have higher risks of hospital admission). The probability that reablement was cost-reducing increased to 99.9% (for starting age 75) and 100% (for starting age 85). The model parameters utilised evidence from primary studies deemed to be of poor to moderate quality, which influenced the quality of this study when assessed by the JBI checklist (moderate quality). Further, in the absence of high quality evidence from England, the model used Australian data. The authors noted expert opinion found both countries’ reablement systems to be similar, but the use of such data influenced generalisability to an English population.

#### 2.3.4 Bottom line results for the cost-effectiveness of reablement interventions

Two of the economic evaluations suggested at-home reablement services could be cost-effective when compared to standard at-home care. The quality of this evidence was mixed however, with one study being of high quality and the other being of moderate quality, with key methodological challenges concerning outcome measurement. The cost-minimisation analysis indicated that at-home reablement interventions can be less costly when compared to standard at-home care. This study used previously published evidence from Australia to inform model parameters which may influence generalisability to an English/UK care system.

### 2.4 Service-level costs and implementation outcomes of reablement interventions

#### 2.4.1 Costs

Five of the included studies reported on service-level costs as one of their outcomes (Glendinning et al., 2010; Lewin et al., 2013b; Lewin et al., 2014; Kjerstad and Tutland, 2016; Beresford et al., 2019).

In England, a medium quality non-randomised controlled trial compared NHS health service use costs and social care service use costs between a reablement service group and a usual domiciliary care control group over a 12-month period (Glendinning et al., 2010).

During the first 8 weeks, reablement was associated with significantly higher NHS and social care service costs due to the cost of the reablement services themselves. At 10 months, NHS costs were similar between groups but the control group incurred significantly higher (p<0.0.001) mean social care service use costs compared to the reablement group including in-house home care (£270 reablement group vs. £590 control), independent home care (£450 reablement group vs. £1,660 control group), day care (£0 reablement group vs. £60 control) and meals on wheels (£60 reablement group vs £70 control). Overall, at 12 months, there was no significant difference between groups in total healthcare costs (Glendinning et al., 2010).

A medium quality UCBA study conducted in England reported an estimated cost of reablement per case of £1,700 (2016 prices) (Beresford et al., 2019). The study reported that public sector health and social care costs were highest before the reablement, with a large reduction in the cost of hospital stays following the implementation of reablement. Informal care time was a main driver of cost before and during implementation (Beresford et al., 2019).

In a low quality RCT conducted in Australia, results reported lower total home care costs for home independence program (HIP) clients (AU$5570) compared to usual domiciliary care services (AU$8541) (Lewin et al., 2013b). Over the 24-month follow-up period, the mean home care and healthcare costs per client were AU$2869 lower for the HIP group compared to usual domiciliary care services (Lewin et al., 2014). A separate study, a medium quality retrospective cohort study compared the costs between two reablement services (HIP and personal independence program (PEP)) with usual domiciliary care services (Lewin et al., 2013b). Both reablement groups (HIP and PEP) were less likely to use home care services compared with usual domiciliary care, which equated to an estimated median cost saving of AU $12,500 per person over 5 years (Lewin et al., 2013b).

A high quality cost-effectiveness analysis used the results from a high quality RCT that compared a multicomponent home-based reablement programme in Norway to usual domiciliary care (Tuntland et al., 2015). Kjerstad and Tutland, 2016 calculated costs of delivering the multicomponent home-based reablement programme investigated in the RCT. Over the ten-week programme, aggregate delivery costs were approximately 12,000 NOK lower for the intervention group than control. At 3-month follow-up, mean costs per participant was approximately 1,134 NOK lower for the intervention group compared with the control (Kjerstad and Tutland, 2016).

#### 2.4.2 Implementation

Three included studies reported on implementation outcomes: Beresford et al., (2019), Whitehead et al., (2016) and Glendinning et al., (2010). The prospective cohort study by Beresford et al., (2019) reported on the appropriateness of the models of English reablement interventions. The study mapped and created a typology of existing statutorily funded adult social care reablement service models in England, before evaluating three distinct models. Data was captured from staff and participants through questionnaires and interview at discharge and 6-month post-discharge. The results indicated that staff advocated reablement interventions over usual domiciliary care. Service users reported the acceptability of the intervention and cited six factors had an impact on outcomes (the service user–worker relationship; the workers’ reablement skills; the service user’s confidence in the worker; the duration of home visits; the service user’s willingness to accept support; and the service user being able to review progress). Beresford et al. reported acceptability issues of implementation, and that staff frequently reported that service users and family members had a poor understanding of reablement, including confusion about its difference from usual domiciliary care, and this acted as a barrier to engagement. The study also identified concerns with monitoring and review processes and with the sustainability of the intervention as staff caseload volume had increased.

Whitehead et al., (2016) conducted a feasibility RCT that compared a model of reablement that was led and delivered by occupational therapists to a model of reablement that was led and delivered by social care workers (control group). In the feasibility outcomes, the authors reported that, overall, it was possible to deliver the intervention as planned and that it was delivered consistently with the protocol. Each participant received an average of 10 hours of occupational therapy time: 4 hours of direct contact, 3 hours of administration and liaison, and 3 hours of travel time. All participants were able to set one or more ADL-related goals. The intervention acceptability was evaluated using a questionnaire and semi structured interviews with five participants in the intervention group. The questionnaire and interviews revealed a high level of satisfaction with the intervention, and participants reported that they believed the intervention helped them to increase their ability to manage ADL. However, this was a feasibility RCT and further research is needed to ascertain whether intervention delivery could be standardised across sites.

Glendinning et al., (2010) reported on the implementation of five English reablement services through the non-randomised controlled trial element of their study. The study captured qualitative and quantitative data from service users, carers and staff through validated and bespoke questionnaires, interviews and focus groups. Service users and their carers reported positive experiences of reablement, particularly the good relationships they were able to develop with the team of reablement workers. The reassurance provided by visits from reablement workers was particularly important for users living alone. Factors contributing to the successful delivery of five reablement services included not having highly selective intake policies, unsustainable levels of resourcing, or particularly highly motivated staff (often features of new or pilot services which are likely to enhance positive findings and outcomes). Thus, the authors concluded that the promotion of home care reablement was well-founded. However, the authors also noted that a number of service users and carers were disappointed that home care reablement had not been able to address two areas of importance to them, in this instance preparing meals and rebuilding confidence and improving mobility to walk outside the home.

## 3. SUMMARY OF THE FINDINGS OF A QUALITATIVE EVIDENCE SYNTHESIS EXPLORING OLDER PEOPLE AND THEIR CARERS’ EXPERIENCES AND PERCEPTIONS REABLEMENT SERVICES

Our rapid review focused on the assessment of the clinical and cost-effectiveness of reablement service and was limited to comparative studies. However, we also acknowledge the value of qualitative research exploring older people and their carers’ experiences and perceptions of reablement, which is likely to provide important information on the barriers and enables for implementing the service. This section summarises the findings of a systematic review and qualitative evidence synthesis that was identified during the preliminary searches for this rapid review.

A systematic review of qualitative studies investigated the experiences and perceptions of older individuals and their carers (formal and informal) regarding the reablement model used in community aged care (Mulquiny and Oakman, 2022). The review encompassed 19 qualitative studies which highlighted the potential of reablement to foster independence, challenges associated with goal setting, and the necessity for enhanced engagement strategies. A prominent finding across the studies was that both older people and their carers viewed goals as crucial measures and motivators during the reablement process, with goal attainment positively influencing their experiences. Findings across studies demonstrated reablement improved the health and well-being of older individuals. However, the success of these improvements was closely tied to the level of engagement and understanding of the reablement process by the older individuals. Poor engagement in reablement services among older people was attributed to their limited understanding of their role in the process. Expectations often aligned with traditional aged care services, which contrasted with the more hands-off and rehabilitative approach of reablement. Enhanced collaboration in setting reablement goals between practitioners, older individuals, and their carers was found to be essential in improving understanding and engagement. While older individuals appreciated the principles of reablement, such as supporting people to remain independent within their usual place of residence, the short duration of reablement services were considered insufficient for the level of disability experienced by some individuals, who expressed a need for ongoing interventions. Additionally, older people felt that their reablement goals often overlooked their need for social connectedness, with leisure and social activities rarely included in the goals of reablement programmes (Mulquiny and Oakman, 2022).

## 4. DISCUSSION

### 4.1 Summary of the findings

This rapid review identified a significant amount of international evidence that reablement led by multi-professional teams that includes allied health professionals, or an allied health professional such as an occupational therapist is effective in improving person-related outcomes and reducing the need for long term care needs. Twelve studies evaluated reablement models led by multi-professional teams that included allied health professionals, while five studies specifically examined AHP-led models. Only one study involved no AHPs, evaluating a reablement model led by a nurse case manager. In response to the rapid review question, there was evidence to demonstrate that reablement interventions were effective for improving independence in terms of increasing mobility and activities of daily living outcomes. Other outcomes relating to clients’ health reported in the studies identified in this rapid review included quality of life, falls outcomes, grip strength, sense of coherence, mortality and social support. The rapid review findings suggest that reablement interventions were effective in improving quality of life and may have been effective in improving falls outcomes. The review also identified that reablement may have been effective in reducing the risk of mortality and improving clients’ coping in terms of sense of coherence. The review did not identify evidence to suggest that reablement was effective for improving grip strength or increasing clients’ social support.

Another aim of this rapid review was to assess whether reablement was effective in reducing the need for long term care. Based on the available evidence identified in this rapid review, reablement was found to reduce the need for long term home care services. There was also evidence that reablement interventions were effective in reducing residential care admissions. In terms of other service-level outcomes, there were contradictory findings on the effectiveness of reablement to reduce hospital admissions and inconsistent findings on the effectiveness of reablement to reduce emergency department visits. One study found that reablement was effective in reducing the number of outpatient treatments compared with usual domiciliary care. The rapid review also found contradictory findings on the effectiveness of reablement to reduce community care service use and social care service use.

This review also identified a medium-quality feasibility RCT conducted in England that compared a model of reablement that was led and delivered by occupational therapists to a standard model led and delivered by social care workers under the supervision of a reablement care team leader (Whitehead et al., 2016). The study found greater improvements in person-related outcomes—including activities of daily living, health-related and social care-related quality of life, and a reduction in falls in the occupational therapy reablement group. Although the sample size was small (N=30) and findings were not statistically significant, the occupational therapy-led model showed promising trends. At the service level, participants in the occupational therapy-led reablement group were less likely to use community and social care services, including GP visits and community support such as meals at home, suggesting potential efficiency benefits compared to the social care worker–led model.

In terms of the cost-effectiveness of reablement, this rapid review identified three economic evaluations. Two of the economic evaluations (one set in England, one set in Norway) found that at-home reablement services were cost-effective when compared to standard at-home care. The third economic evaluation, a cost-minimisation analysis (set in England but incorporated Australian data), reported that reablement was cost-effective compared to usual domiciliary care when the clinical effectiveness is assumed to be equal between the intervention and usual domiciliary care groups, due to lower health and social care resource use costs in the reablement intervention group.

Some of the studies included in this rapid review also reported on other relevant outcomes, such as costs and implementation outcomes. Five studies reported on service-level costs. One UK study reported similar total healthcare costs for people who received a reablement intervention compared to people who received usual domiciliary care services at 10-month and 12-month study follow-up periods; however, the costs of social care services were significantly higher in the usual domiciliary care group compared with the reablement group at the 10-month follow-up. Another UK study reported that reablement was associated with a large reduction in hospital stay costs. Two studies reported service-level costs of reablement in Australia. Both studies reported lower home care service use costs for reablement compared with usual domiciliary care. One Norwegian study reported aggregate delivery costs were lower for the intervention group than control, and at 3-month follow-up, mean costs per participant was lower for the intervention group than control.

Three studies reported on implementation outcomes in terms of the acceptability of reablement interventions. There were mixed findings in relation to the acceptability of reablement as noted by the experiences of service users, carers and staff. Some of the reported barriers to the acceptability of reablement included a poor understanding of reablement among service users, not addressing goals that were important to the service user, concerns with monitoring and review processes, and concerns around the sustainability of the intervention as staff caseload volume had increased. There was also some evidence that reablement interventions were feasible to deliver.

### 4.2 Strengths and limitations of the available evidence

All evidence included in this review was from academic papers, with the exception of Glendinning et al., 2010 which was not published in an academic journal and did not go through peer review process. All included studies evaluated a form of reablement care. When assessed against key components of reablement (see Table 1), most interventions from included studies satisfied all five criterion [1) participants must have had an identified need for formal care and support or be at risk of functional decline 2) intervention must have been time-limited (up to 12 weeks) and intensive (e.g., multiple home visits) 3) intervention must have been delivered in the person’s own home, and provided by an interdisciplinary team 4) intervention must have been focused on maximising independence and 5) intervention must have been person-centred and goal-directed]. Of criterion not satisfied, we cannot be certain that four studies provided time-bound reablement as they did not report on duration. Most of the studies reported on reablement that appeared to be provided by healthcare providers, with fewer studies reporting on reablement that was hosted within or delivered by local authorities. It is important to note that occupational therapists working within local authorities currently face significant challenges in engaging in research activities.

The evidence comprehensively covered both person-related and service-level outcomes, with evidence of key outcomes in each category identified. Only three economic evaluations were identified and had some methodological flaws, highlighting an evidence gap. Study designs were variable, with four studies using full RCTs. Some studies employed a single cohort design, which did not allow for comparisons of the effectiveness of reablement services to be made against a comparator.

Most of the identified evidence was rated to be of moderate quality. Only one study was rated as high quality, the RCT by Tuntland et al., 2015. The differences in quality highlights a limitation of the available evidence, affecting the overall strength and reliability of the review’s conclusions. Further research utilising strong study designs and robust methodologies are required to provide more definitive evidence of the effectiveness and cost-effectiveness of reablement services across the domains of person-centred and service-level outcomes.

### 4.3 Strengths and limitations of this Rapid Review

This rapid review systematically searched five bibliographic databases and followed a rigorous and comprehensive search strategy that was co-produced with stakeholder and information scientist input. All results were dual-screened for relevance by two reviewers at title and abstract and full-text stages to ensure consistency and accuracy. Any conflicts were resolved by a third reviewer. Inclusion and exclusion criteria were co-produced by expert stakeholders to ensure suitable and appropriate study selection. This review reported not only on effectiveness and cost-effectiveness, but also on wider outcomes such as implementation.

Included studies were critically appraised individually as well as a quality appraisal of the evidence base for included outcomes in this review. A generic critical appraisal tool was used for all studies of effectiveness, which facilitated the comparison of methodological quality across different study designs while also accounting for study design during the appraisal process.

Due to the differences in terminology used to define reablement across countries, we cannot be certain we have not missed other relevant search terms to identify studies of reablement. Our search strategy was defined with input from the stakeholder team.

### 4.4 Implications for policy and practice

- There is international evidence that reablement (led by a multi-professional team that includes allied health professionals, or an allied health professional such as an occupational therapist) is effective in improving mobility including activities of daily living, quality of life and falls outcomes. There is also evidence that demonstrated reablement can reduce the need for long term care in terms of the use of home care services and admissions to residential care.
- While not all reablement services currently include allied health professionals (AHPs), the evidence from this review makes a strong case for their inclusion in Wales. AHP-led reablement has been shown to improve person-centred outcomes and reduce the need for long-term care, supporting the case for targeted investment in AHP roles within reablement services.
- From a UK and Norwegian context, three economic evaluations reported that reablement was a cost-effective alternative to usual domiciliary care services.
- Given the high costs associated with ongoing care needs for people at risk of frailty, our findings make the case for investing in time-limited reablement interventions in Wales.
- Wales currently has different models of reablement provision, guided by the All Wales Rehabilitation Framework (2022). This review makes the case for more focus on reablement services to provide better outcomes for people, with appropriate resource, support and prioritisation of allied health professional resource in order to achieve optimal systems impact.
- Future recommendations for policy and practice should also consider findings relating to the acceptability of reablement to service users and staff, and also the feasibility of implementing these interventions in practice.

### 4.5 Implications for future research

- There is a clear need for more UK-based studies on reablement, as this review identified only three from the UK, with the remainder drawn from international evidence. While international research can offer valuable insights—particularly where service models align—it is essential to consider the unique context of Wales, where reablement is often hosted within or delivered by local authorities. Most of the included studies identified in this review reported on interventions that appeared to be delivered by the healthcare sector.
- There is a need for high quality evidence on the clinical and cost-effectiveness of reablement services. The majority of included studies were deemed to be of moderate quality, limiting the certainty of the review findings.
- The three identified economic evaluations had some methodological flaws that limited the certainty of review findings, evidencing a need for future economic evaluations on the topic.
- Consider the challenges faced by professionals in undertaking research outside the field of Health, as reablement is often hosted within the remit of local authorities and could involve social care practitioners, such as occupational therapists. Future research should consider sub-group analyses to ensure equitable provision of reablement services across the whole of Wales for people from all socioeconomic backgrounds and ethnicities.

### 4.6 Economic considerations*

- Reablement programmes may provide cost savings to commissioners and the health and social care systems through prevention of or reduced length of hospital admission, reductions in hospital readmission and preventing or reducing domiciliary and residential care demand (Deloitte, 2012).
- Frailty has a sizeable impact on healthcare resource use in the UK. Total additional costs of frailty-related healthcare resource use are £8 billion** per year. Healthcare resource use costs increase with severity of frailty. Additional per person costs from an NHS perspective were £773**, £1,666** and £2,904** for mild, moderate and severe frailty respectively (Han et al., 2019).

*\*This section has been completed by the Centre for Health Economics & Medicines Evaluation (CHEME), Bangor University*

*\*\*Inflated to April 2025 prices using Bank of England inflation calculator* https://www.bankofengland.co.uk/monetary-policy/inflation/inflation-calculator

## Abbreviations

Acronym: Full Description
ADL: Activities of Daily Living
AHP: Allied Health Professional
ASCOT: Adult Social Care Outcomes Toolkit
AU $: Australian Dollar
CAT: Critical Appraisal Toolkit
CEA: Cost-Effectiveness Analysis
CHEME: Centre for Health Economics & Medicines Evaluation
CI: Confidence Interval
CMA: Cost-Minimisation Analysis
COOP/Wonka (WONCA): The Dartmouth Co-op Functional Health Assessment Charts/WONCA (World Organization of Family Doctors)
COPM: Canadian Occupational Performance Measure
EQ-5D: EuroQol-5 Dimension
FIRST: Flexible Integrated Responsive Support Team
GP: General Practitioner
HRQoL: Health-related quality of life
HIP: Home Independence Program
HIPC: Home Independence Program coordinator
HACC: Home and Community Care
IADL: Instrumental Activities of Daily Living
ICER: Incremental Cost-Effectiveness Ratio
ITT: Intention To Treat
JBI: Joanna Briggs Institute
MDT: Multidisciplinary Team
NEADL: Nottingham Extended Activities of Daily Living Scale
NHS: National Health Service
NOK: Norwegian Kroner
NRCT: Non-Randomised Controlled Trial
OECD: Organisation for Economic Co-operation and Development
OTHERS: Occupational Therapy in HomEcare Re-ablement Services
PRISMA: Preferred Reporting Items for Systematic reviews and Meta-Analyses
PSSRU: Personal Social Services Research Unit
PEP: Personal Enablement Program
QoL: Quality of life
RCT: Randomised Controlled Trial
RR: Rapid Review
SF-36: Short Form 36-item survey
SOC-13: Sense of Coherence Questionnaire
SPPB: Short Physical Performance Measure Battery
STRC: Short-Term Restorative Care Program
TUG: Timed Up and Go
UCBA: Uncontrolled Before-After
UK: United Kingdom
US: United States

## Glossary

**Allied health professional-led (AHP-led):** describes an intervention that is led, coordinated, or managed by *an* allied health professional (AHP), such as a physiotherapist, occupational therapist, or speech and language therapist, even if the intervention itself is delivered by other staff (e.g., support workers or care assistants). This is distinct from a multi-professional team approach, which involves active collaboration and shared decision-making among professionals from multiple disciplines (that could include AHPs).

**Economic evaluation:** an assessment of the costs and effects of alternate healthcare interventions. The aim of an economic evaluation is to help decision makers maximise the level of health benefits relative to the finite resources available.

**Coherence**: how people view life and their capacity to respond to stressful situations. It consists of three core components: Comprehensibility, Manageability, and Meaningfulness. Sense of coherence is measured using the Sense of Coherence Questionnaire.

**Cohort study:** a type of observational study in which two or more groups of people—one with a particular exposure, treatment, or characteristic (e.g., smokers), and one without (e.g., non-smokers)—are followed over time to assess differences in the occurrence of a specific outcome. The inclusion of a comparison or “control” group allows researchers to examine the effect of the exposure by contrasting outcomes between the groups. Unlike randomised trials, participants are not randomly assigned, but observed in their natural exposure settings.

**Consistency of results:** consistent results report findings from studies that found similar results, inconsistent results report some variation in results but overall trend related to the effect is clear, contradictory results report varying results with no clear overall trend related to the effect.

**Cost-effectiveness analysis:** a type of economic evaluation whereby costs are compared with a treatment’s common therapeutic goal, expressed in terms of one main outcome measured in natural units (e.g., improvement in blood pressure or cholesterol level). These outcomes are typically condition-specific, meaning comparison within conditions is possible, but difficulty arises in comparing between conditions.

**Cost-minimisation analysis:** is a form economic evaluation which compares the cost of alternative interventions or services that achieve the same outcomes. In the analysis, if the overall cost of new intervention is lower than the comparator intervention, the intervention of interest is considered cost-effective. This method aims to identify the least costly option.

**Directness of evidence:** direct evidence comes from studies that specifically researched the association of interest, whereas extrapolation is inferences drawn from studies that researched a different but related research question or researched the same question but in an artificial setting.

**Health and social care resource use or utilisation:** refers to the use of healthcare resources by end users (patients) and intervention deliverers. This can take the form of contacts with health professionals across health and social care, medicines or healthcare consumables used. In economic evaluations, resource use is collected, and costs are assigned to them to identify the costs an intervention places on the healthcare system.

**Intervention deliverer**: personnel who deliver the intervention to participants.

**Intervention leads:** individuals responsible for overseeing the design, coordination, and strategic implementation of an intervention. They guide the overall direction and support those delivering it, but are not necessarily involved in direct delivery to participants.

**Model-based economic evaluation:** Model-based economic evaluations are often used especially when long term evidence is important but not available. It can extrapolate findings beyond the trial-based follow-up period (time horizon). Model-based economic evaluations use past knowledge and patterns to understand the performance of new and existing treatments. This can be performed alongside empirical studies or as standalone studies. The model can be used to synthesise evidence across multiple trials for decision making. The model types widely used are simple decision trees, Markov cohort, and Markov microsimulation models.

**Multi-professional team/ multi-disciplinary team:** a group of healthcare professionals from different disciplines who come together to discuss cases, share expertise, and jointly develop coordinated management plans.

**Multi-professional workforce:** the broader composition of personnel from various professional backgrounds who are employed within a service or system. Unlike a multi-professional team, this term refers to the overall staffing structure, not necessarily implying active collaboration or joint working, but rather the presence of diverse professional roles within the workforce.

**Non-randomised controlled trial:** is a study design that participants are not randomly assigned into an experimental group or a control group. Participants may choose which group they want to be in, or they may be assigned to the groups by the researchers.

**Overall rating of the body of evidence:** considers the strength of the study designs, quality of the studies, directness of evidence and consistency of results across the studies to draw a conclusion for each outcome. Please see section 6.8 of this report for further information about the grading criteria for rating the overall body of evidence.

**Randomised controlled trial:** a study in which a number of similar people are randomly assigned to 2 (or more) groups to test a specific drug, treatment or other intervention. One group (the intervention group) has the intervention being tested, the other (the comparison or control group) has an alternative intervention, a dummy intervention (placebo) or no intervention at all. The groups are followed up at set time periods to see how effective the experimental intervention was. Outcomes are measured at specific times and any difference in response between the groups is assessed statistically. Randomised controlled trials are the highest standard of research trials as their design helps to reduce biases that may impact the findings.

**Reablement:** referred to by NICE (2017) as a community-based intervention that aims to increase a service user’s independence by helping them recover lost skills and confidence. The All Wales Rehabilitation Framework (2022) refers to reablement as an enabling approach with services that provide rehabilitation for all people with physical or mental disabilities. It helps them adapt to their condition by learning or re-learning the skills needed to function in everyday life. The focus is on promoting and optimising functional independence by practicing activities, rather than interventions aimed at improving underlying impairments.

**Sample size range:** presents the highest and lowest sample size across the studies for a given outcome of interest.

**Statistical significance:** a statistically significant result is one that is deemed to be down to a true effect rather than random chance. It is a way to determine if a relationship between variables is caused by something other than chance.

**Strength of study designs:** each analytical study design has particular strengths and limitations and are assigned a strength of study design available from the critical appraisal tool guidance.

**Step down reablement:** support after hospital discharge.

**Step up reablement:** proactive support for individuals at risk of deteriorating, aiming to prevent hospital admissions, improve health outcomes, and reduce long-term care costs

**Quality of studies:** each analytical study is given a quality rating based on the scoring on the critical appraisal tool quality assessment for individual studies.

**Trial-based economic evaluation:** an economic evaluation of the costs and effects of healthcare interventions that are being studied as part of a clinical trial. The primary aim of clinical trials is typically a measurement of clinical effect of administering an intervention (e.g., change in blood pressure). These clinical effects of the intervention are then assessed together with the costs of the intervention in the economic evaluation.

**Uncontrolled before-after study:** is a study that evaluates the outcome of an intervention before and after implementation of the desired intervention. This method is known as an uncontrolled before-after study. It aims to assess whether the intervention has an effect on the measured outcome. The term ‘uncontrolled’ is used to distinguish this design from a controlled before-after study. In an uncontrolled before-and-after study, the effect of the intervention is not compared to a control group that did not receive the intervention.

## Data Availability

All data produced in the present study are available upon reasonable request to the authors

## 6 RAPID REVIEW METHODS

### 6.1 Eligibility criteria

An extended version of the Population, Intervention (exposure), Comparison, Outcomes (PICO) framework table is presented below detailing the inclusion and exclusion criteria of this review.

**Table 8:**
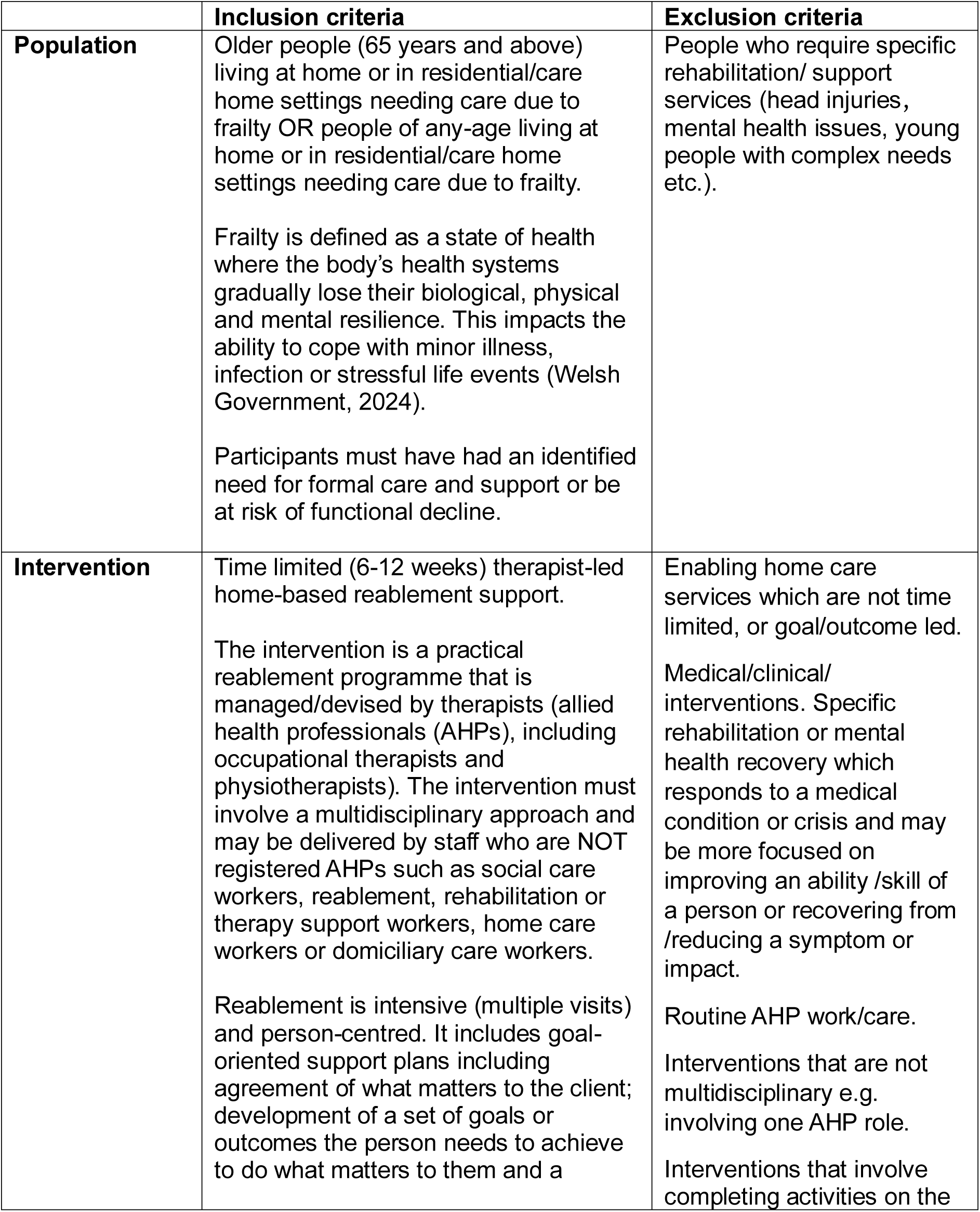
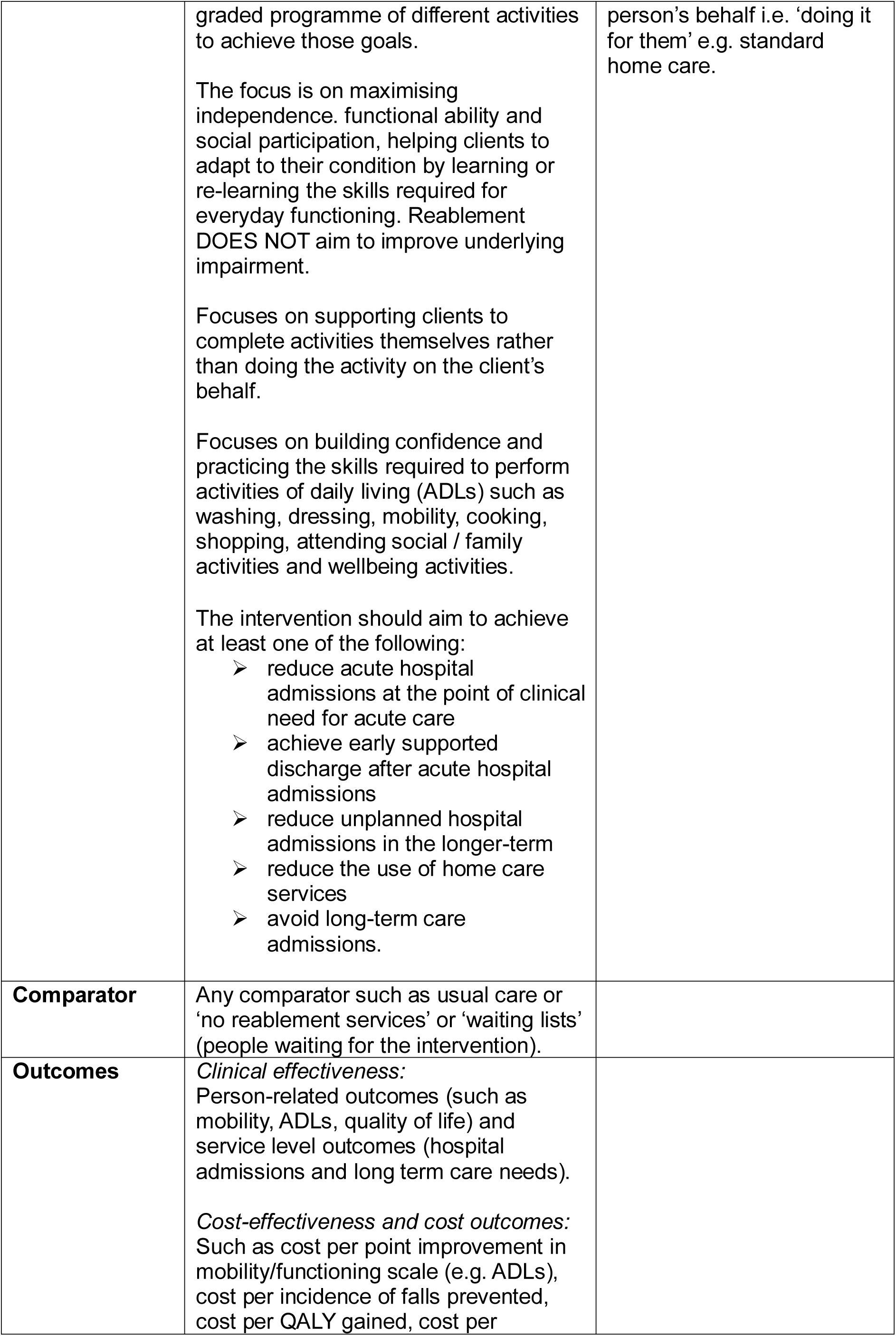
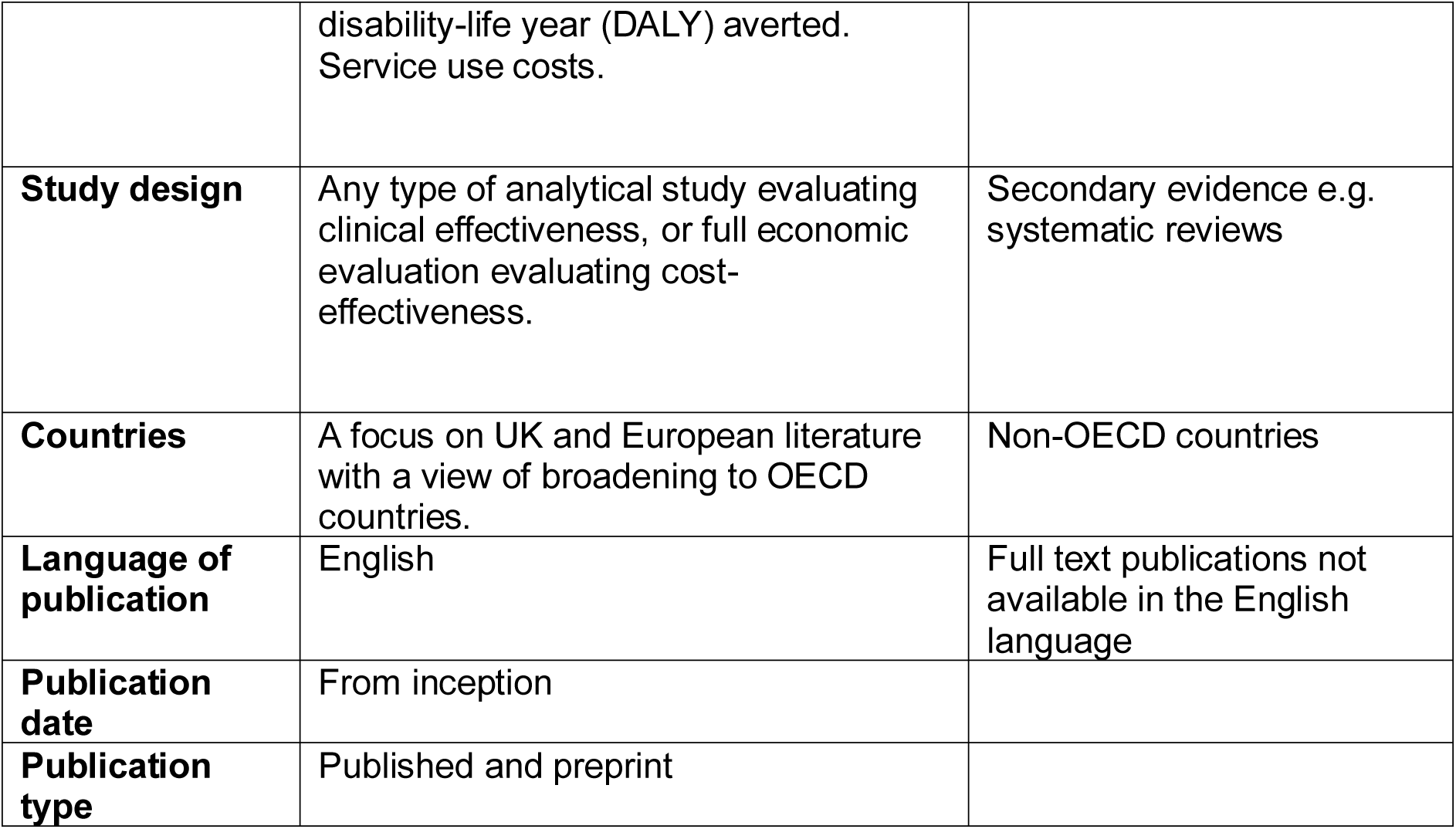
Eligibility criteria.

### 6.2 Literature search

A comprehensive search of bibliographic databases was conducted for publications from database inception to January 2025. The following databases were searched:

- On the OVID platform: Medline and EMBASE
- On the EBSCO platform: CINAHL
- Cochrane: CENTRAL & CDSR

### 6.3 Study selection process

Four reviewers independently screened titles and abstracts using the Covidence review management software. Two reviewers independently screened 100% of the full text articles. Following the independent full text screening stage, discrepancies were resolved through discussion with the review team to come to an agreement on the final inclusions.

### 6.4 Data extraction

Data extraction was based on the outlined eligibility criteria. For the economic evaluation studies, the review team extracted data on study country, type of intervention, study design, sample size, length of follow-up, type of economic evaluation, perspective of analysis, currency and cost year, details of discounting and sensitivity analysis, main costs and outcomes measures, and main health economics findings. For the remaining primary studies, the review team extracted data of study country, type of intervention, study design, sample size, participants and settings, intervention and comparator/control, outcomes that are relevant to the review question. Resource use, costs and implementation were also extracted if applicable to the study.

### 6.5 Study design classification

Studies were classified using the algorithm supplemented in the Critical Appraisal Toolkit (CAT) designed by the Public Health Agency of Canada (Public Health Agency of Canada, 2014). An evidence summary table was utilised in order to extract further detail surrounding the outcomes of the intervention and the results specific to each individual outcome.

### 6.6 Quality appraisal

The Critical Appraisal Toolkit (CAT) for assessing multiple types of evidence will be used to critically appraise the included studies (Moralejo et al., 2017).

### 6.7 Synthesis

Quantitative data from each study was narratively synthesised and analysed. Due to the heterogeneity of intervention type, study outcomes and outcome measures, it was not possible to conduct a meta-analysis. Following detailed extraction of interventions, outcomes, and outcome measures, a thematic approach was undertaken to synthesis findings.

The review findings were presented under the following outcome categories:

1. Person-related outcomes

a. Mobility including ADLs
b. QoL
c. Falls outcomes
d. Other person-related outcomes
2. Service-level outcomes

a. Admissions to residential care
b. Long-term home care needs
c. Hospital admissions
d. Other health and social care resource use
3. Cost-effectiveness
4. Other relevant outcomes

a. Costs
b. Implementation
c. Summary of findings from a previously identified qualitative evidence synthesis

Evidence of any outcomes relating to acceptability, feasibility and implementation reported in the identified studies were summarised to highlight implementation issues and recommendations for policy and practice.

### 6.8 Assessment of body of evidence

The CAT was used to assess the body of evidence identified in this rapid review (Moralejo et al., 2017). This toolkit provides three main appraisal tools for analytical studies, descriptive studies, and literature reviews. The CAT facilitates a comprehensive assessment of the body of evidence for different study designs and was used to produce an evidence summary table detailing information on the relevant methods and outcome measures, results and conclusions from each study. The CAT was used to rank each study as ‘strong’, ‘moderate’ or ‘weak’ which is based on a number of variables such as the type of study design, assessment of the study population and sampling method, internal validity, control of confounding variables, ethics, control of analysis, assessment of applicability, and overall conclusions (Moralejo et al., 2017).

To assess the overall body evidence for each outcome, five outcomes suggested by Moralejo et al. (2017) were taking into account, including the CAT quality decisions; the number of studies evaluating the same population; directedness of the evidence; and consistency of results. Based on these items, a rating for the overall body of evidence for each outcome can be described as strong, moderate, or weak (Moralejo et al. 2017). Grading criteria is presented below.

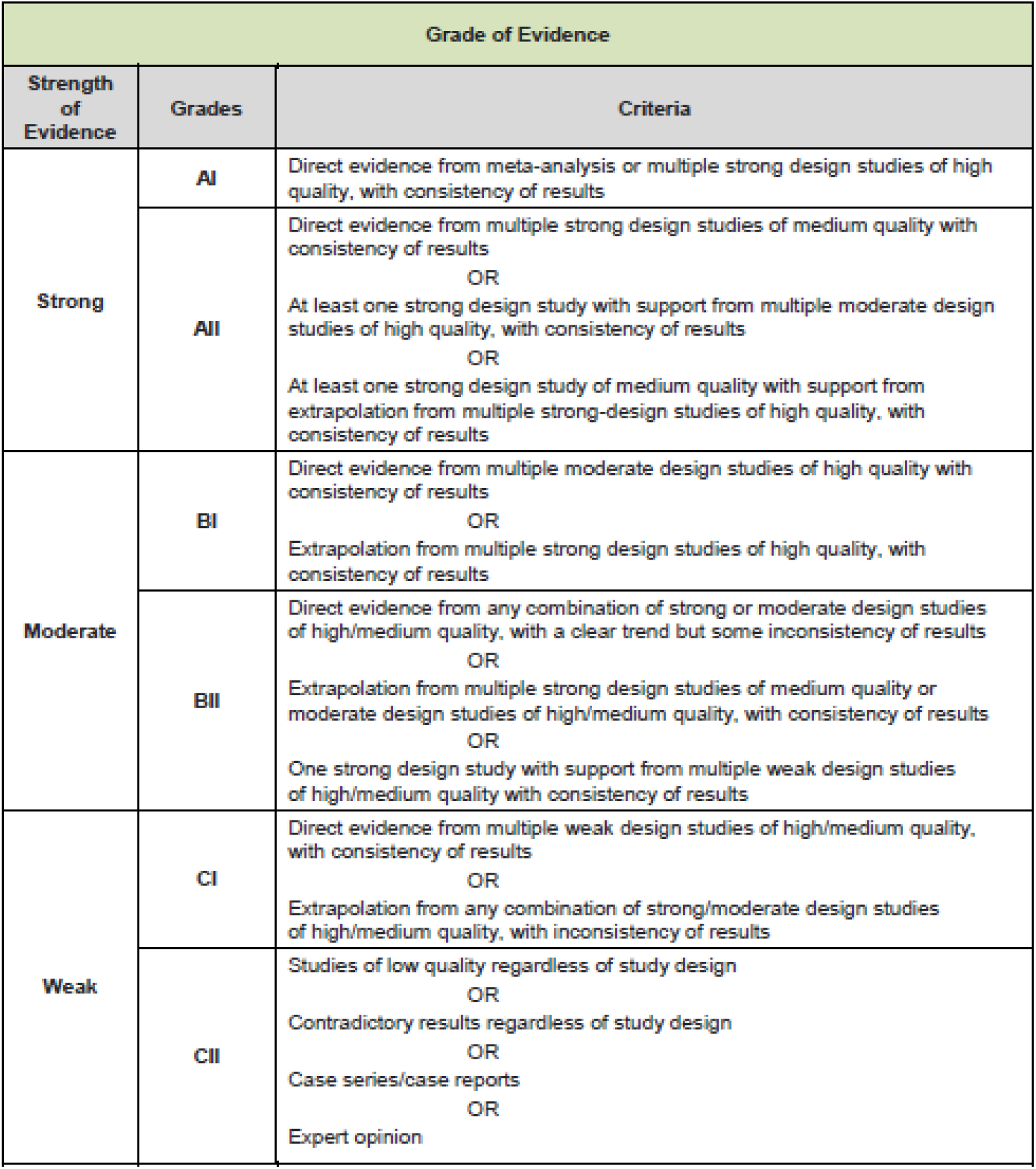

## EVIDENCE

### 7.1 Search results and study selection

Following the removal of duplicates, the search identified 4,316 records. Full texts (n=46) were reviewed, and eighteen studies were included in this rapid review: studies of clinical effectiveness (n=15), economic evaluations (n=2), study including both an evaluation of clinical effectiveness and an economic evaluation (n=1). Please see Section 9.3 of this report for further information on the reasons for the exclusion of records during the full text screening stage.

### 7.2 Data extraction

**Table 9.**
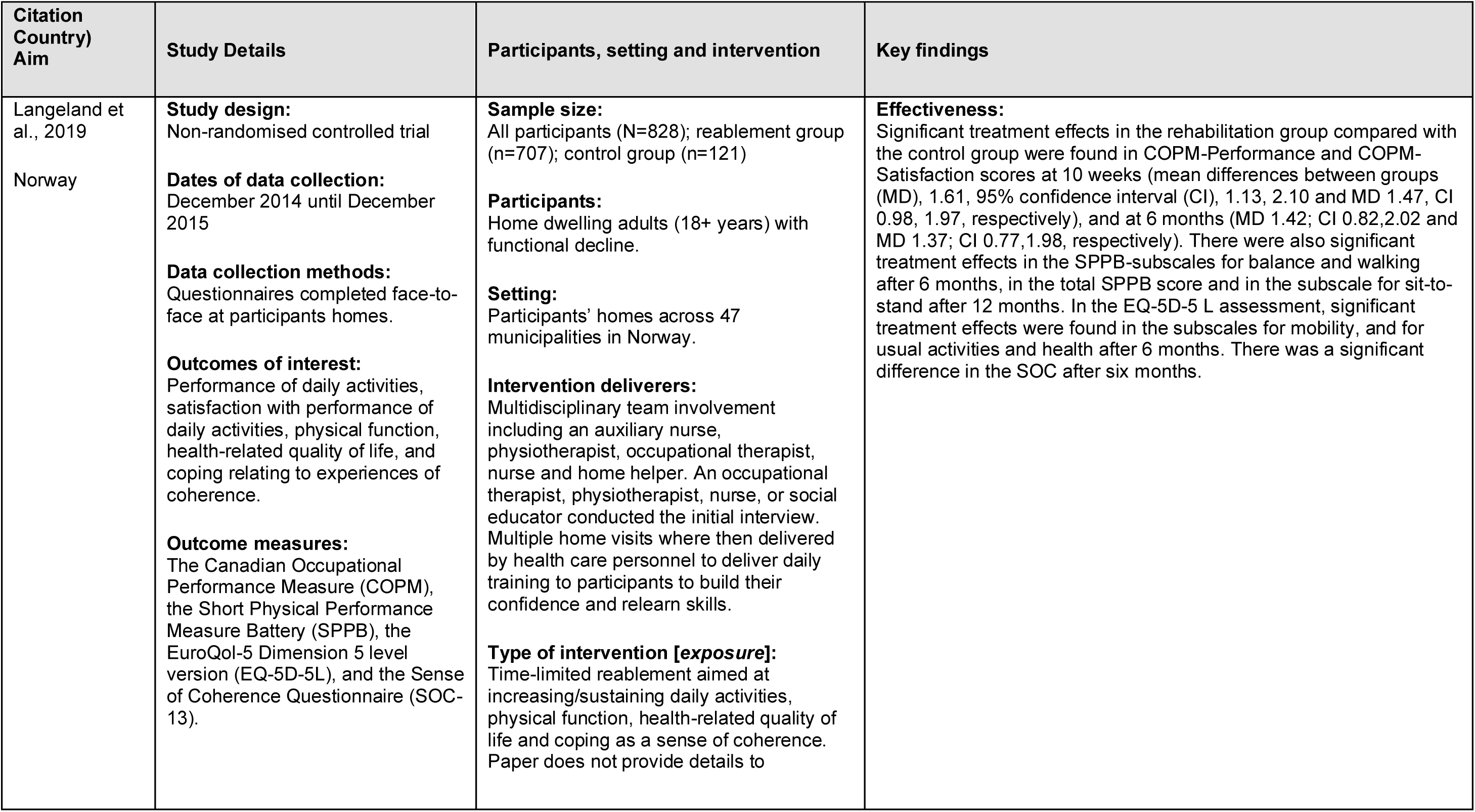
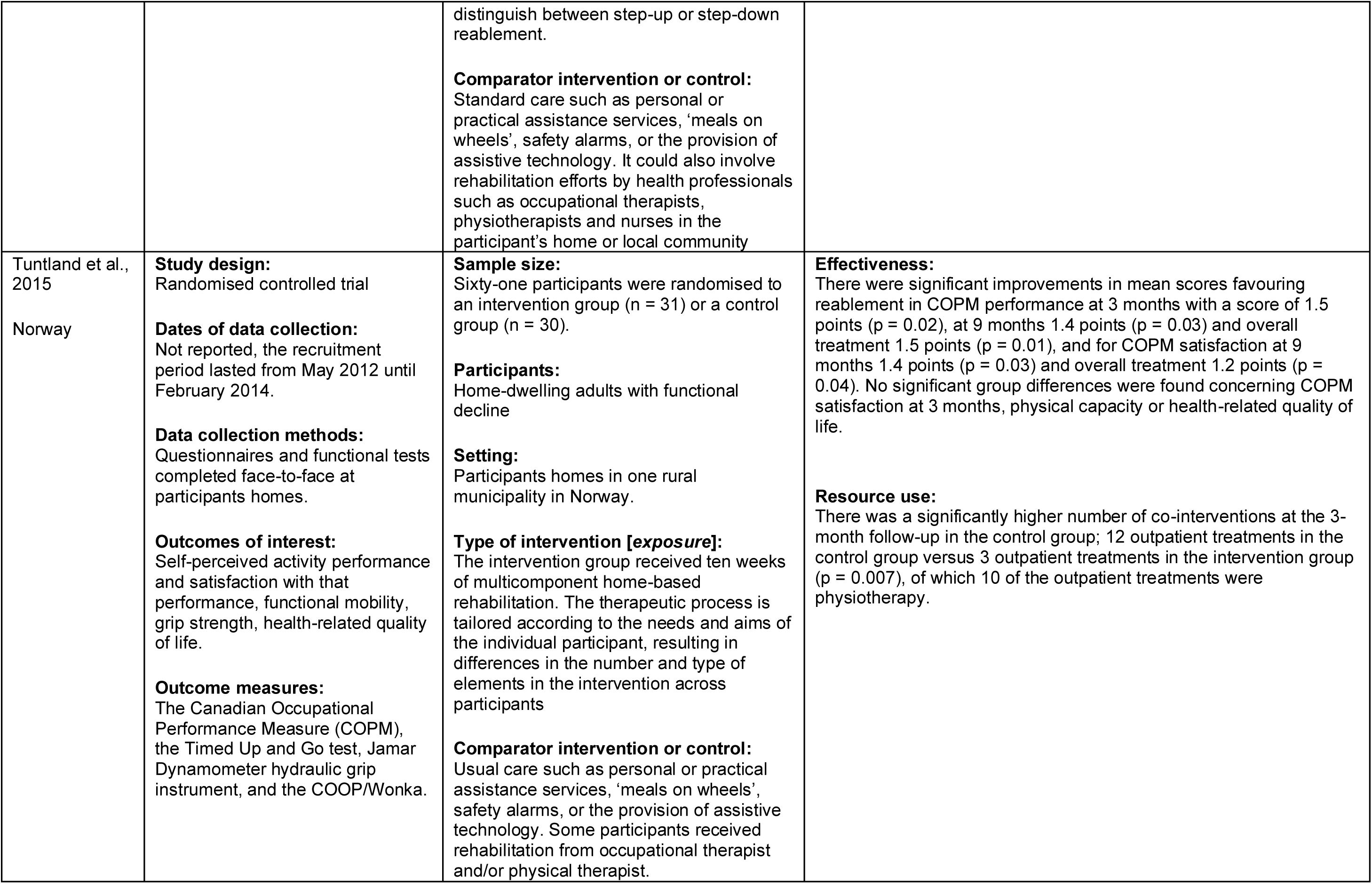
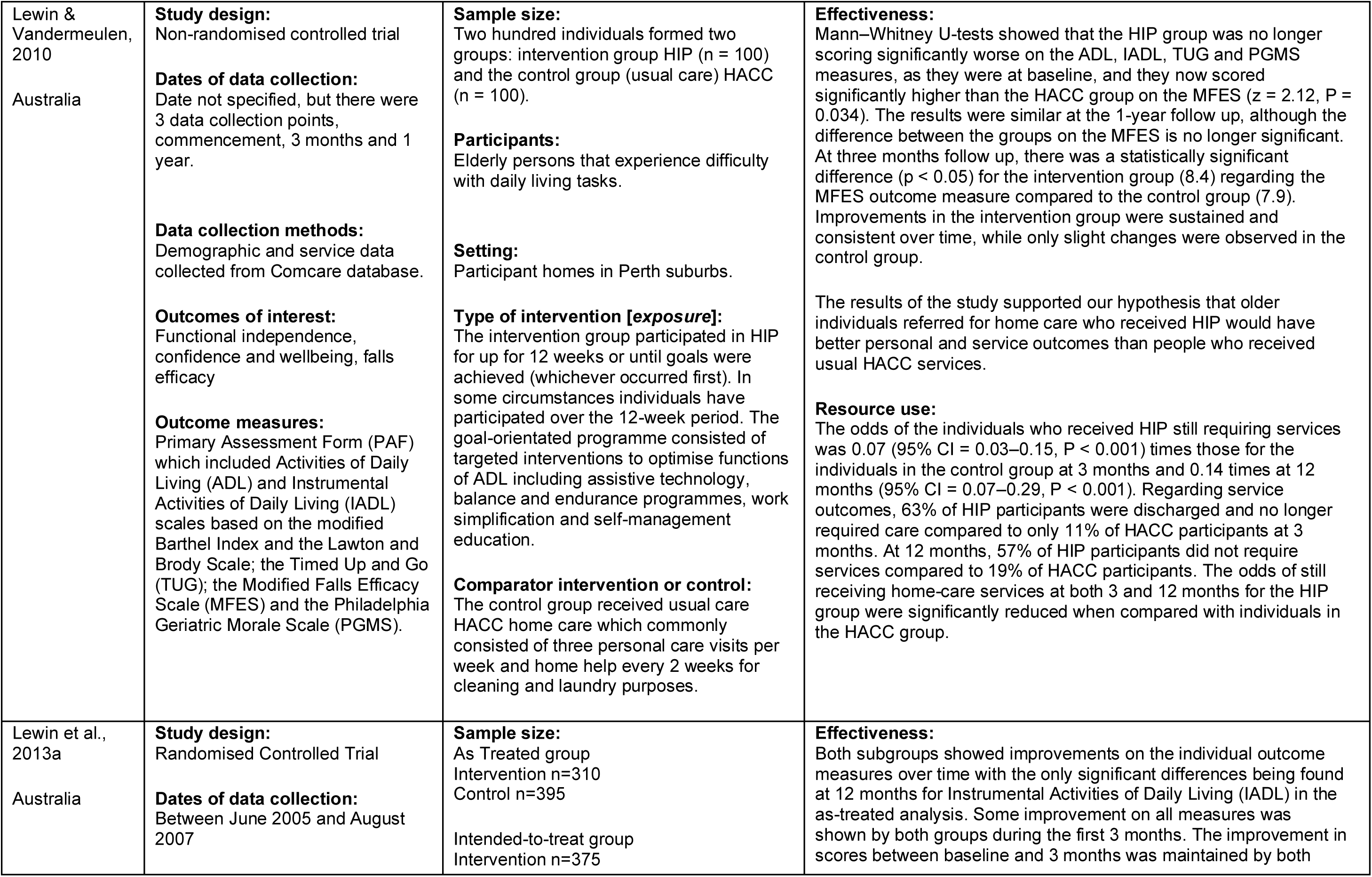
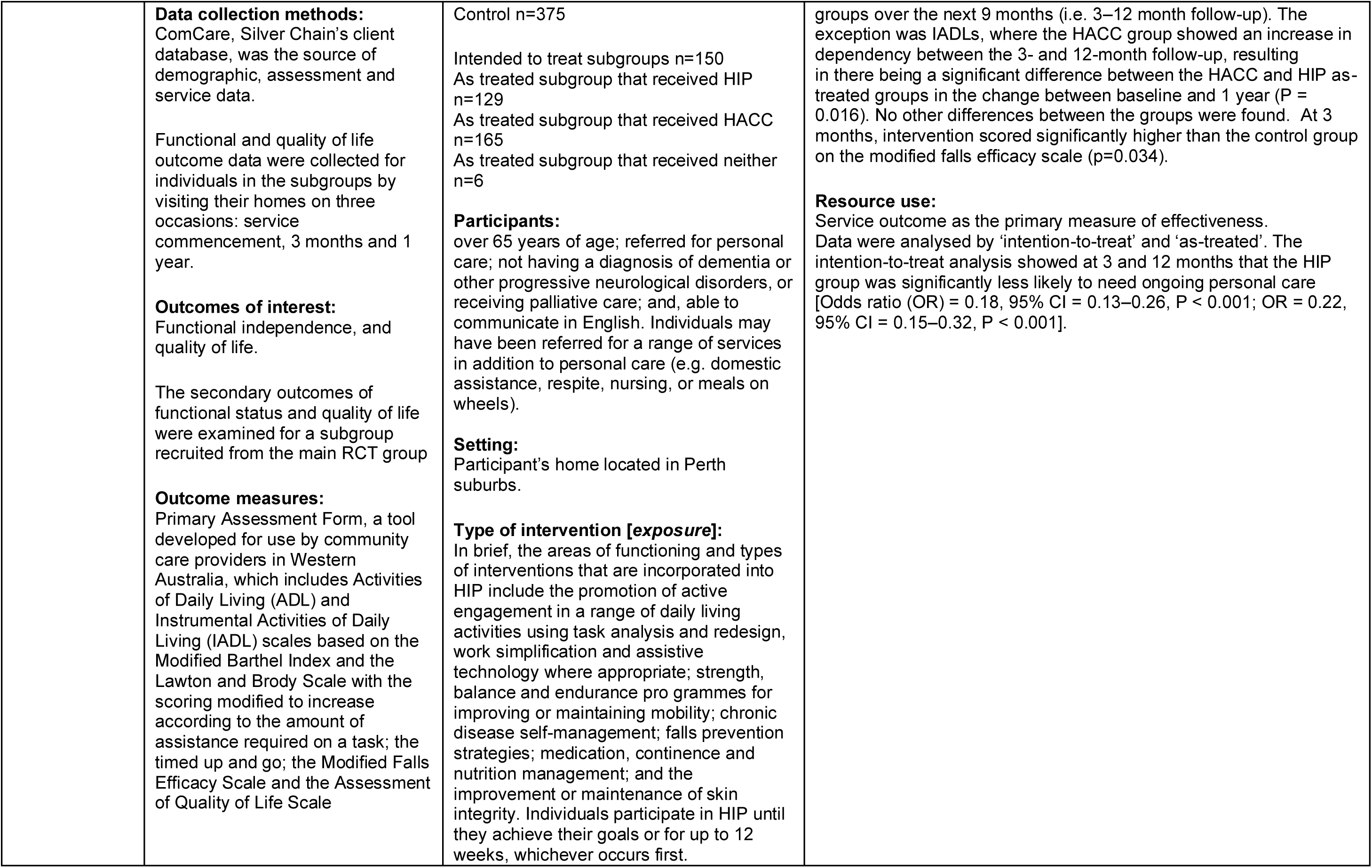
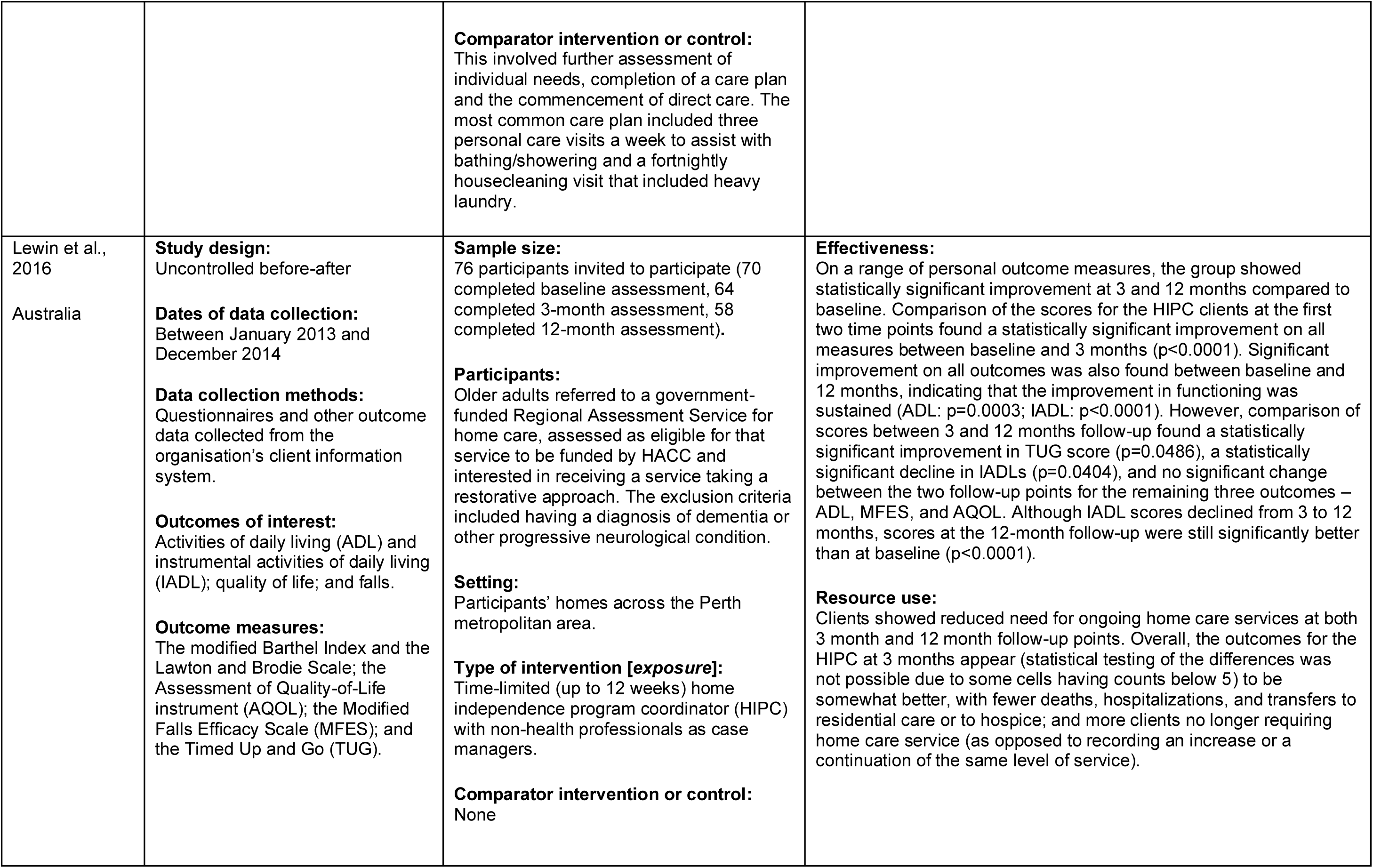
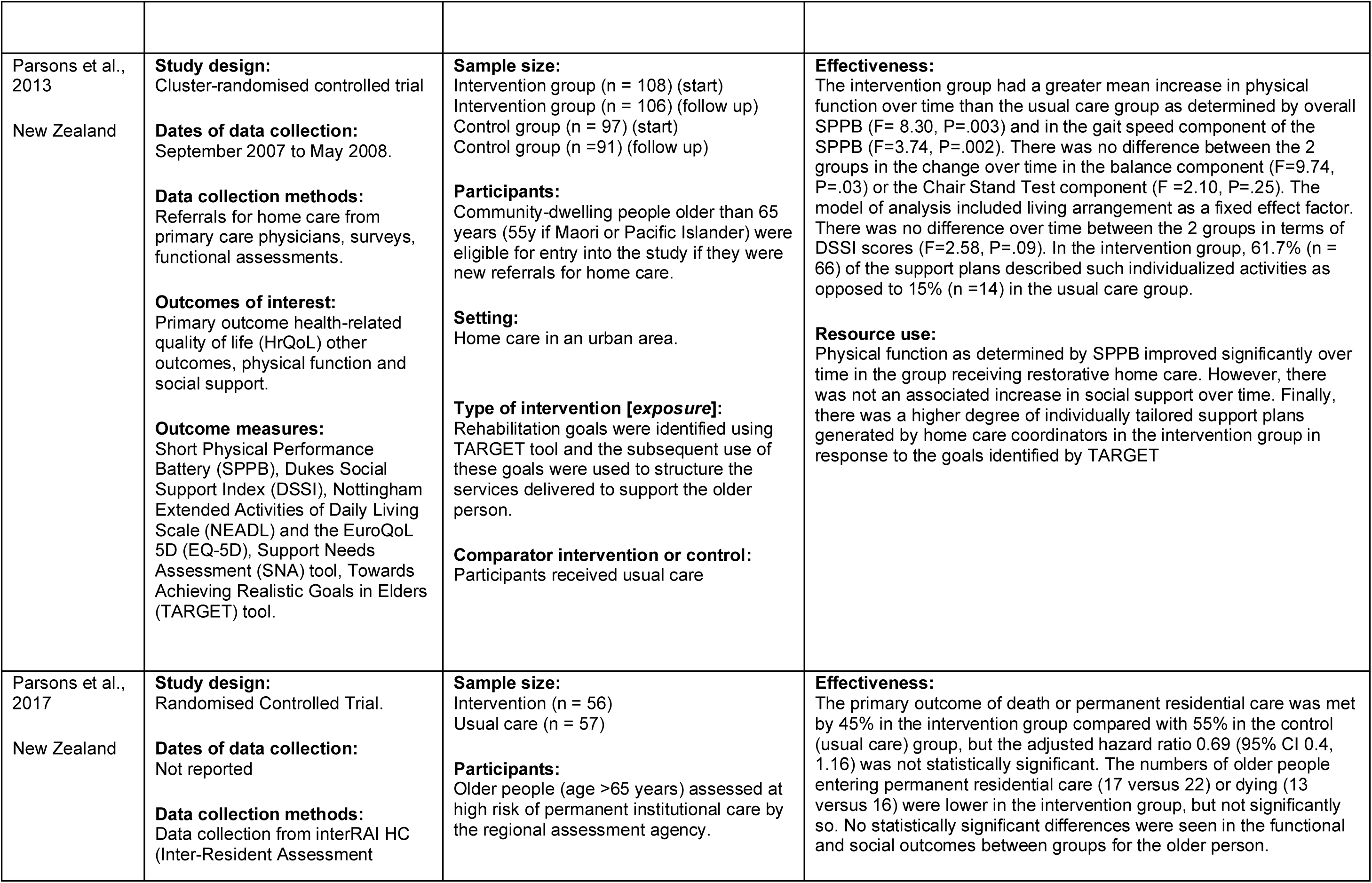
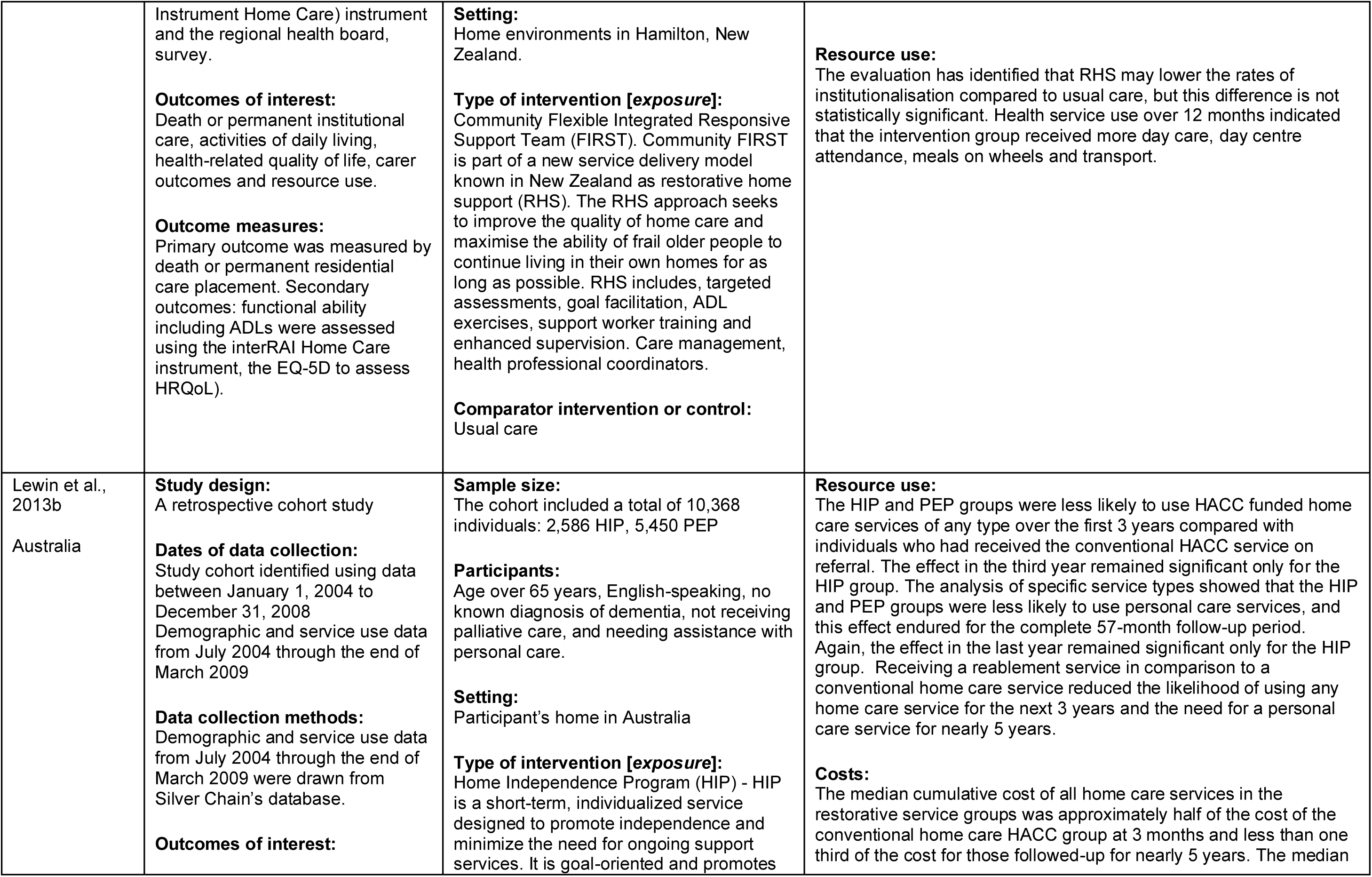
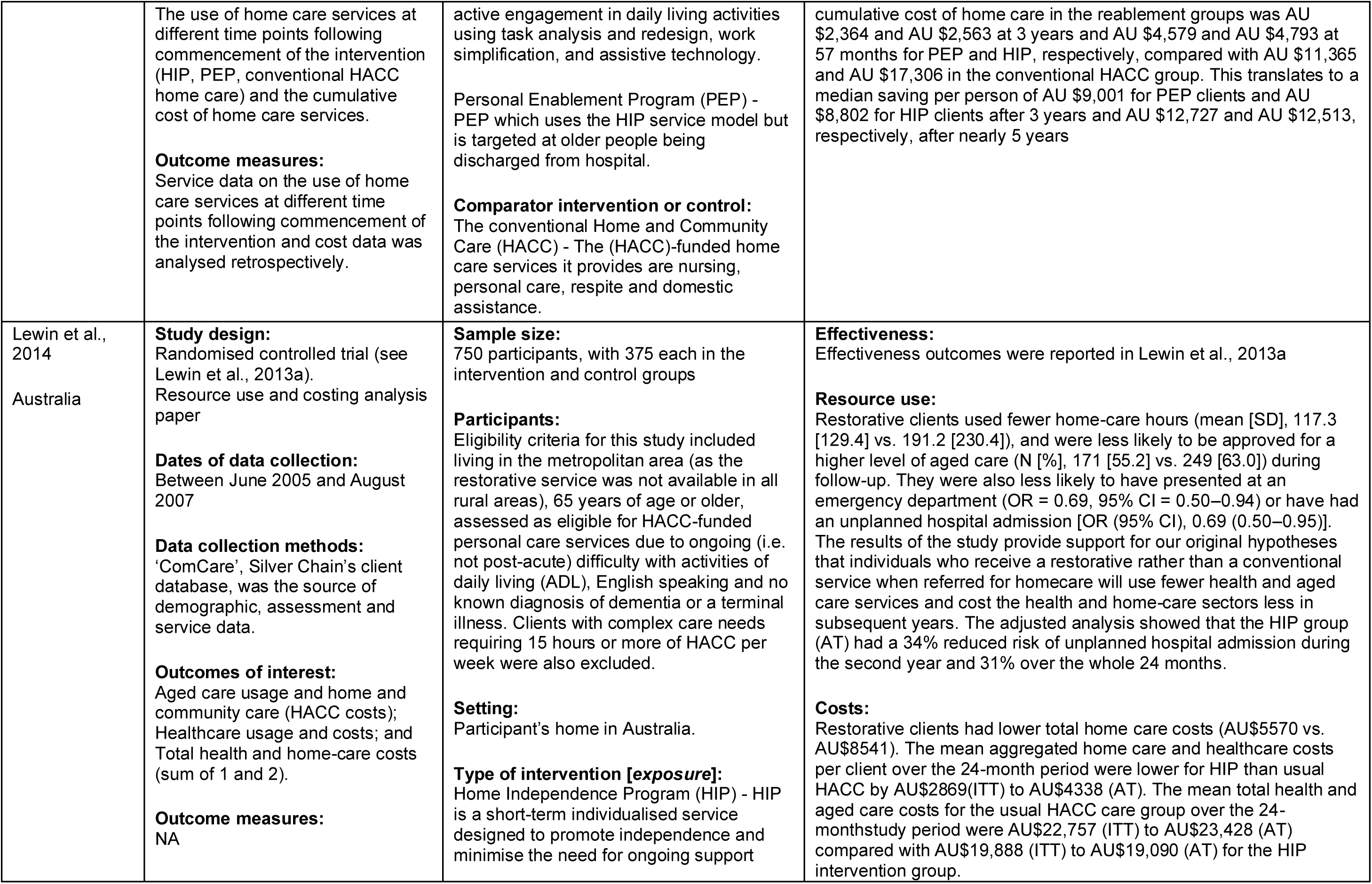
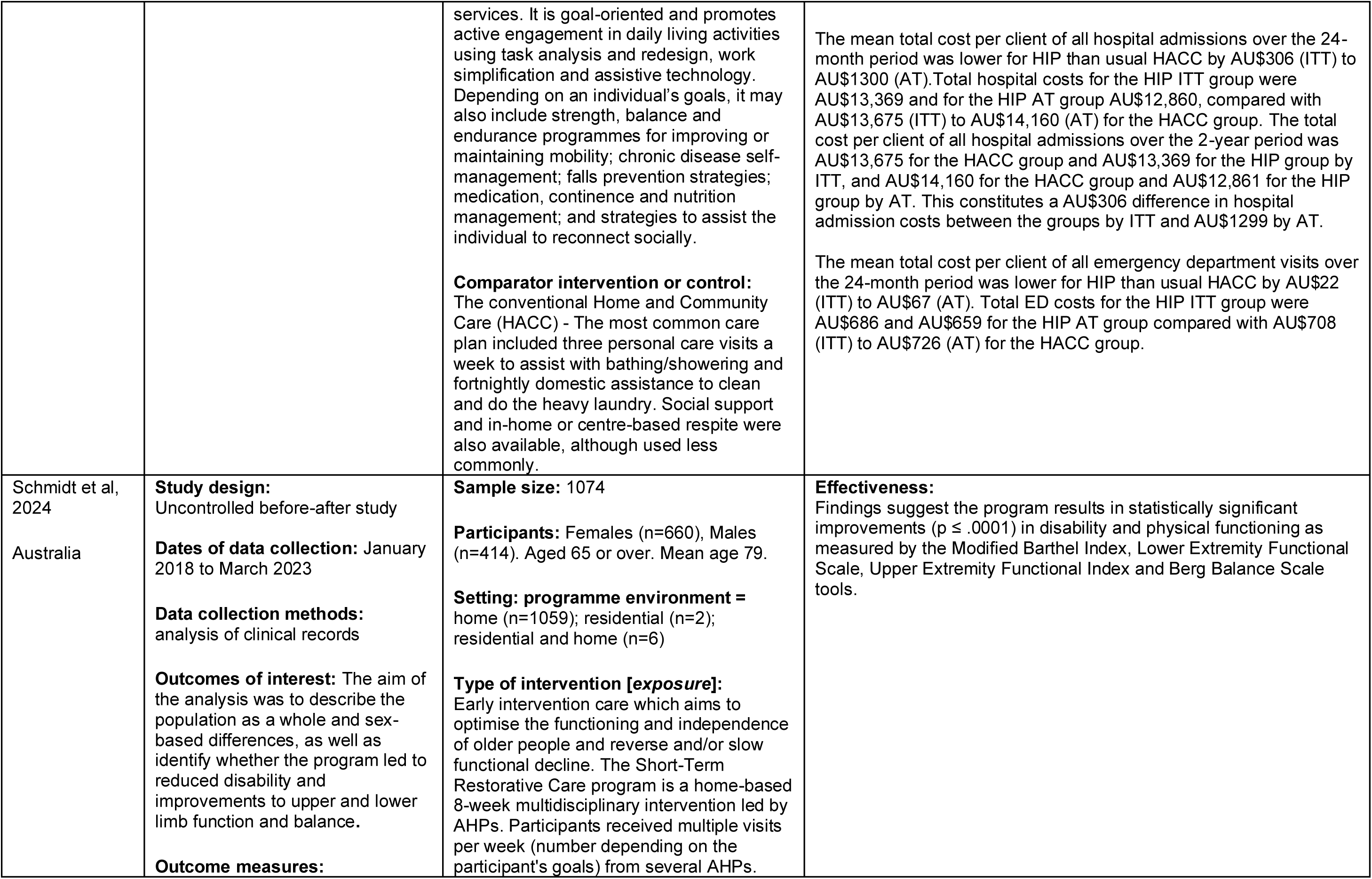
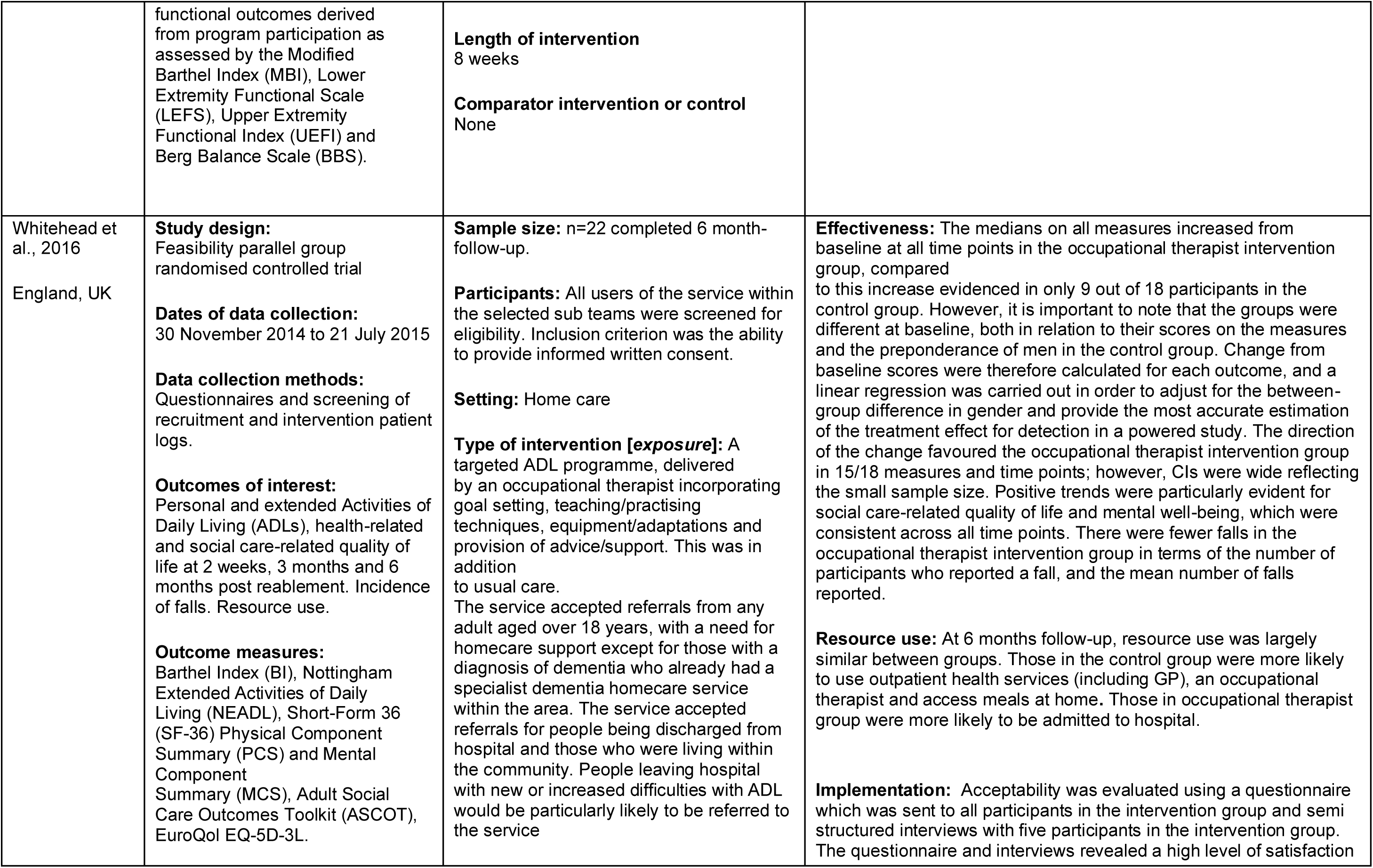
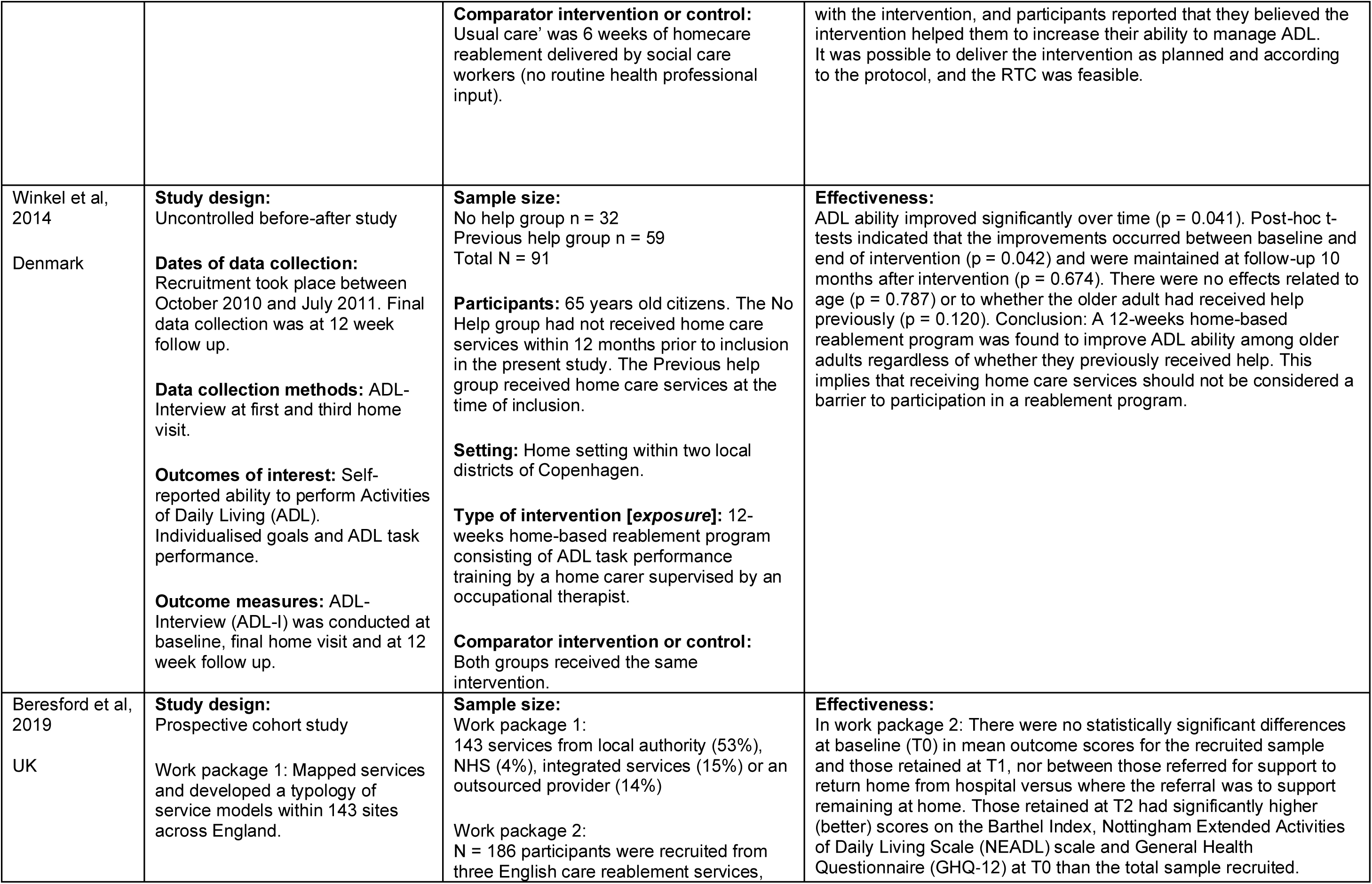
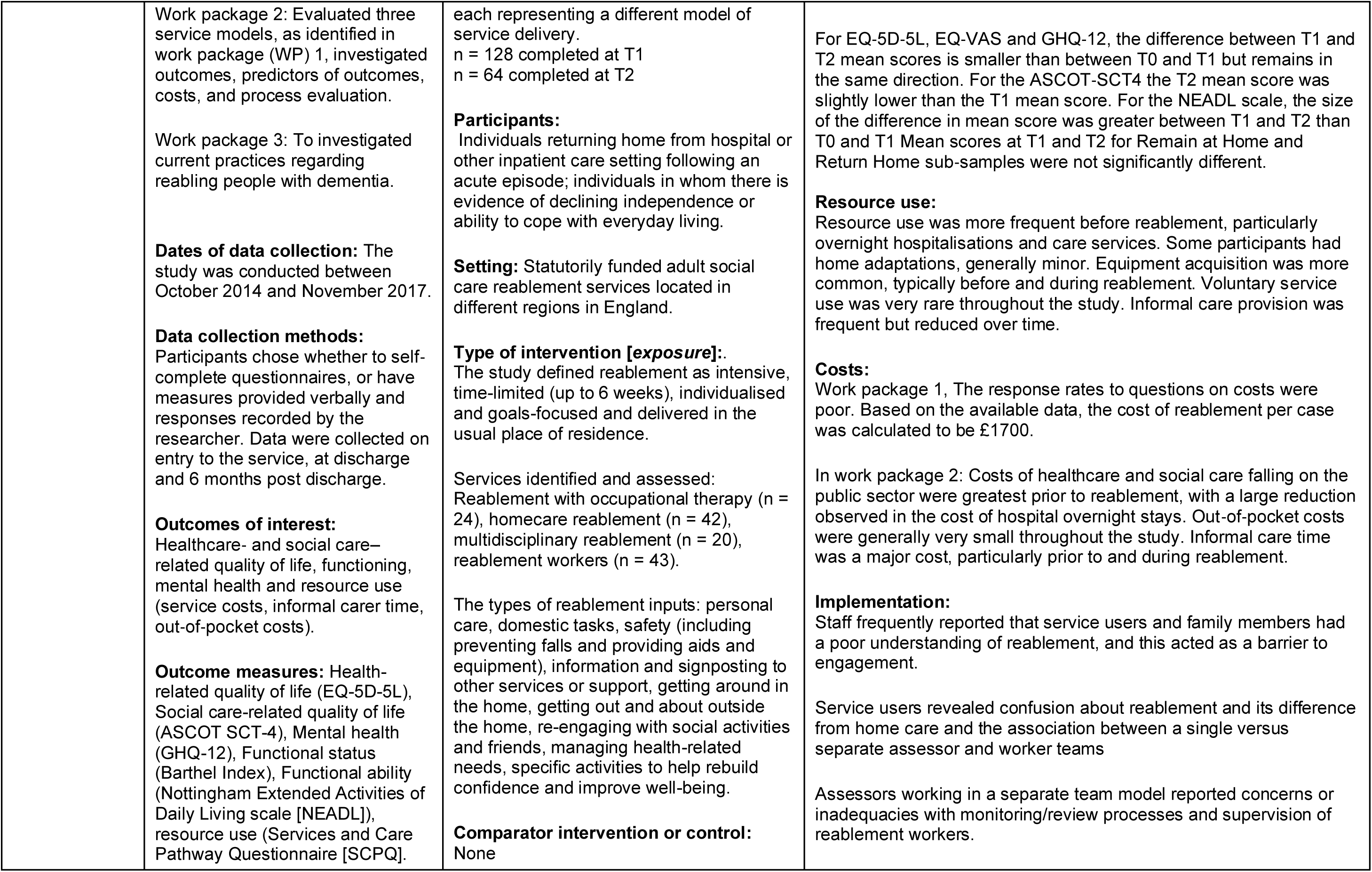
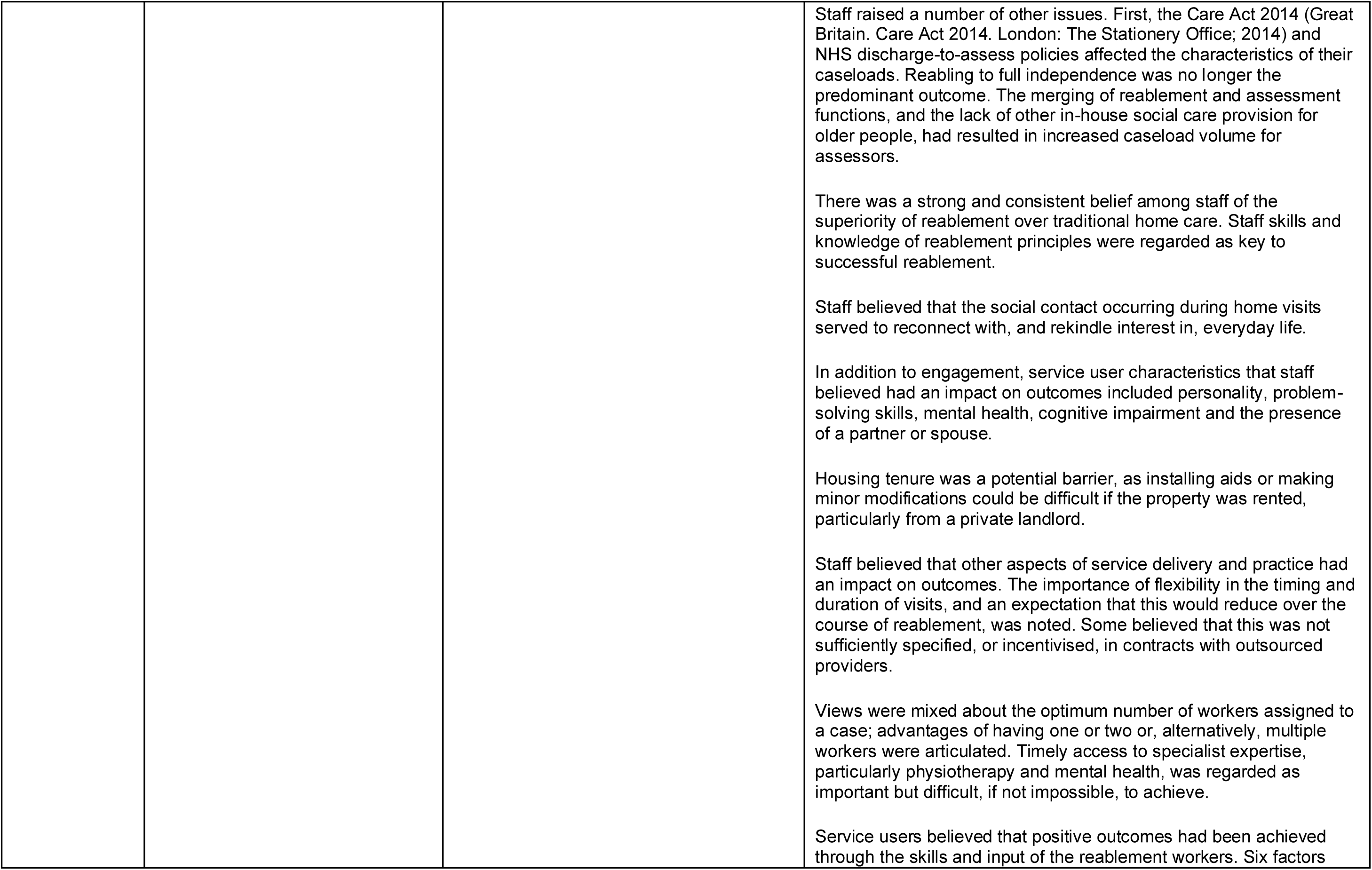
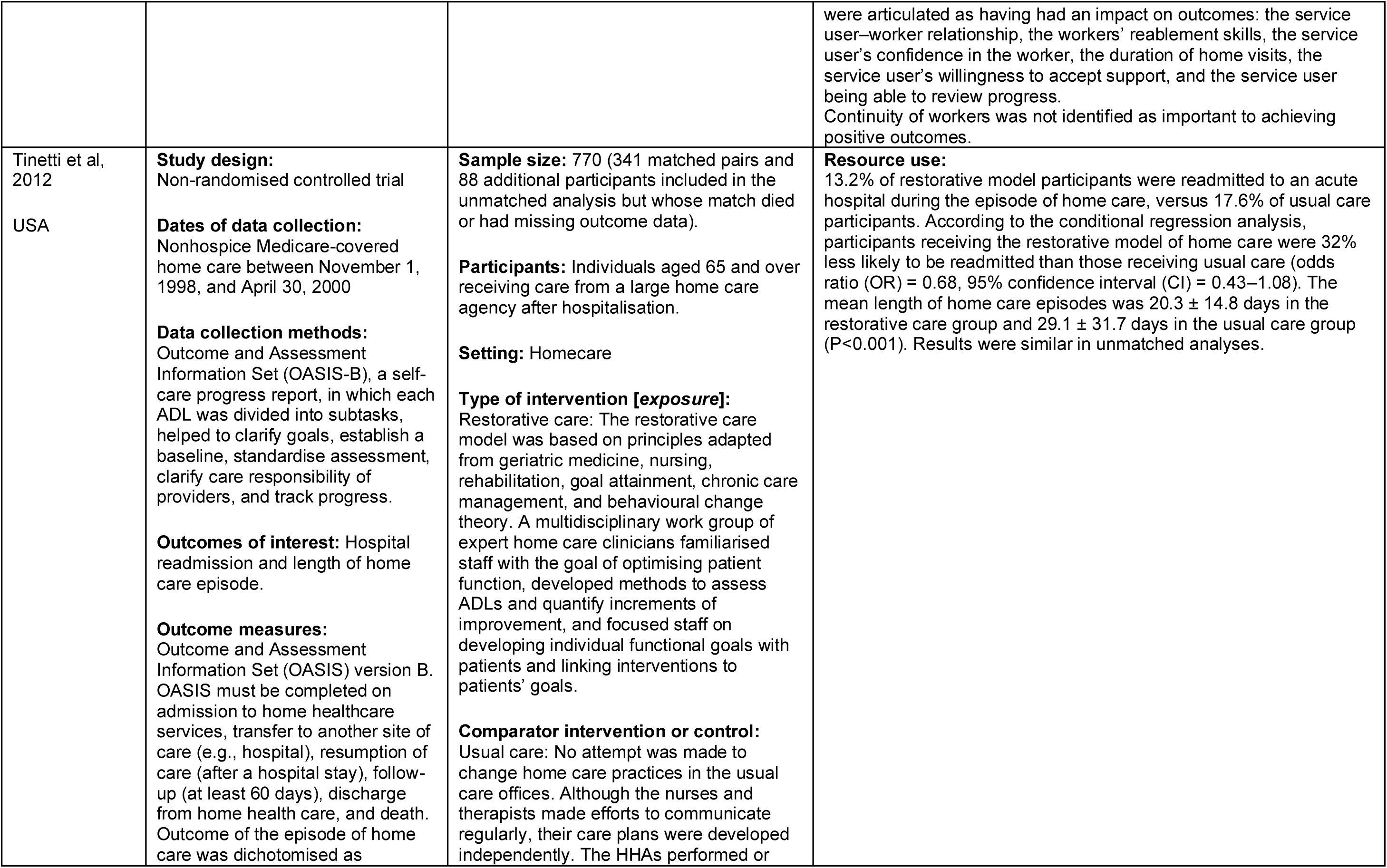
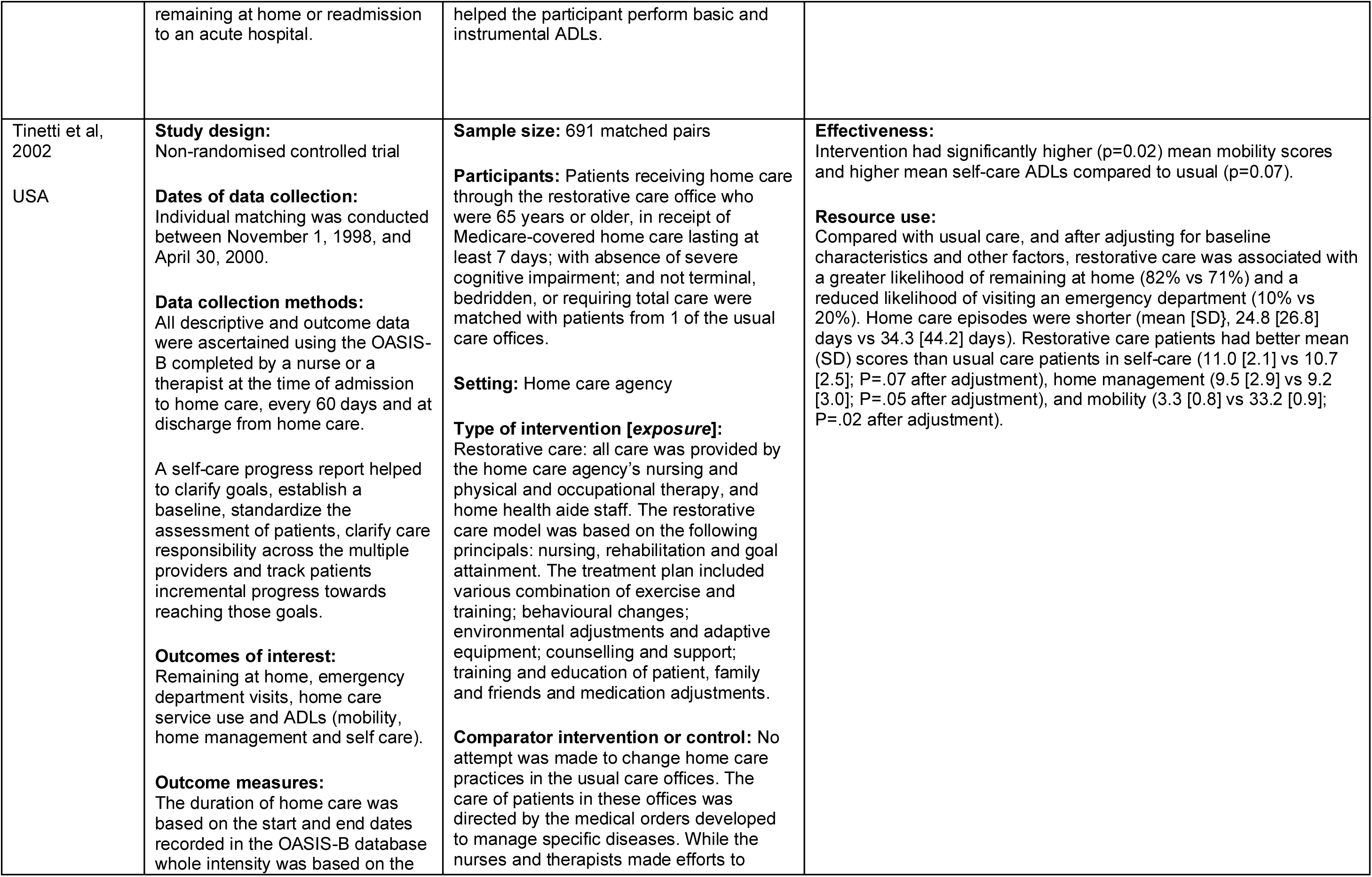
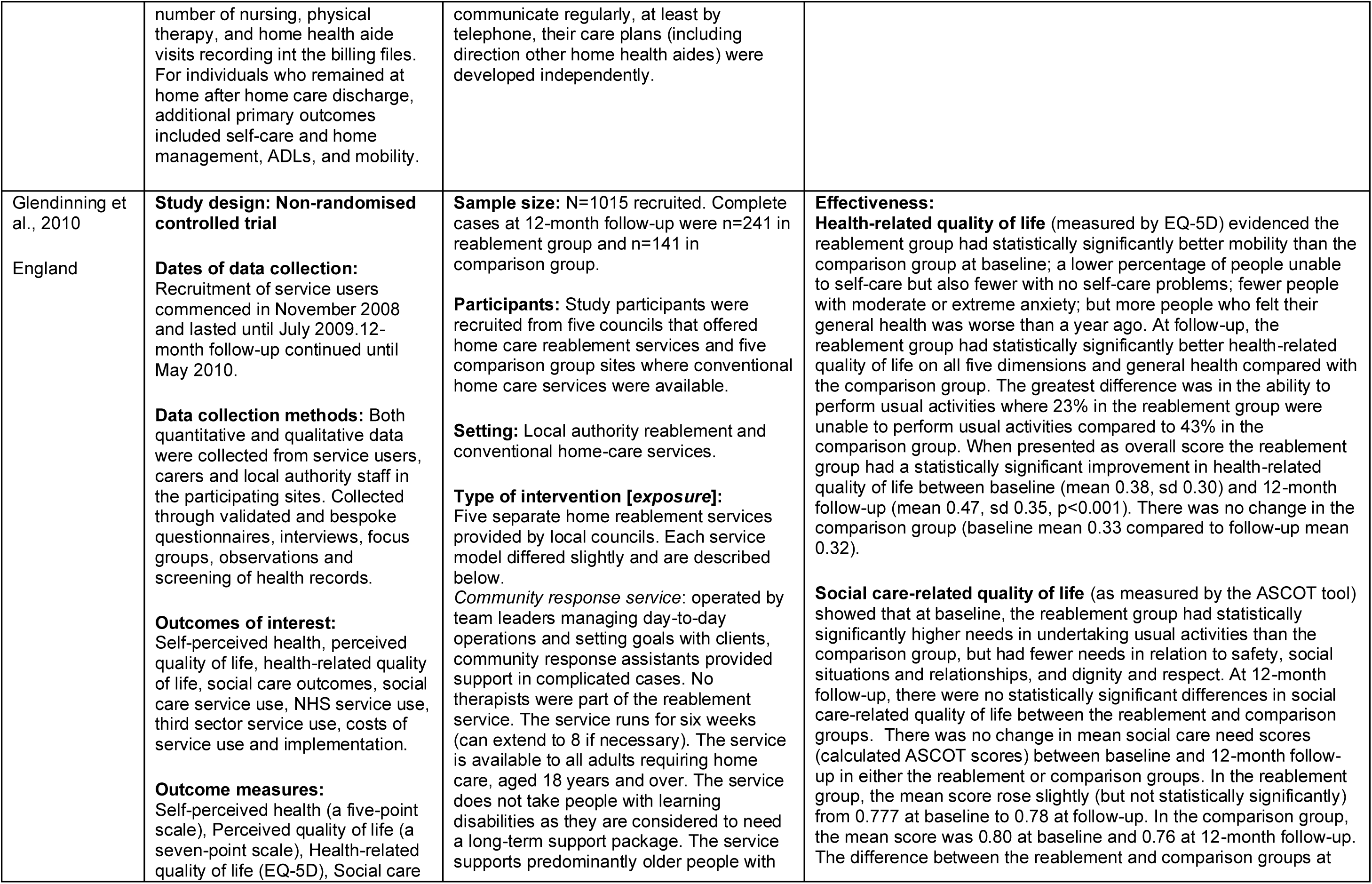
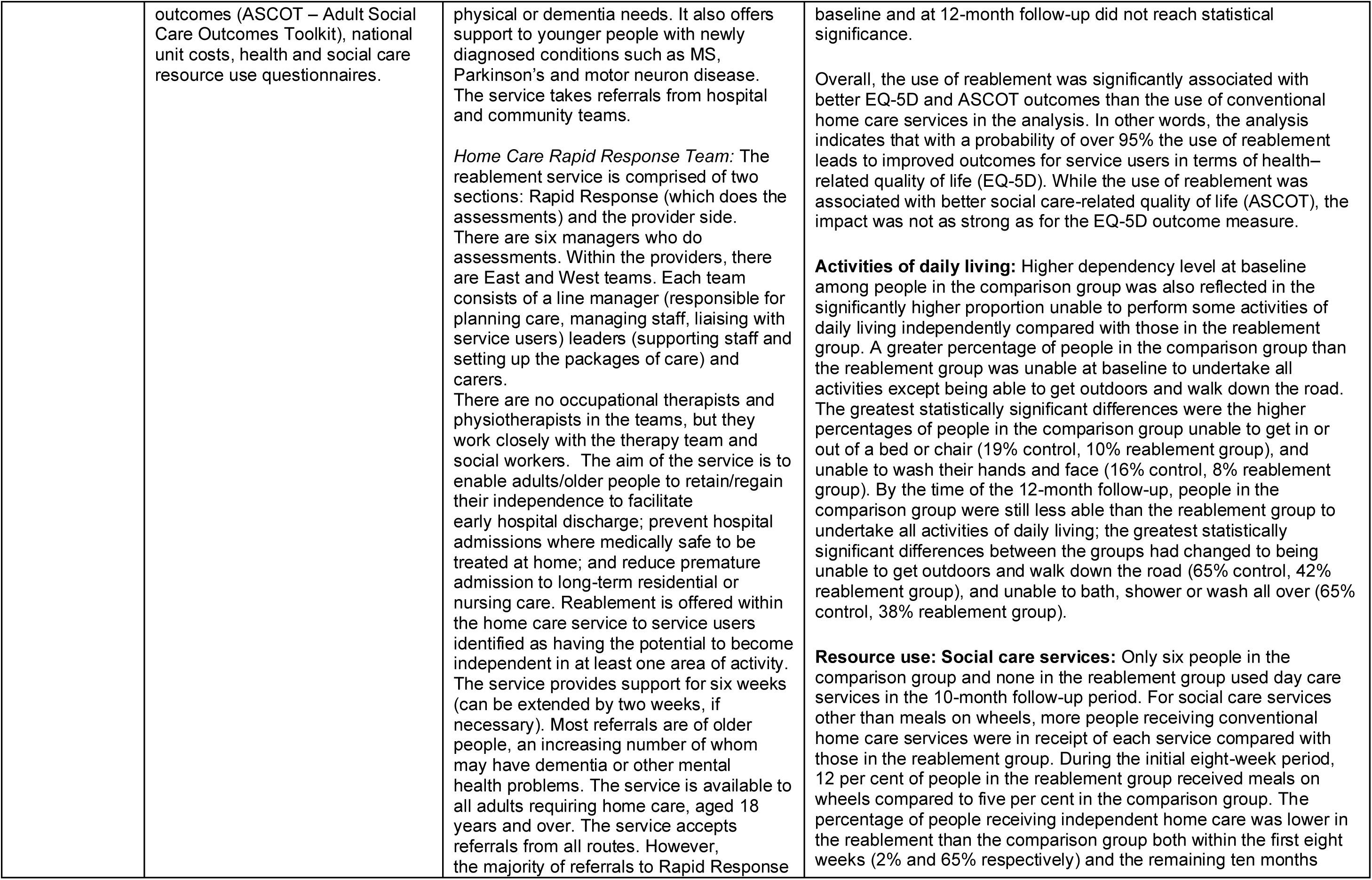
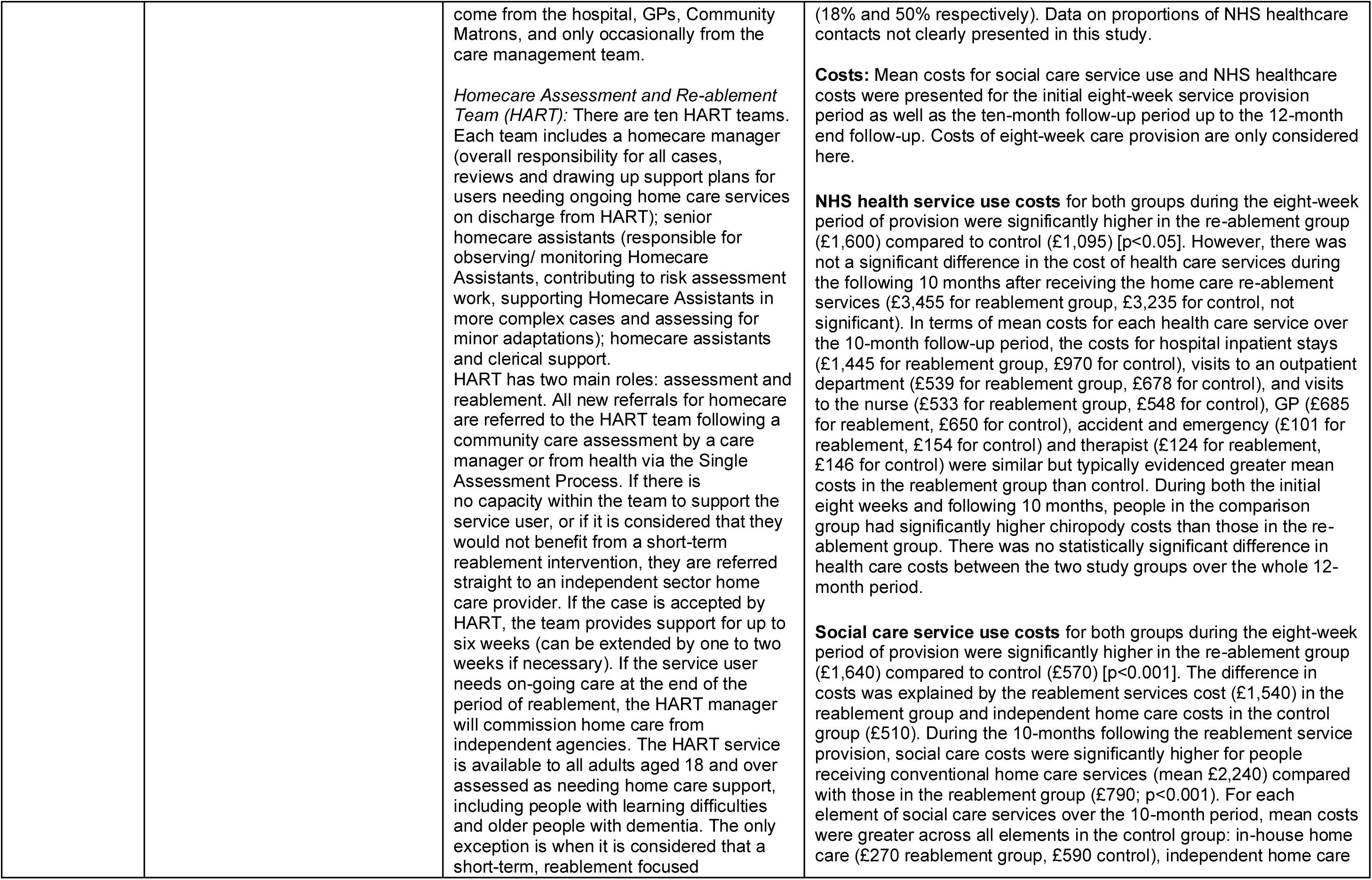
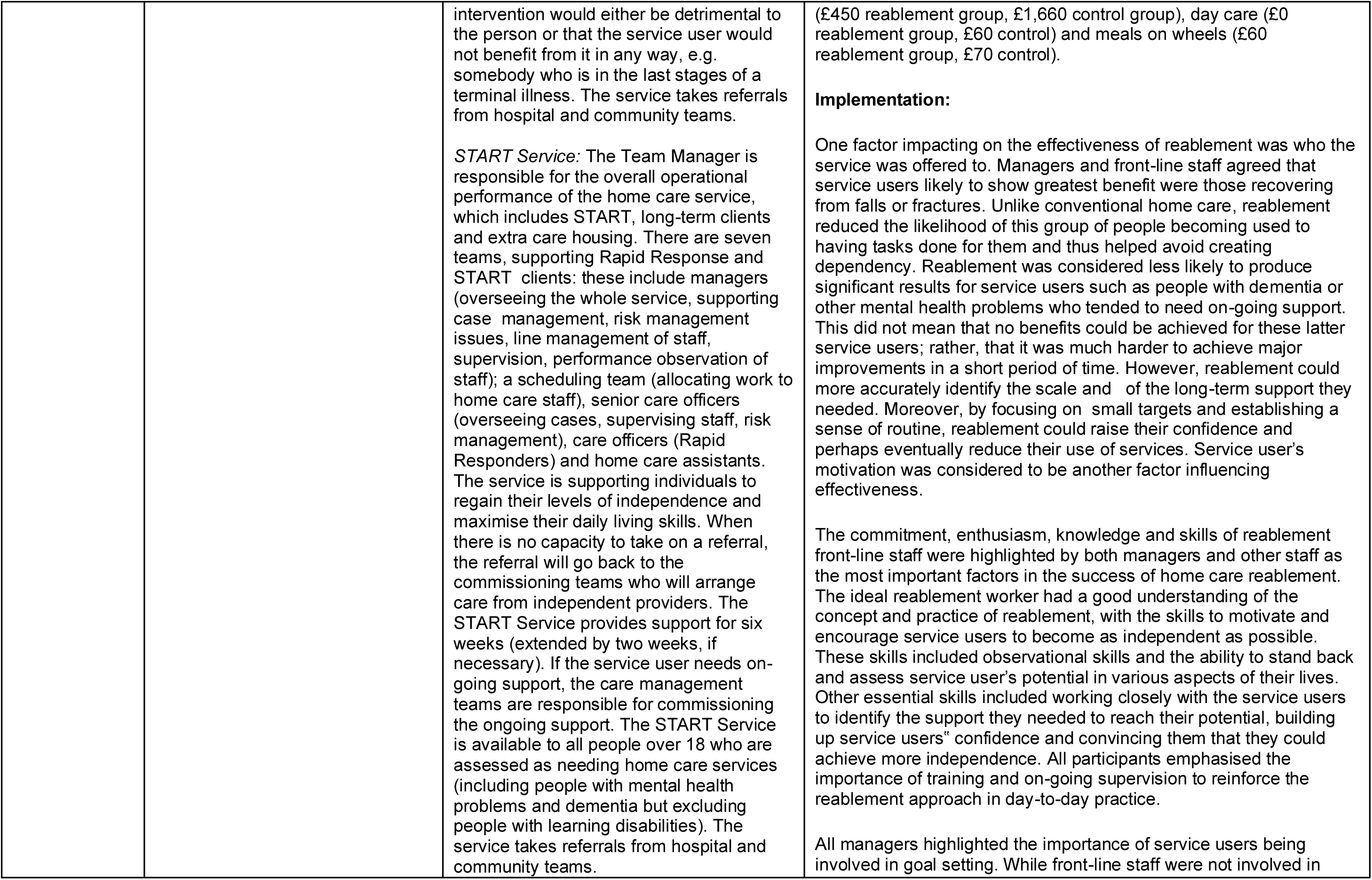
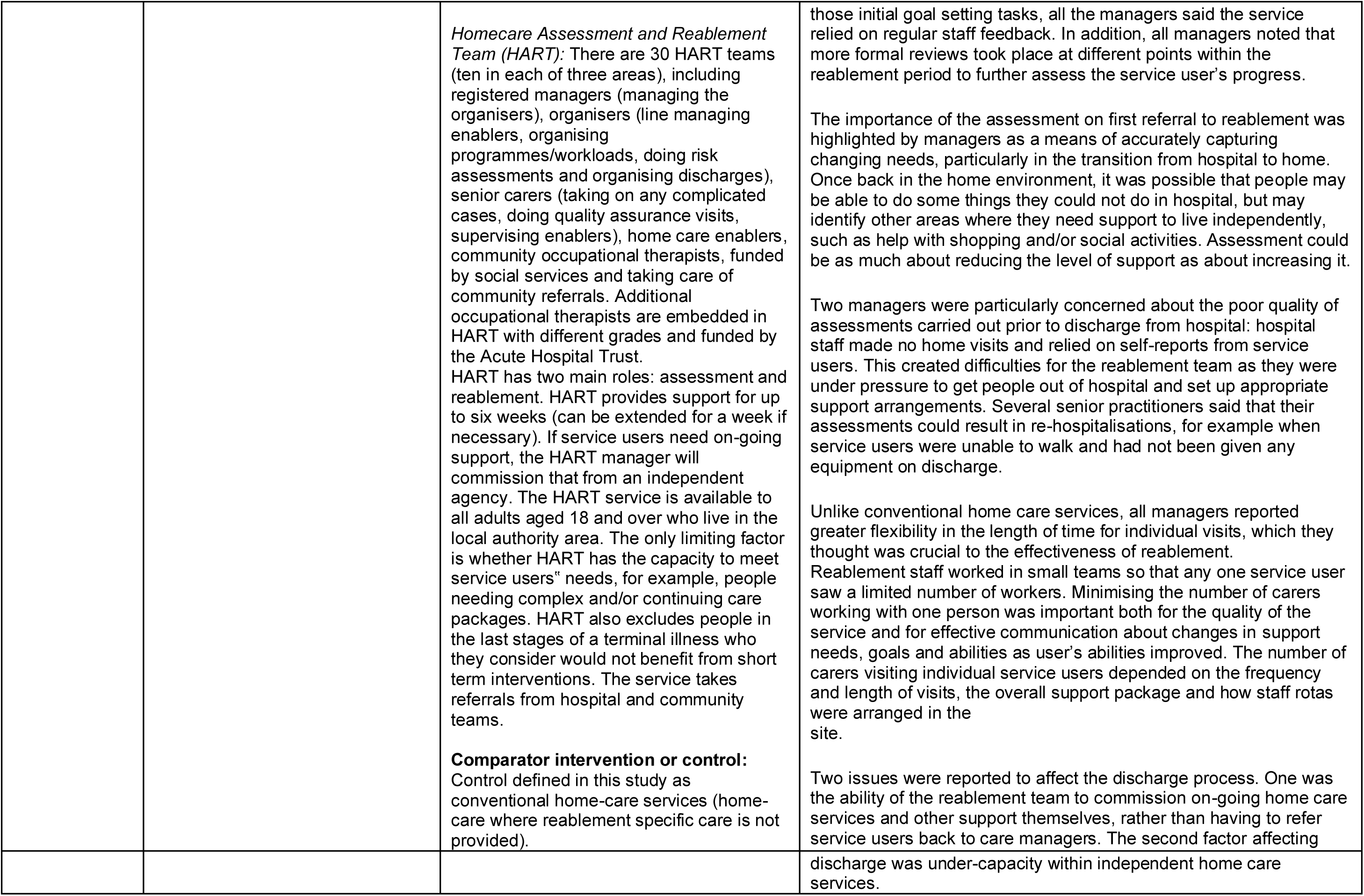
Summary of primary studies of clinical effectiveness.

| Citation<br>Country)<br>Aim | Study Details | Participants, setting and intervention | Key findings |
| --- | --- | --- | --- |
| Langeland et al., 2019<br>Norway | **Study design:**<br>Non-randomised controlled trial<br>**Dates of data collection:**<br>December 2014 until December 2015<br>**Data collection methods:**<br>Questionnaires completed face-to-face at participants homes.<br>**Outcomes of interest:**<br>Performance of daily activities, satisfaction with performance of daily activities, physical function, health-related quality of life, and coping relating to experiences of coherence.<br>**Outcome measures:**<br>The Canadian Occupational Performance Measure (COPM), the Short Physical Performance Measure Battery (SPPB), the EuroQoL-5 Dimension 5 level version (EQ-5D-5L), and the Sense of Coherence Questionnaire (SOC-13). | **Sample size:**<br>All participants (N=828); reablement group (n=707); control group (n=121)<br>**Participants:**<br>Home dwelling adults (18+ years) with functional decline.<br>**Setting:**<br>Participants' homes across 47 municipalities in Norway.<br>**Intervention deliverers:**<br>Multidisciplinary team involvement including an auxiliary nurse, physiotherapist, occupational therapist, nurse and home helper. An occupational therapist, physiotherapist, nurse, or social educator conducted the initial interview. Multiple home visits were then delivered by health care personnel to deliver daily training to participants to build their confidence and relearn skills.<br>**Type of intervention [exposure]:**<br>Time-limited reablement aimed at increasing/sustaining daily activities, physical function, health-related quality of life and coping as a sense of coherence. Paper does not provide details to<br>distinguish between step-up or step-down reablement.<br>**Comparator intervention or control:**<br>Standard care such as personal or practical assistance services, 'meals on wheels', safety alarms, or the provision of assistive technology. It could also involve rehabilitation efforts by health professionals such as occupational therapists, physiotherapists and nurses in the participant's home or local community | **Effectiveness:**<br>Significant treatment effects in the rehabilitation group compared with the control group were found in COPM-Performance and COPM-Satisfaction scores at 10 weeks (mean differences between groups (MD), 1.61, 95% confidence interval (CI), 1.13, 2.10 and MD 1.47, CI 0.98, 1.97, respectively), and at 6 months (MD 1.42; CI 0.82,2.02 and MD 1.37; CI 0.77,1.98, respectively). There were also significant treatment effects in the SPPB-subscale for balance and walking after 6 months, in the total SPPB score and in the subscale for sit-to-stand after 12 months. In the EQ-5D-5 L assessment, significant treatment effects were found in the subscales for mobility, and for usual activities and health after 6 months. There was a significant difference in the SOC after six months. |
| Tuntland et al., 2015<br>Norway | **Study design:**<br>Randomised controlled trial<br>**Dates of data collection:**<br>Not reported, the recruitment period lasted from May 2012 until February 2014.<br>**Data collection methods:**<br>Questionnaires and functional tests completed face-to-face at participants homes.<br>**Outcomes of interest:**<br>Self-perceived activity performance and satisfaction with that performance, functional mobility, grip strength, health-related quality of life.<br>**Outcome measures:**<br>The Canadian Occupational Performance Measure (COPM), the Timed Up and Go test, Jamar Dynamometer hydraulic grip instrument, and the COOP/Wonka. | **Sample size:**<br>Sixty-one participants were randomised to an intervention group (n = 31) or a control group (n = 30).<br>**Participants:**<br>Home-dwelling adults with functional decline<br>**Setting:**<br>Participants homes in one rural municipality in Norway.<br>**Type of intervention [exposure]:**<br>The intervention group received ten weeks of multicomponent home-based rehabilitation. The therapeutic process is tailored according to the needs and aims of the individual participant, resulting in differences in the number and type of elements in the intervention across participants<br>**Comparator intervention or control:**<br>Usual care such as personal or practical assistance services, 'meals on wheels', safety alarms, or the provision of assistive technology. Some participants received rehabilitation from occupational therapist and/or physical therapist. | **Effectiveness:**<br>There were significant improvements in mean scores favouring reablement in COPM performance at 3 months with a score of 1.5 points (p = 0.02), at 9 months 1.4 points (p = 0.03) and overall treatment 1.5 points (p = 0.01), and for COPM satisfaction at 9 months 1.4 points (p = 0.03) and overall treatment 1.2 points (p = 0.04). No significant group differences were found concerning COPM satisfaction at 3 months, physical capacity or health-related quality of life.<br>**Resource use:**<br>There was a significantly higher number of co-interventions at the 3-month follow-up in the control group; 12 outpatient treatments in the control group versus 3 outpatient treatments in the intervention group (p = 0.007), of which 10 of the outpatient treatments were physiotherapy. |
| Lewin & Vandermeulen, 2010<br>Australia | **Study design:**<br>Non-randomised controlled trial<br>**Dates of data collection:**<br>Date not specified, but there were 3 data collection points, commencement, 3 months and 1 year.<br>**Data collection methods:**<br>Demographic and service data collected from Comcare database.<br>**Outcomes of interest:**<br>Functional independence, confidence and wellbeing, falls efficacy<br>**Outcome measures:**<br>Primary Assessment Form (PAF) which included Activities of Daily Living (ADL) and Instrumental Activities of Daily Living (IADL) scales based on the modified Barthel Index and the Lawton and Brody Scale; the Timed Up and Go (TUG); the Modified Falls Efficacy Scale (MFES) and the Philadelphia Geriatric Morale Scale (PGMS). | **Sample size:**<br>Two hundred individuals formed two groups: intervention group HIP (n = 100) and the control group (usual care) HACC (n = 100).<br>**Participants:**<br>Elderly persons that experience difficulty with daily living tasks.<br>**Setting:**<br>Participant homes in Perth suburbs.<br>**Type of intervention [exposure]:**<br>The intervention group participated in HIP for up for 12 weeks or until goals were achieved (whichever occurred first). In some circumstances individuals have participated over the 12-week period. The goal-orientated programme consisted of targeted interventions to optimise functions of ADL including assistive technology, balance and endurance programmes, work simplification and self-management education.<br>**Comparator intervention or control:**<br>The control group received usual care HACC home care which commonly consisted of three personal care visits per week and home help every 2 weeks for cleaning and laundry purposes. | **Effectiveness:**<br>Mann–Whitney U-tests showed that the HIP group was no longer scoring significantly worse on the ADL, IADL, TUG and PGMS measures, as they were at baseline, and they now scored significantly higher than the HACC group on the MFES (z = 2.12, P = 0.034). The results were similar at the 1-year follow up, although the difference between the groups on the MFES is no longer significant. At three months follow up, there was a statistically significant difference (p < 0.05) for the intervention group (8.4) regarding the MFES outcome measure compared to the control group (7.9). Improvements in the intervention group were sustained and consistent over time, while only slight changes were observed in the control group.<br>The results of the study supported our hypothesis that older individuals referred for home care who received HIP would have better personal and service outcomes than people who received usual HACC services.<br>**Resource use:**<br>The odds of the individuals who received HIP still requiring services was 0.07 (95% CI = 0.03–0.15, P < 0.001) times those for the individuals in the control group at 3 months and 0.14 times at 12 months (95% CI = 0.07–0.29, P < 0.001). Regarding service outcomes, 63% of HIP participants were discharged and no longer required care compared to only 11% of HACC participants at 3 months. At 12 months, 57% of HIP participants did not require services compared to 19% of HACC participants. The odds of still receiving home-care services at both 3 and 12 months for the HIP group were significantly reduced when compared with individuals in the HACC group. |
| Lewin et al., 2013a<br>Australia | **Study design:**<br>Randomised Controlled Trial<br>**Dates of data collection:**<br>Between June 2005 and August 2007 | **Sample size:**<br>As Treated group<br>Intervention n=310<br>Control n=395<br>Intended-to-treat group<br>Intervention n=375 | **Effectiveness:**<br>Both subgroups showed improvements on the individual outcome measures over time with the only significant differences being found at 12 months for Instrumental Activities of Daily Living (IADL) in the as-treated analysis. Some improvement on all measures was shown by both groups during the first 3 months. The improvement in scores between baseline and 3 months was maintained by both |
|  | **Data collection methods:**<br>ComCare, Silver Chain's client database, was the source of demographic, assessment and service data.<br>Functional and quality of life outcome data were collected for individuals in the subgroups by visiting their homes on three occasions: service commencement, 3 months and 1 year.<br>**Outcomes of interest:**<br>Functional independence, and quality of life.<br>The secondary outcomes of functional status and quality of life were examined for a subgroup recruited from the main RCT group<br>**Outcome measures:**<br>Primary Assessment Form, a tool developed for use by community care providers in Western Australia, which includes Activities of Daily Living (ADL) and Instrumental Activities of Daily Living (IADL) scales based on the Modified Barthel Index and the Lawton and Brody Scale with the scoring modified to increase according to the amount of assistance required on a task; the timed up and go; the Modified Falls Efficacy Scale and the Assessment of Quality of Life Scale | Control n=375<br>Intended to treat subgroups n=150<br>As treated subgroup that received HIP n=129<br>As treated subgroup that received HACC n=165<br>As treated subgroup that received neither n=6<br>**Participants:**<br>over 65 years of age; referred for personal care; not having a diagnosis of dementia or other progressive neurological disorders, or receiving palliative care; and, able to communicate in English. Individuals may have been referred for a range of services in addition to personal care (e.g. domestic assistance, respite, nursing, or meals on wheels).<br>**Setting:**<br>Participant's home located in Perth suburbs.<br>**Type of intervention [exposure]:**<br>In brief, the areas of functioning and types of interventions that are incorporated into HIP include the promotion of active engagement in a range of daily living activities using task analysis and redesign, work simplification and assistive technology where appropriate; strength, balance and endurance programmes for improving or maintaining mobility; chronic disease self-management; falls prevention strategies; medication, continence and nutrition management; and the improvement or maintenance of skin integrity. Individuals participate in HIP until they achieve their goals or for up to 12 weeks, whichever occurs first. | groups over the next 9 months (i.e. 3–12 month follow-up). The exception was IADLs, where the HACC group showed an increase in dependency between the 3- and 12-month follow-up, resulting in there being a significant difference between the HACC and HIP as-treated groups in the change between baseline and 1 year ($P = 0.016$). No other differences between the groups were found. At 3 months, intervention scored significantly higher than the control group on the modified falls efficacy scale ($p=0.034$).<br>**Resource use:**<br>Service outcome as the primary measure of effectiveness. Data were analysed by 'intention-to-treat' and 'as-treated'. The intention-to-treat analysis showed at 3 and 12 months that the HIP group was significantly less likely to need ongoing personal care [Odds ratio (OR) = 0.18, 95% CI = 0.13–0.26, $P < 0.001$; OR = 0.22, 95% CI = 0.15–0.32, $P < 0.001$]. |
|  |  | **Comparator intervention or control:**<br>This involved further assessment of individual needs, completion of a care plan and the commencement of direct care. The most common care plan included three personal care visits a week to assist with bathing/showering and a fortnightly housecleaning visit that included heavy laundry. |  |
| Lewin et al., 2016<br>Australia | **Study design:**<br>Uncontrolled before-after<br>**Dates of data collection:**<br>Between January 2013 and December 2014<br>**Data collection methods:**<br>Questionnaires and other outcome data collected from the organisation's client information system.<br>**Outcomes of interest:**<br>Activities of daily living (ADL) and instrumental activities of daily living (IADL); quality of life; and falls.<br>**Outcome measures:**<br>The modified Barthel Index and the Lawton and Brodie Scale; the Assessment of Quality-of-Life instrument (AQOL); the Modified Falls Efficacy Scale (MFES); and the Timed Up and Go (TUG). | **Sample size:**<br>76 participants invited to participate (70 completed baseline assessment, 64 completed 3-month assessment, 58 completed 12-month assessment).<br>**Participants:**<br>Older adults referred to a government-funded Regional Assessment Service for home care, assessed as eligible for that service to be funded by HACC and interested in receiving a service taking a restorative approach. The exclusion criteria included having a diagnosis of dementia or other progressive neurological condition.<br>**Setting:**<br>Participants' homes across the Perth metropolitan area.<br>**Type of intervention [exposure]:**<br>Time-limited (up to 12 weeks) home independence program coordinator (HIPC) with non-health professionals as case managers.<br>**Comparator intervention or control:**<br>None | **Effectiveness:**<br>On a range of personal outcome measures, the group showed statistically significant improvement at 3 and 12 months compared to baseline. Comparison of the scores for the HIPC clients at the first two time points found a statistically significant improvement on all measures between baseline and 3 months ($p<0.0001$). Significant improvement on all outcomes was also found between baseline and 12 months, indicating that the improvement in functioning was sustained (ADL: $p=0.0003$; IADL: $p<0.0001$). However, comparison of scores between 3 and 12 months follow-up found a statistically significant improvement in TUG score ($p=0.0486$), a statistically significant decline in IADLs ($p=0.0404$), and no significant change between the two follow-up points for the remaining three outcomes – ADL, MFES, and AQOL. Although IADL scores declined from 3 to 12 months, scores at the 12-month follow-up were still significantly better than at baseline ($p<0.0001$).<br>**Resource use:**<br>Clients showed reduced need for ongoing home care services at both 3 month and 12 month follow-up points. Overall, the outcomes for the HIPC at 3 months appear (statistical testing of the differences was not possible due to some cells having counts below 5) to be somewhat better, with fewer deaths, hospitalizations, and transfers to residential care or to hospice; and more clients no longer requiring home care service (as opposed to recording an increase or a continuation of the same level of service). |
| Parsons et al., 2013<br>New Zealand | **Study design:**<br>Cluster-randomised controlled trial<br>**Dates of data collection:**<br>September 2007 to May 2008.<br>**Data collection methods:**<br>Referrals for home care from primary care physicians, surveys, functional assessments.<br>**Outcomes of interest:**<br>Primary outcome health-related quality of life (HrQoL) other outcomes, physical function and social support.<br>**Outcome measures:**<br>Short Physical Performance Battery (SPPB), Dukes Social Support Index (DSSI), Nottingham Extended Activities of Daily Living Scale (NEADL) and the EuroQoL 5D (EQ-5D), Support Needs Assessment (SNA) tool, Towards Achieving Realistic Goals in Elders (TARGET) tool. | **Sample size:**<br>Intervention group (n = 108) (start)<br>Intervention group (n = 106) (follow up)<br>Control group (n = 97) (start)<br>Control group (n = 91) (follow up)<br>**Participants:**<br>Community-dwelling people older than 65 years (55y if Maori or Pacific Islander) were eligible for entry into the study if they were new referrals for home care.<br>**Setting:**<br>Home care in an urban area.<br>**Type of intervention [exposure]:**<br>Rehabilitation goals were identified using TARGET tool and the subsequent use of these goals were used to structure the services delivered to support the older person.<br>**Comparator intervention or control:**<br>Participants received usual care | **Effectiveness:**<br>The intervention group had a greater mean increase in physical function over time than the usual care group as determined by overall SPPB (F= 8.30, P=.003) and in the gait speed component of the SPPB (F=3.74, P=.002). There was no difference between the 2 groups in the change over time in the balance component (F=9.74, P=.03) or the Chair Stand Test component (F =2.10, P=.25). The model of analysis included living arrangement as a fixed effect factor. There was no difference over time between the 2 groups in terms of DSSI scores (F=2.58, P=.09). In the intervention group, 61.7% (n = 66) of the support plans described such individualized activities as opposed to 15% (n =14) in the usual care group.<br>**Resource use:**<br>Physical function as determined by SPPB improved significantly over time in the group receiving restorative home care. However, there was not an associated increase in social support over time. Finally, there was a higher degree of individually tailored support plans generated by home care coordinators in the intervention group in response to the goals identified by TARGET |
| Parsons et al., 2017<br>New Zealand | **Study design:**<br>Randomised Controlled Trial.<br>**Dates of data collection:**<br>Not reported<br>**Data collection methods:**<br>Data collection from interRAI HC (Inter-Resident Assessment | **Sample size:**<br>Intervention (n = 56)<br>Usual care (n = 57)<br>**Participants:**<br>Older people (age >65 years) assessed at high risk of permanent institutional care by the regional assessment agency. | **Effectiveness:**<br>The primary outcome of death or permanent residential care was met by 45% in the intervention group compared with 55% in the control (usual care) group, but the adjusted hazard ratio 0.69 (95% CI 0.4, 1.16) was not statistically significant. The numbers of older people entering permanent residential care (17 versus 22) or dying (13 versus 16) were lower in the intervention group, but not significantly so. No statistically significant differences were seen in the functional and social outcomes between groups for the older person. |
|  | Instrument Home Care) instrument and the regional health board, survey.<br>**Outcomes of interest:**<br>Death or permanent institutional care, activities of daily living, health-related quality of life, carer outcomes and resource use.<br>**Outcome measures:**<br>Primary outcome was measured by death or permanent residential care placement. Secondary outcomes: functional ability including ADLs were assessed using the interRAI Home Care instrument, the EQ-5D to assess HRQoL). | **Setting:**<br>Home environments in Hamilton, New Zealand.<br>**Type of intervention [exposure]:**<br>Community Flexible Integrated Responsive Support Team (FIRST). Community FIRST is part of a new service delivery model known in New Zealand as restorative home support (RHS). The RHS approach seeks to improve the quality of home care and maximise the ability of frail older people to continue living in their own homes for as long as possible. RHS includes, targeted assessments, goal facilitation, ADL exercises, support worker training and enhanced supervision. Care management, health professional coordinators.<br>**Comparator intervention or control:**<br>Usual care | **Resource use:**<br>The evaluation has identified that RHS may lower the rates of institutionalisation compared to usual care, but this difference is not statistically significant. Health service use over 12 months indicated that the intervention group received more day care, day centre attendance, meals on wheels and transport. |
| Lewin et al., 2013b<br>Australia | **Study design:**<br>A retrospective cohort study<br>**Dates of data collection:**<br>Study cohort identified using data between January 1, 2004 to December 31, 2008<br>Demographic and service use data from July 2004 through the end of March 2009<br>**Data collection methods:**<br>Demographic and service use data from July 2004 through the end of March 2009 were drawn from Silver Chain's database.<br>**Outcomes of interest:** | **Sample size:**<br>The cohort included a total of 10,368 individuals: 2,586 HIP, 5,450 PEP<br>**Participants:**<br>Age over 65 years, English-speaking, no known diagnosis of dementia, not receiving palliative care, and needing assistance with personal care.<br>**Setting:**<br>Participant's home in Australia<br>**Type of intervention [exposure]:**<br>Home Independence Program (HIP) - HIP is a short-term, individualized service designed to promote independence and minimize the need for ongoing support services. It is goal-oriented and promotes | **Resource use:**<br>The HIP and PEP groups were less likely to use HACC funded home care services of any type over the first 3 years compared with individuals who had received the conventional HACC service on referral. The effect in the third year remained significant only for the HIP group. The analysis of specific service types showed that the HIP and PEP groups were less likely to use personal care services, and this effect endured for the complete 57-month follow-up period. Again, the effect in the last year remained significant only for the HIP group. Receiving a reablement service in comparison to a conventional home care service reduced the likelihood of using any home care service for the next 3 years and the need for a personal care service for nearly 5 years.<br>**Costs:**<br>The median cumulative cost of all home care services in the restorative service groups was approximately half of the cost of the conventional home care HACC group at 3 months and less than one third of the cost for those followed-up for nearly 5 years. The median |
| | The use of home care services at different time points following commencement of the intervention (HIP, PEP, conventional HACC home care) and the cumulative cost of home care services.<br>**Outcome measures:**<br>Service data on the use of home care services at different time points following commencement of the intervention and cost data was analysed retrospectively. | active engagement in daily living activities using task analysis and redesign, work simplification, and assistive technology.<br>Personal Enablement Program (PEP) - PEP which uses the HIP service model but is targeted at older people being discharged from hospital.<br>**Comparator intervention or control:**<br>The conventional Home and Community Care (HACC) - The (HACC)-funded home care services it provides are nursing, personal care, respite and domestic assistance. | cumulative cost of home care in the reablement groups was AU \$2,364 and AU \$2,563 at 3 years and AU \$4,579 and AU \$4,793 at 57 months for PEP and HIP, respectively, compared with AU \$11,365 and AU \$17,306 in the conventional HACC group. This translates to a median saving per person of AU \$9,001 for PEP clients and AU \$8,802 for HIP clients after 3 years and AU \$12,727 and AU \$12,513, respectively, after nearly 5 years |
| Lewin et al., 2014<br>Australia | **Study design:**<br>Randomised controlled trial (see Lewin et al., 2013a).<br>Resource use and costing analysis paper<br>**Dates of data collection:**<br>Between June 2005 and August 2007<br>**Data collection methods:**<br>'ComCare', Silver Chain's client database, was the source of demographic, assessment and service data.<br>**Outcomes of interest:**<br>Aged care usage and home and community care (HACC costs); Healthcare usage and costs; and Total health and home-care costs (sum of 1 and 2).<br>**Outcome measures:**<br>NA | **Sample size:**<br>750 participants, with 375 each in the intervention and control groups<br>**Participants:**<br>Eligibility criteria for this study included living in the metropolitan area (as the restorative service was not available in all rural areas), 65 years of age or older, assessed as eligible for HACC-funded personal care services due to ongoing (i.e. not post-acute) difficulty with activities of daily living (ADL), English speaking and no known diagnosis of dementia or a terminal illness. Clients with complex care needs requiring 15 hours or more of HACC per week were also excluded.<br>**Setting:**<br>Participant's home in Australia.<br>**Type of intervention [exposure]:**<br>Home Independence Program (HIP) - HIP is a short-term individualised service designed to promote independence and minimise the need for ongoing support | **Effectiveness:**<br>Effectiveness outcomes were reported in Lewin et al., 2013a<br>**Resource use:**<br>Restorative clients used fewer home-care hours (mean [SD], 117.3 [129.4] vs. 191.2 [230.4]), and were less likely to be approved for a higher level of aged care (N [%], 171 [55.2] vs. 249 [63.0]) during follow-up. They were also less likely to have presented at an emergency department (OR = 0.69, 95% CI = 0.50–0.94) or have had an unplanned hospital admission [OR (95% CI), 0.69 (0.50–0.95)]. The results of the study provide support for our original hypotheses that individuals who receive a restorative rather than a conventional service when referred for homecare will use fewer health and aged care services and cost the health and home-care sectors less in subsequent years. The adjusted analysis showed that the HIP group (AT) had a 34% reduced risk of unplanned hospital admission during the second year and 31% over the whole 24 months.<br>**Costs:**<br>Restorative clients had lower total home care costs (AU\$5570 vs. AU\$8541). The mean aggregated home care and healthcare costs per client over the 24-month period were lower for HIP than usual HACC by AU\$2869 (ITT) to AU\$4338 (AT). The mean total health and aged care costs for the usual HACC care group over the 24-month study period were AU\$22,757 (ITT) to AU\$23,428 (AT) compared with AU\$19,888 (ITT) to AU\$19,090 (AT) for the HIP intervention group. |
| | | services. It is goal-oriented and promotes active engagement in daily living activities using task analysis and redesign, work simplification and assistive technology. Depending on an individual's goals, it may also include strength, balance and endurance programmes for improving or maintaining mobility; chronic disease self-management; falls prevention strategies; medication, continence and nutrition management; and strategies to assist the individual to reconnect socially.<br>**Comparator intervention or control:**<br>The conventional Home and Community Care (HACC) - The most common care plan included three personal care visits a week to assist with bathing/showering and fortnightly domestic assistance to clean and do the heavy laundry. Social support and in-home or centre-based respite were also available, although used less commonly. | The mean total cost per client of all hospital admissions over the 24-month period was lower for HIP than usual HACC by AU\$306 (ITT) to AU\$1300 (AT). Total hospital costs for the HIP ITT group were AU\$13,369 and for the HIP AT group AU\$12,860, compared with AU\$13,675 (ITT) to AU\$14,160 (AT) for the HACC group. The total cost per client of all hospital admissions over the 2-year period was AU\$13,675 for the HACC group and AU\$13,369 for the HIP group by ITT, and AU\$14,160 for the HACC group and AU\$12,861 for the HIP group by AT. This constitutes a AU\$306 difference in hospital admission costs between the groups by ITT and AU\$1299 by AT.<br>The mean total cost per client of all emergency department visits over the 24-month period was lower for HIP than usual HACC by AU\$22 (ITT) to AU\$67 (AT). Total ED costs for the HIP ITT group were AU\$686 and AU\$659 for the HIP AT group compared with AU\$708 (ITT) to AU\$726 (AT) for the HACC group. |
| Schmidt et al, 2024<br>Australia | **Study design:**<br>Uncontrolled before-after study<br>**Dates of data collection:** January 2018 to March 2023<br>**Data collection methods:**<br>analysis of clinical records<br>**Outcomes of interest:** The aim of the analysis was to describe the population as a whole and sex-based differences, as well as identify whether the program led to reduced disability and improvements to upper and lower limb function and balance.<br>**Outcome measures:** | **Sample size:** 1074<br>**Participants:** Females (n=660), Males (n=414). Aged 65 or over. Mean age 79.<br>**Setting: programme environment =** home (n=1059); residential (n=2); residential and home (n=6)<br>**Type of intervention [exposure]:**<br>Early intervention care which aims to optimise the functioning and independence of older people and reverse and/or slow functional decline. The Short-Term Restorative Care program is a home-based 8-week multidisciplinary intervention led by AHPs. Participants received multiple visits per week (number depending on the participant's goals) from several AHPs. | **Effectiveness:**<br>Findings suggest the program results in statistically significant improvements ($p \leq .0001$) in disability and physical functioning as measured by the Modified Barthel Index, Lower Extremity Functional Scale, Upper Extremity Functional Index and Berg Balance Scale tools. |
|  | functional outcomes derived from program participation as assessed by the Modified Barthel Index (MBI), Lower Extremity Functional Scale (LEFS), Upper Extremity Functional Index (UEFI) and Berg Balance Scale (BBS). | **Length of intervention**<br>8 weeks<br>**Comparator intervention or control**<br>None |  |
| Whitehead et al., 2016<br>England, UK | **Study design:**<br>Feasibility parallel group randomised controlled trial<br>**Dates of data collection:**<br>30 November 2014 to 21 July 2015<br>**Data collection methods:**<br>Questionnaires and screening of recruitment and intervention patient logs.<br>**Outcomes of interest:**<br>Personal and extended Activities of Daily Living (ADLs), health-related and social care-related quality of life at 2 weeks, 3 months and 6 months post reablement. Incidence of falls. Resource use.<br>**Outcome measures:**<br>Barthel Index (BI), Nottingham Extended Activities of Daily Living (NEADL), Short-Form 36 (SF-36) Physical Component Summary (PCS) and Mental Component Summary (MCS), Adult Social Care Outcomes Toolkit (ASCOT), EuroQol EQ-5D-3L. | **Sample size:** n=22 completed 6 month-follow-up.<br>**Participants:** All users of the service within the selected sub teams were screened for eligibility. Inclusion criterion was the ability to provide informed written consent.<br>**Setting:** Home care<br>**Type of intervention [exposure]:** A targeted ADL programme, delivered by an occupational therapist incorporating goal setting, teaching/practising techniques, equipment/adaptations and provision of advice/support. This was in addition to usual care.<br>The service accepted referrals from any adult aged over 18 years, with a need for homecare support except for those with a diagnosis of dementia who already had a specialist dementia homecare service within the area. The service accepted referrals for people being discharged from hospital and those who were living within the community. People leaving hospital with new or increased difficulties with ADL would be particularly likely to be referred to the service | **Effectiveness:** The medians on all measures increased from baseline at all time points in the occupational therapist intervention group, compared to this increase evidenced in only 9 out of 18 participants in the control group. However, it is important to note that the groups were different at baseline, both in relation to their scores on the measures and the preponderance of men in the control group. Change from baseline scores were therefore calculated for each outcome, and a linear regression was carried out in order to adjust for the between-group difference in gender and provide the most accurate estimation of the treatment effect for detection in a powered study. The direction of the change favoured the occupational therapist intervention group in 15/18 measures and time points; however, CIs were wide reflecting the small sample size. Positive trends were particularly evident for social care-related quality of life and mental well-being, which were consistent across all time points. There were fewer falls in the occupational therapist intervention group in terms of the number of participants who reported a fall, and the mean number of falls reported.<br>**Resource use:** At 6 months follow-up, resource use was largely similar between groups. Those in the control group were more likely to use outpatient health services (including GP), an occupational therapist and access meals at home. Those in occupational therapist group were more likely to be admitted to hospital.<br>**Implementation:** Acceptability was evaluated using a questionnaire which was sent to all participants in the intervention group and semi structured interviews with five participants in the intervention group. The questionnaire and interviews revealed a high level of satisfaction |
|  |  | **Comparator intervention or control:**<br>Usual care' was 6 weeks of homecare reablement delivered by social care workers (no routine health professional input). | with the intervention, and participants reported that they believed the intervention helped them to increase their ability to manage ADL. It was possible to deliver the intervention as planned and according to the protocol, and the RTC was feasible. |
| Winkel et al, 2014<br>Denmark | **Study design:**<br>Uncontrolled before-after study<br>**Dates of data collection:**<br>Recruitment took place between October 2010 and July 2011. Final data collection was at 12 week follow up.<br>**Data collection methods:** ADL-Interview at first and third home visit.<br>**Outcomes of interest:** Self-reported ability to perform Activities of Daily Living (ADL). Individualised goals and ADL task performance.<br>**Outcome measures:** ADL-Interview (ADL-I) was conducted at baseline, final home visit and at 12 week follow up. | **Sample size:**<br>No help group n = 32<br>Previous help group n = 59<br>Total N = 91<br>**Participants:** 65 years old citizens. The No Help group had not received home care services within 12 months prior to inclusion in the present study. The Previous help group received home care services at the time of inclusion.<br>**Setting:** Home setting within two local districts of Copenhagen.<br>**Type of intervention [exposure]:** 12-weeks home-based reablement program consisting of ADL task performance training by a home carer supervised by an occupational therapist.<br>**Comparator intervention or control:**<br>Both groups received the same intervention. | **Effectiveness:**<br>ADL ability improved significantly over time ($p = 0.041$). Post-hoc t-tests indicated that the improvements occurred between baseline and end of intervention ($p = 0.042$) and were maintained at follow-up 10 months after intervention ($p = 0.674$). There were no effects related to age ($p = 0.787$) or to whether the older adult had received help previously ($p = 0.120$). Conclusion: A 12-weeks home-based reablement program was found to improve ADL ability among older adults regardless of whether they previously received help. This implies that receiving home care services should not be considered a barrier to participation in a reablement program. |
| Beresford et al, 2019<br>UK | **Study design:**<br>Prospective cohort study<br>Work package 1: Mapped services and developed a typology of service models within 143 sites across England. | **Sample size:**<br>Work package 1:<br>143 services from local authority (53%), NHS (4%), integrated services (15%) or an outsourced provider (14%)<br>Work package 2:<br>N = 186 participants were recruited from three English care reablement services, | **Effectiveness:**<br>In work package 2: There were no statistically significant differences at baseline (T0) in mean outcome scores for the recruited sample and those retained at T1, nor between those referred for support to return home from hospital versus where the referral was to support remaining at home. Those retained at T2 had significantly higher (better) scores on the Barthel Index, Nottingham Extended Activities of Daily Living Scale (NEADL) scale and General Health Questionnaire (GHQ-12) at T0 than the total sample recruited. |
|  | Work package 2: Evaluated three service models, as identified in work package (WP) 1, investigated outcomes, predictors of outcomes, costs, and process evaluation.<br>Work package 3: To investigated current practices regarding reabling people with dementia.<br>**Dates of data collection:** The study was conducted between October 2014 and November 2017.<br>**Data collection methods:** Participants chose whether to self-complete questionnaires, or have measures provided verbally and responses recorded by the researcher. Data were collected on entry to the service, at discharge and 6 months post discharge.<br>**Outcomes of interest:** Healthcare- and social care–related quality of life, functioning, mental health and resource use (service costs, informal carer time, out-of-pocket costs).<br>**Outcome measures:** Health-related quality of life (EQ-5D-5L), Social care-related quality of life (ASCOT SCT-4), Mental health (GHQ-12), Functional status (Barthel Index), Functional ability (Nottingham Extended Activities of Daily Living scale [NEADL]), resource use (Services and Care Pathway Questionnaire [SCPQ]). | each representing a different model of service delivery.<br>n = 128 completed at T1<br>n = 64 completed at T2<br>**Participants:**<br>Individuals returning home from hospital or other inpatient care setting following an acute episode; individuals in whom there is evidence of declining independence or ability to cope with everyday living.<br>**Setting:** Statutorily funded adult social care reablement services located in different regions in England.<br>**Type of intervention [exposure]:**<br>The study defined reablement as intensive, time-limited (up to 6 weeks), individualised and goals-focused and delivered in the usual place of residence.<br>Services identified and assessed:<br>Reablement with occupational therapy (n = 24), homecare reablement (n = 42), multidisciplinary reablement (n = 20), reablement workers (n = 43).<br>The types of reablement inputs: personal care, domestic tasks, safety (including preventing falls and providing aids and equipment), information and signposting to other services or support, getting around in the home, getting out and about outside the home, re-engaging with social activities and friends, managing health-related needs, specific activities to help rebuild confidence and improve well-being.<br>**Comparator intervention or control:**<br>None | For EQ-5D-5L, EQ-VAS and GHQ-12, the difference between T1 and T2 mean scores is smaller than between T0 and T1 but remains in the same direction. For the ASCOT-SCT4 the T2 mean score was slightly lower than the T1 mean score. For the NEADL scale, the size of the difference in mean score was greater between T1 and T2 than T0 and T1 Mean scores at T1 and T2 for Remain at Home and Return Home sub-samples were not significantly different.<br>**Resource use:**<br>Resource use was more frequent before reablement, particularly overnight hospitalisations and care services. Some participants had home adaptations, generally minor. Equipment acquisition was more common, typically before and during reablement. Voluntary service use was very rare throughout the study. Informal care provision was frequent but reduced over time.<br>**Costs:**<br>Work package 1, The response rates to questions on costs were poor. Based on the available data, the cost of reablement per case was calculated to be £1700.<br>In work package 2: Costs of healthcare and social care falling on the public sector were greatest prior to reablement, with a large reduction observed in the cost of hospital overnight stays. Out-of-pocket costs were generally very small throughout the study. Informal care time was a major cost, particularly prior to and during reablement.<br>**Implementation:**<br>Staff frequently reported that service users and family members had a poor understanding of reablement, and this acted as a barrier to engagement.<br>Service users revealed confusion about reablement and its difference from home care and the association between a single versus separate assessor and worker teams<br>Assessors working in a separate team model reported concerns or inadequacies with monitoring/review processes and supervision of reablement workers. |

|  |  |  |
| --- | --- | --- |
|  |  | Staff raised a number of other issues. First, the Care Act 2014 (Great Britain. Care Act 2014. London: The Stationery Office; 2014) and NHS discharge-to-assess policies affected the characteristics of their caseloads. Reabling to full independence was no longer the predominant outcome. The merging of reablement and assessment functions, and the lack of other in-house social care provision for older people, had resulted in increased caseload volume for assessors.<br>There was a strong and consistent belief among staff of the superiority of reablement over traditional home care. Staff skills and knowledge of reablement principles were regarded as key to successful reablement.<br>Staff believed that the social contact occurring during home visits served to reconnect with, and rekindle interest in, everyday life.<br>In addition to engagement, service user characteristics that staff believed had an impact on outcomes included personality, problem-solving skills, mental health, cognitive impairment and the presence of a partner or spouse.<br>Housing tenure was a potential barrier, as installing aids or making minor modifications could be difficult if the property was rented, particularly from a private landlord.<br>Staff believed that other aspects of service delivery and practice had an impact on outcomes. The importance of flexibility in the timing and duration of visits, and an expectation that this would reduce over the course of reablement, was noted. Some believed that this was not sufficiently specified, or incentivised, in contracts with outsourced providers.<br>Views were mixed about the optimum number of workers assigned to a case; advantages of having one or two or, alternatively, multiple workers were articulated. Timely access to specialist expertise, particularly physiotherapy and mental health, was regarded as important but difficult, if not impossible, to achieve.<br>Service users believed that positive outcomes had been achieved through the skills and input of the reablement workers. Six factors |
|  |  |  | were articulated as having had an impact on outcomes: the service user–worker relationship, the workers’ reablement skills, the service user’s confidence in the worker, the duration of home visits, the service user’s willingness to accept support, and the service user being able to review progress.<br>Continuity of workers was not identified as important to achieving positive outcomes. |
| Tinetti et al, 2012<br>USA | **Study design:**<br>Non-randomised controlled trial<br>**Dates of data collection:**<br>Nonhospice Medicare-covered home care between November 1, 1998, and April 30, 2000<br>**Data collection methods:**<br>Outcome and Assessment Information Set (OASIS-B), a self-care progress report, in which each ADL was divided into subtasks, helped to clarify goals, establish a baseline, standardise assessment, clarify care responsibility of providers, and track progress.<br>**Outcomes of interest:** Hospital readmission and length of home care episode.<br>**Outcome measures:**<br>Outcome and Assessment Information Set (OASIS) version B. OASIS must be completed on admission to home healthcare services, transfer to another site of care (e.g., hospital), resumption of care (after a hospital stay), follow-up (at least 60 days), discharge from home health care, and death. Outcome of the episode of home care was dichotomised as | **Sample size:** 770 (341 matched pairs and 88 additional participants included in the unmatched analysis but whose match died or had missing outcome data).<br>**Participants:** Individuals aged 65 and over receiving care from a large home care agency after hospitalisation.<br>**Setting:** Homecare<br>**Type of intervention [exposure]:**<br>Restorative care: The restorative care model was based on principles adapted from geriatric medicine, nursing, rehabilitation, goal attainment, chronic care management, and behavioural change theory. A multidisciplinary work group of expert home care clinicians familiarised staff with the goal of optimising patient function, developed methods to assess ADLs and quantify increments of improvement, and focused staff on developing individual functional goals with patients and linking interventions to patients’ goals.<br>**Comparator intervention or control:**<br>Usual care: No attempt was made to change home care practices in the usual care offices. Although the nurses and therapists made efforts to communicate regularly, their care plans were developed independently. The HHAs performed or | **Resource use:**<br>13.2% of restorative model participants were readmitted to an acute hospital during the episode of home care, versus 17.6% of usual care participants. According to the conditional regression analysis, participants receiving the restorative model of home care were 32% less likely to be readmitted than those receiving usual care (odds ratio (OR) = 0.68, 95% confidence interval (CI) = 0.43–1.08). The mean length of home care episodes was 20.3 ± 14.8 days in the restorative care group and 29.1 ± 31.7 days in the usual care group (P<0.001). Results were similar in unmatched analyses. |
|  | remaining at home or readmission to an acute hospital. | helped the participant perform basic and instrumental ADLs. |  |
| Tinetti et al, 2002<br>USA | **Study design:**<br>Non-randomised controlled trial<br>**Dates of data collection:**<br>Individual matching was conducted between November 1, 1998, and April 30, 2000.<br>**Data collection methods:**<br>All descriptive and outcome data were ascertained using the OASIS-B completed by a nurse or a therapist at the time of admission to home care, every 60 days and at discharge from home care.<br>A self-care progress report helped to clarify goals, establish a baseline, standardize the assessment of patients, clarify care responsibility across the multiple providers and track patients incremental progress towards reaching those goals.<br>**Outcomes of interest:**<br>Remaining at home, emergency department visits, home care service use and ADLs (mobility, home management and self care).<br>**Outcome measures:**<br>The duration of home care was based on the start and end dates recorded in the OASIS-B database whole intensity was based on the | **Sample size:** 691 matched pairs<br>**Participants:** Patients receiving home care through the restorative care office who were 65 years or older, in receipt of Medicare-covered home care lasting at least 7 days; with absence of severe cognitive impairment; and not terminal, bedridden, or requiring total care were matched with patients from 1 of the usual care offices.<br>**Setting:** Home care agency<br>**Type of intervention [exposure]:**<br>Restorative care: all care was provided by the home care agency's nursing and physical and occupational therapy, and home health aide staff. The restorative care model was based on the following principals: nursing, rehabilitation and goal attainment. The treatment plan included various combination of exercise and training; behavioural changes; environmental adjustments and adaptive equipment; counselling and support; training and education of patient, family and friends and medication adjustments.<br>**Comparator intervention or control:** No attempt was made to change home care practices in the usual care offices. The care of patients in these offices was directed by the medical orders developed to manage specific diseases. While the nurses and therapists made efforts to | **Effectiveness:**<br>Intervention had significantly higher ($p=0.02$) mean mobility scores and higher mean self-care ADLs compared to usual ($p=0.07$).<br>**Resource use:**<br>Compared with usual care, and after adjusting for baseline characteristics and other factors, restorative care was associated with a greater likelihood of remaining at home (82% vs 71%) and a reduced likelihood of visiting an emergency department (10% vs 20%). Home care episodes were shorter (mean [SD], 24.8 [26.8] days vs 34.3 [44.2] days). Restorative care patients had better mean (SD) scores than usual care patients in self-care (11.0 [2.1] vs 10.7 [2.5]; $P=.07$ after adjustment), home management (9.5 [2.9] vs 9.2 [3.0]; $P=.05$ after adjustment), and mobility (3.3 [0.8] vs 33.2 [0.9]; $P=.02$ after adjustment). |
|  | number of nursing, physical therapy, and home health aide visits recording into the billing files. For individuals who remained at home after home care discharge, additional primary outcomes included self-care and home management, ADLs, and mobility. | communicate regularly, at least by telephone, their care plans (including direction other home health aides) were developed independently. |  |
| Glendinning et al., 2010<br>England | **Study design: Non-randomised controlled trial**<br>**Dates of data collection:** Recruitment of service users commenced in November 2008 and lasted until July 2009. 12-month follow-up continued until May 2010.<br>**Data collection methods:** Both quantitative and qualitative data were collected from service users, carers and local authority staff in the participating sites. Collected through validated and bespoke questionnaires, interviews, focus groups, observations and screening of health records.<br>**Outcomes of interest:** Self-perceived health, perceived quality of life, health-related quality of life, social care outcomes, social care service use, NHS service use, third sector service use, costs of service use and implementation.<br>**Outcome measures:** Self-perceived health (a five-point scale), Perceived quality of life (a seven-point scale), Health-related quality of life (EQ-5D), Social care | **Sample size:** N=1015 recruited. Complete cases at 12-month follow-up were n=241 in reablement group and n=141 in comparison group.<br>**Participants:** Study participants were recruited from five councils that offered home care reablement services and five comparison group sites where conventional home care services were available.<br>**Setting:** Local authority reablement and conventional home-care services.<br>**Type of intervention [exposure]:** Five separate home reablement services provided by local councils. Each service model differed slightly and are described below.<br>*Community response service:* operated by team leaders managing day-to-day operations and setting goals with clients, community response assistants provided support in complicated cases. No therapists were part of the reablement service. The service runs for six weeks (can extend to 8 if necessary). The service is available to all adults requiring home care, aged 18 years and over. The service does not take people with learning disabilities as they are considered to need a long-term support package. The service supports predominantly older people with | **Effectiveness:**<br>**Health-related quality of life** (measured by EQ-5D) evidenced the reablement group had statistically significantly better mobility than the comparison group at baseline; a lower percentage of people unable to self-care but also fewer with no self-care problems; fewer people with moderate or extreme anxiety; but more people who felt their general health was worse than a year ago. At follow-up, the reablement group had statistically significantly better health-related quality of life on all five dimensions and general health compared with the comparison group. The greatest difference was in the ability to perform usual activities where 23% in the reablement group were unable to perform usual activities compared to 43% in the comparison group. When presented as overall score the reablement group had a statistically significant improvement in health-related quality of life between baseline (mean 0.38, sd 0.30) and 12-month follow-up (mean 0.47, sd 0.35, $p<0.001$). There was no change in the comparison group (baseline mean 0.33 compared to follow-up mean 0.32).<br>**Social care-related quality of life** (as measured by the ASCOT tool) showed that at baseline, the reablement group had statistically significantly higher needs in undertaking usual activities than the comparison group, but had fewer needs in relation to safety, social situations and relationships, and dignity and respect. At 12-month follow-up, there were no statistically significant differences in social care-related quality of life between the reablement and comparison groups. There was no change in mean social care need scores (calculated ASCOT scores) between baseline and 12-month follow-up in either the reablement or comparison groups. In the reablement group, the mean score rose slightly (but not statistically significantly) from 0.777 at baseline to 0.78 at follow-up. In the comparison group, the mean score was 0.80 at baseline and 0.76 at 12-month follow-up. The difference between the reablement and comparison groups at |
|  | outcomes (ASCOT – Adult Social Care Outcomes Toolkit), national unit costs, health and social care resource use questionnaires. | physical or dementia needs. It also offers support to younger people with newly diagnosed conditions such as MS, Parkinson's and motor neuron disease. The service takes referrals from hospital and community teams.<br>*Home Care Rapid Response Team:* The reablement service is comprised of two sections: Rapid Response (which does the assessments) and the provider side. There are six managers who do assessments. Within the providers, there are East and West teams. Each team consists of a line manager (responsible for planning care, managing staff, liaising with service users) leaders (supporting staff and setting up the packages of care) and carers.<br>There are no occupational therapists and physiotherapists in the teams, but they work closely with the therapy team and social workers. The aim of the service is to enable adults/older people to retain/regain their independence to facilitate early hospital discharge; prevent hospital admissions where medically safe to be treated at home; and reduce premature admission to long-term residential or nursing care. Reablement is offered within the home care service to service users identified as having the potential to become independent in at least one area of activity. The service provides support for six weeks (can be extended by two weeks, if necessary). Most referrals are of older people, an increasing number of whom may have dementia or other mental health problems. The service is available to all adults requiring home care, aged 18 years and over. The service accepts referrals from all routes. However, the majority of referrals to Rapid Response | baseline and at 12-month follow-up did not reach statistical significance.<br>Overall, the use of reablement was significantly associated with better EQ-5D and ASCOT outcomes than the use of conventional home care services in the analysis. In other words, the analysis indicates that with a probability of over 95% the use of reablement leads to improved outcomes for service users in terms of health-related quality of life (EQ-5D). While the use of reablement was associated with better social care-related quality of life (ASCOT), the impact was not as strong as for the EQ-5D outcome measure.<br>**Activities of daily living:** Higher dependency level at baseline among people in the comparison group was also reflected in the significantly higher proportion unable to perform some activities of daily living independently compared with those in the reablement group. A greater percentage of people in the comparison group than the reablement group was unable at baseline to undertake all activities except being able to get outdoors and walk down the road. The greatest statistically significant differences were the higher percentages of people in the comparison group unable to get in or out of a bed or chair (19% control, 10% reablement group), and unable to wash their hands and face (16% control, 8% reablement group). By the time of the 12-month follow-up, people in the comparison group were still less able than the reablement group to undertake all activities of daily living; the greatest statistically significant differences between the groups had changed to being unable to get outdoors and walk down the road (65% control, 42% reablement group), and unable to bath, shower or wash all over (65% control, 38% reablement group).<br>**Resource use: Social care services:** Only six people in the comparison group and none in the reablement group used day care services in the 10-month follow-up period. For social care services other than meals on wheels, more people receiving conventional home care services were in receipt of each service compared with those in the reablement group. During the initial eight-week period, 12 per cent of people in the reablement group received meals on wheels compared to five per cent in the comparison group. The percentage of people receiving independent home care was lower in the reablement than the comparison group both within the first eight weeks (2% and 65% respectively) and the remaining ten months |
|  |  | come from the hospital, GPs, Community Matrons, and only occasionally from the care management team.<br>*Homecare Assessment and Re-ablement Team (HART):* There are ten HART teams. Each team includes a homecare manager (overall responsibility for all cases, reviews and drawing up support plans for users needing ongoing home care services on discharge from HART); senior homecare assistants (responsible for observing/ monitoring Homecare Assistants, contributing to risk assessment work, supporting Homecare Assistants in more complex cases and assessing for minor adaptations); homecare assistants and clerical support.<br>HART has two main roles: assessment and reablement. All new referrals for homecare are referred to the HART team following a community care assessment by a care manager or from health via the Single Assessment Process. If there is no capacity within the team to support the service user, or if it is considered that they would not benefit from a short-term reablement intervention, they are referred straight to an independent sector home care provider. If the case is accepted by HART, the team provides support for up to six weeks (can be extended by one to two weeks if necessary). If the service user needs on-going care at the end of the period of reablement, the HART manager will commission home care from independent agencies. The HART service is available to all adults aged 18 and over assessed as needing home care support, including people with learning difficulties and older people with dementia. The only exception is when it is considered that a short-term, reablement focused | (18% and 50% respectively). Data on proportions of NHS healthcare contacts not clearly presented in this study.<br>**Costs:** Mean costs for social care service use and NHS healthcare costs were presented for the initial eight-week service provision period as well as the ten-month follow-up period up to the 12-month end follow-up. Costs of eight-week care provision are only considered here.<br>**NHS health service use costs** for both groups during the eight-week period of provision were significantly higher in the re-ablement group (£1,600) compared to control (£1,095) [p<0.05]. However, there was not a significant difference in the cost of health care services during the following 10 months after receiving the home care re-ablement services (£3,455 for reablement group, £3,235 for control, not significant). In terms of mean costs for each health care service over the 10-month follow-up period, the costs for hospital inpatient stays (£1,445 for reablement group, £970 for control), visits to an outpatient department (£539 for reablement group, £678 for control), and visits to the nurse (£533 for reablement group, £548 for control), GP (£685 for reablement, £650 for control), accident and emergency (£101 for reablement, £154 for control) and therapist (£124 for reablement, £146 for control) were similar but typically evidenced greater mean costs in the reablement group than control. During both the initial eight weeks and following 10 months, people in the comparison group had significantly higher chiropody costs than those in the re-ablement group. There was no statistically significant difference in health care costs between the two study groups over the whole 12-month period.<br>**Social care service use costs** for both groups during the eight-week period of provision were significantly higher in the re-ablement group (£1,640) compared to control (£570) [p<0.001]. The difference in costs was explained by the reablement services cost (£1,540) in the reablement group and independent home care costs in the control group (£510). During the 10-months following the reablement service provision, social care costs were significantly higher for people receiving conventional home care services (mean £2,240) compared with those in the reablement group (£790; p<0.001). For each element of social care services over the 10-month period, mean costs were greater across all elements in the control group: in-house home care (£270 reablement group, £590 control), independent home care |
|  |  | intervention would either be detrimental to the person or that the service user would not benefit from it in any way, e.g. somebody who is in the last stages of a terminal illness. The service takes referrals from hospital and community teams.<br>*START Service:* The Team Manager is responsible for the overall operational performance of the home care service, which includes START, long-term clients and extra care housing. There are seven teams, supporting Rapid Response and START clients: these include managers (overseeing the whole service, supporting case management, risk management issues, line management of staff, supervision, performance observation of staff); a scheduling team (allocating work to home care staff), senior care officers (overseeing cases, supervising staff, risk management), care officers (Rapid Responders) and home care assistants. The service is supporting individuals to regain their levels of independence and maximise their daily living skills. When there is no capacity to take on a referral, the referral will go back to the commissioning teams who will arrange care from independent providers. The START Service provides support for six weeks (extended by two weeks, if necessary). If the service user needs on-going support, the care management teams are responsible for commissioning the ongoing support. The START Service is available to all people over 18 who are assessed as needing home care services (including people with mental health problems and dementia but excluding people with learning disabilities). The service takes referrals from hospital and community teams. | (£450 reablement group, £1,660 control group), day care (£0 reablement group, £60 control) and meals on wheels (£60 reablement group, £70 control).<br>**Implementation:**<br>One factor impacting on the effectiveness of reablement was who the service was offered to. Managers and front-line staff agreed that service users likely to show greatest benefit were those recovering from falls or fractures. Unlike conventional home care, reablement reduced the likelihood of this group of people becoming used to having tasks done for them and thus helped avoid creating dependency. Reablement was considered less likely to produce significant results for service users such as people with dementia or other mental health problems who tended to need on-going support. This did not mean that no benefits could be achieved for these latter service users; rather, that it was much harder to achieve major improvements in a short period of time. However, reablement could more accurately identify the scale and of the long-term support they needed. Moreover, by focusing on small targets and establishing a sense of routine, reablement could raise their confidence and perhaps eventually reduce their use of services. Service user's motivation was considered to be another factor influencing effectiveness.<br>The commitment, enthusiasm, knowledge and skills of reablement front-line staff were highlighted by both managers and other staff as the most important factors in the success of home care reablement. The ideal reablement worker had a good understanding of the concept and practice of reablement, with the skills to motivate and encourage service users to become as independent as possible. These skills included observational skills and the ability to stand back and assess service user's potential in various aspects of their lives. Other essential skills included working closely with the service users to identify the support they needed to reach their potential, building up service users' confidence and convincing them that they could achieve more independence. All participants emphasised the importance of training and on-going supervision to reinforce the reablement approach in day-to-day practice.<br>All managers highlighted the importance of service users being involved in goal setting. While front-line staff were not involved in |
|  |  | *Homecare Assessment and Reablement Team (HART):* There are 30 HART teams (ten in each of three areas), including registered managers (managing the organisers), organisers (line managing enablers, organising programmes/workloads, doing risk assessments and organising discharges), senior carers (taking on any complicated cases, doing quality assurance visits, supervising enablers), home care enablers, community occupational therapists, funded by social services and taking care of community referrals. Additional occupational therapists are embedded in HART with different grades and funded by the Acute Hospital Trust.<br>HART has two main roles: assessment and reablement. HART provides support for up to six weeks (can be extended for a week if necessary). If service users need on-going support, the HART manager will commission that from an independent agency. The HART service is available to all adults aged 18 and over who live in the local authority area. The only limiting factor is whether HART has the capacity to meet service users' needs, for example, people needing complex and/or continuing care packages. HART also excludes people in the last stages of a terminal illness who they consider would not benefit from short term interventions. The service takes referrals from hospital and community teams.<br>**Comparator intervention or control:**<br>Control defined in this study as conventional home-care services (home-care where reablement specific care is not provided). | those initial goal setting tasks, all the managers said the service relied on regular staff feedback. In addition, all managers noted that more formal reviews took place at different points within the reablement period to further assess the service user's progress.<br>The importance of the assessment on first referral to reablement was highlighted by managers as a means of accurately capturing changing needs, particularly in the transition from hospital to home. Once back in the home environment, it was possible that people may be able to do some things they could not do in hospital, but may identify other areas where they need support to live independently, such as help with shopping and/or social activities. Assessment could be as much about reducing the level of support as about increasing it.<br>Two managers were particularly concerned about the poor quality of assessments carried out prior to discharge from hospital: hospital staff made no home visits and relied on self-reports from service users. This created difficulties for the reablement team as they were under pressure to get people out of hospital and set up appropriate support arrangements. Several senior practitioners said that their assessments could result in re-hospitalisations, for example when service users were unable to walk and had not been given any equipment on discharge.<br>Unlike conventional home care services, all managers reported greater flexibility in the length of time for individual visits, which they thought was crucial to the effectiveness of reablement. Reablement staff worked in small teams so that any one service user saw a limited number of workers. Minimising the number of carers working with one person was important both for the quality of the service and for effective communication about changes in support needs, goals and abilities as user's abilities improved. The number of carers visiting individual service users depended on the frequency and length of visits, the overall support package and how staff rotas were arranged in the site.<br>Two issues were reported to affect the discharge process. One was the ability of the reablement team to commission on-going home care services and other support themselves, rather than having to refer service users back to care managers. The second factor affecting<br>discharge was under-capacity within independent home care services. |

**Table 10.** Summary of economic evaluation studies assessing cost-effectiveness.

| Citation (Country of study) | Study characteristics and health economics methods | Outcomes and costs measured | Main health economics findings |
| --- | --- | --- | --- |
| Kjerstad and Tuntland, 2016<br>Norway | **Aim:**<br>To assess the cost-effectiveness of a reablement programme in Norway.<br>**Type of intervention:**<br>The intervention group received ten weeks of multicomponent home-based rehabilitation. The therapeutic process is tailored according to the needs and aims of the individual participant, resulting in differences in the number and type of elements in the intervention across participants.<br>**Sample size**<br>N=46 (25 participants in the intervention group and 21 participants in the control group).<br>**Length of follow-up**<br>3 months and 9 months.<br>**Analytical approach (e.g. trial-based or model-based).**<br>Trial-based (RCT)<br>**Type of economic evaluation**<br>Cost-effectiveness analysis | **Outcomes**<br>The economic evaluation was based on outcomes from the RCT (Tuntland et al., 2015). The outcome measure used in the cost-effectiveness analysis was the Canadian Occupational Performance Measure (COPM) was used to measure self-perceived activity performance and satisfaction with performance.<br>**Costs**<br>The cost data used in this study were based on registrations of the number of home visits to each individual participant; the duration of the visits; the type of professional delivering the care and wage costs differentiated according to profession. | **Cost-effectiveness**<br>In summary, reablement was found to be more cost-effective than usual care. The assessments of performance and satisfaction regarding daily activities were significantly higher in the reablement group compared with the control group and this was achieved at lower cost.<br>The incremental cost-effectiveness ratio (ICER) reported that the cost per one-point improvement in the COPM-performance was 868.18 NOK lower on average for the intervention group compared with the control group. The cost per one-point improvement in the COPM-satisfaction was 666.30 NOK lower on average for the intervention group compared with the control group.<br>**Intervention delivery costs**<br>The significant difference in costs per visit was rather modest though (NOK 9.61), so, in total, the aggregate cost during the intervention period was approximately 12,000 NOK lower for the intervention group. The lower number of visits received by the intervention group, a somewhat unexpected result, explains the cost difference. The difference in mean cost per participant between the two groups was not significant based on per participant estimations.<br>At 3 month follow-up, the difference in mean costs per participant was approximately 1,134 NOK, that is, the mean |
|  | **Perspective of analysis**<br>Not stated<br>**Currency and cost year**<br>Norwegian kroner (NOK), cost year not stated<br>**Discounting**<br>No discounting as time horizon was less than 12 months. |  | costs per participant were lower for the intervention group compared with the control group at 3 months follow up.<br>**Resource use costs**<br>Wider resource use costs were assessed beyond the interventions period between 3 months and 9 months follow-up to explore long-term expenditure on home-based care services after the trial had ended. The intervention group received on average 88 home visits compared with the control group receiving 158 visits on average, a difference of 78 visits on average. This equated to an aggregate difference in total expenditure in the 6-month period of approximately 130,000 NOK. The mean cost per person in the intervention group was 6,470 NOK, while the average in the control group was 13,914 NOK. |
| Glendinning et al., 2010<br>England, UK | **Aim**<br>Identify the effectiveness and cost-effectiveness of home care re-ablement services compared to traditional home care services. Identify any impact on and savings in the use of social care and other services that can be set against the costs of re-ablement services.<br>**Intervention type**<br>Five separate home re-ablement services provided by local councils. Each service model differed slightly and are described below.<br>*Community response service*: operated by team leaders managing day-to-day operations and setting goals with clients, community response assistants provided support in complicated cases. No therapists were part of the re-ablement service. The service runs for six weeks (can extend to 8 if necessary). The service is available to all adults requiring home care, aged 18 years and over. The service does not take people with learning disabilities as they are considered to need a long-term support package. The service supports predominantly | **Outcomes**<br>Individual outcomes (self-perceived health, perceived quality of life, health-related quality of life and social care outcomes), social care service use (details of all social care services used by trial participants completed by care providers), NHS service and voluntary organisations service use (self-reported service use).<br>**Costs**<br>Unit costs of re-ablement services and subsequent health and social care resource following discharge from home re-ablement service (Client Service Receipt Inventory completed by care providers). | **Cost-effectiveness**<br>Cost-effectiveness was based on health-related quality of life (EQ-5D) and social care-related quality of life (ASCOT). At a threshold of £30,000 per EQ-5D outcome gain, there was a 99% probability of re-ablement being cost-effective when both health and social care costs were included and just under 100 per cent if social care costs only were included. At a more stringent threshold of £20,000 per EQ-5D outcome gain, in the case of health and social care costs the probability of cost-effectiveness fell to around 98% but was still over 99% when only social care costs were considered.<br>In terms of social care outcomes (ASCOT) at a threshold of £30,000 per gain, the probability of re-ablement being cost-effective was 78% if health and social care costs were included and rose to 98 per cent when social care costs only were included. At a more stringent threshold of £20,000 per ASCOT outcome gain, in the case of health and social care costs, the probability of re-ablement being cost-effective was 68 per cent, but still 98 per cent when only social care costs were included. Taking a social care perspective, as the probability reached above 95 per cent at both the £30,000 and £20,000 thresholds, we can conclude with a high statistical probability that re-ablement is cost-effective on the basis of data in this study. Taking a wider perspective that includes health care costs, at a |
|  | older people with physical or dementia needs. It also offers support to younger people with newly diagnosed conditions such as MS, Parkinson's and motor neuron disease. The service takes referrals from hospital and community teams.<br>*Home Care Rapid Response Team:* The re-ablement service is comprised of two sections: Rapid Response (which does the assessments) and the provider side. There are six managers who do assessments. Within the providers, there are East and West teams. Each team consists of a line manager (responsible for planning care, managing staff, liaising with service users) leaders (supporting staff and setting up the packages of care); carers. There are no occupational therapists and physiotherapists in the teams but they work closely with the therapy team and social workers.<br>The aim of the service is to enable adults/older people to retain/regain their independence to facilitate early hospital discharge; prevent hospital admissions where medically safe to be treated at home; and reduce premature admission to long-term residential or nursing care. Re-ablement is offered within the home care service to service users identified as having the potential to become independent in at least one area of activity. The service provides support for six weeks (can be extended by two weeks, if necessary). Most referrals are of older people, an increasing number of whom may have dementia or other mental health problems. The service is available to all adults requiring home care, aged 18 years and over. The service accepts referrals from all routes. However, the<br>majority of referrals to Rapid Response come from the hospital, GPs, Community Matrons, and only occasionally from the care management team.<br>*Homecare Assessment and Re-ablement Team (HART):* There are ten HART teams. Each team includes a homecare manager (overall responsibility for all cases, reviews and drawing up support plans for users needing ongoing home care services on discharge from HART); senior homecare assistants (responsible for observing/monitoring Homecare Assistants, contributing to risk assessment work, supporting Homecare Assistants in more complex cases and assessing for minor adaptations); homecare assistants and clerical support.<br>HART has two main roles: assessment and re-ablement. All new referrals for homecare are referred to the HART team following a community care assessment by a care manager or from health via the Single Assessment Process. If there is no capacity within the team to support the service user, or if it is considered that they would not benefit from a short-term re-ablement intervention, they are referred straight to an independent sector home care provider. If the case is accepted by HART, the team provides support for up to six weeks (can be extended by one to two weeks if necessary). If the service user needs ongoing care at the end of the period of re-ablement, the HART manager will commission home care from independent agencies. The HART service is available to all adults aged 18 and over assessed as needing home care support, including people with learning difficulties and older people with dementia. The only exception is when it is considered that a short-term, re-ablement<br>focused intervention would either be detrimental to the person or that the service user would not benefit from it in any way, e.g. somebody who is in the last stages of a terminal illness. The service takes referrals from hospital and community teams.<br>*START Service:* The Team Manager is responsible for the overall operational performance of the home care service, which includes START, long-term clients and extra care housing. There are seven teams, supporting Rapid Response and START clients: these include managers (overseeing the whole service, supporting case management, risk management issues, line management of staff, supervision, performance observation of staff); a scheduling team (allocating work to home care staff), senior care officers (overseeing cases, supervising staff, risk management), care officers (Rapid Responders) and home care assistants.<br>The service is supporting individuals to regain their levels of independence and maximise their daily living skills. When there is no capacity to take on a referral, the referral will go back to the commissioning teams who will arrange care from independent providers. The START Service provides support for six weeks (extended by two weeks, if necessary). If the service user needs on-going support, the care management teams are responsible for commissioning the ongoing support. The START Service is available to all people over 18 who are assessed as needing home care services (including people with mental health problems and dementia but excluding people with learning disabilities). The service takes referrals from hospital and community teams. |  | threshold of £30,000 per EQ-5D outcome gain, re-ablement was cost-effective. However, it was less certain for ASCOT outcome gain, but re-ablement is more likely to be cost-effective than not cost-effective.<br>**Resource use costs**<br>Taking total healthcare, social care and re-ablement costs together, there was no statistically significant difference in the costs of all the services used by the re-ablement and comparison group over the 12 month study period.<br>Re-ablement was associated with a significant decrease in subsequent social care service use. The costs of the social care services used by people in the reablement group during the 12 months of the study (excluding the costs of the reablement intervention itself) were 60% less than the costs of the social care services used by people using conventional home care services.<br>However, this reduction in social care costs was almost entirely offset by the initial cost of the re-ablement intervention. The total (including re-ablement) mean cost of the social care services used by the re-ablement group was £380 lower than the total mean cost of the social care services used by the comparison group. |
|  | *Homecare Assessment and Re-ablement Team (HART):* There are 30 HART teams (ten in each of three areas), including registered managers (managing the organisers), organisers (line managing enablers, organising programmes/workloads, doing risk assessments and organising discharges), senior carers (taking on any complicated cases, doing quality assurance visits, supervising enablers), home care enablers, community occupational therapists, funded by social services and taking care of community referrals. Additional occupational therapists are embedded in HART with different grades and funded by the Acute Hospital Trust.<br>HART has two main roles: assessment and re-ablement. HART provides support for up to six weeks (can be extended for a week if necessary). If service users need on-going support, the HART manager will commission that from an independent agency. The HART service is available to all adults aged 18 and over who live in the local authority area. The only limiting factor is whether HART has the capacity to meet service users' needs, for example, people needing complex and/or continuing care packages. HART also excludes people in the last stages of a terminal illness who they consider would not benefit from short term interventions. The service takes referrals from hospital and community teams.<br>**Sample size**<br>N=1015 recruited, n=381 completed baseline, post-intervention and follow-up). (n=241 intervention group, n=141 control group).<br>**Length of follow-up**<br>9 to 12 months follow-up |  |  |
|  | **Analytical approach (e.g. trial-based or model-based).**<br>Trial-based economic evaluation<br>**Type of economic evaluation**<br>Cost-effectiveness analysis<br>**Perspective of analysis**<br>NHS and social care perspective, wider societal perspective<br>**Currency and cost year**<br>GBP (£) 2009/10 cost year<br>**Discounting**<br>Not conducted as follow-up 1 year<br>**Sensitivity analysis**<br>The probability that reablement is cost-effective when re-ablement health care costs are between 5% and 25% higher than reported, and where health care costs for the comparison group are unchanged were 98.1% and 89.3% respectively. These scenarios were chosen because they will reduce the probability that re-ablement is cost-effective and therefore challenge the main conclusions. Developing a cost-effectiveness acceptability curve, In this case, at a threshold of £30,000 per ASCOT gained, for social care costs, re-ablement is cost-effective with a 98% probability. When all costs are included, the probability falls to 78%. At a threshold of £20,000 per ASCOT outcome gain, the probability of cost-effectiveness is 98% against social care costs and 68% against all costs. |  |  |
| Bauer et al., 2018<br>England, UK | **Aim**<br>To examine whether home-care reablement schemes for older people at home have the potential to reduce costs from the perspective | **Outcomes**<br>Outcomes used in development of decision analytic model comparing reablement and standard care included: relative risk of hospital | **Cost-effectiveness**<br>Not reported in this analysis, costs only reported as cost-minimisation analysis. Cost-minimisation typically assumes equivalence of outcomes but this wasn't stated. |
|  | of the National Health Service (NHS) and Personal Social Services (PSS) in England.<br>**Intervention type**<br>Home-care reablement schemes for older people at home with the aim of improving outcomes and reducing costs.<br>**Sample size**<br>Two synthetic cohorts of n=1000 each using previously published data.<br>**Length of follow-up**<br>Unclear, but model cycles ran for 1 year each time.<br>**Analytical approach (e.g. trial-based or model-based).**<br>Model-based approach using a decision-analytic Markov model.<br>**Type of economic evaluation**<br>Cost-minimisation analysis<br>**Perspective of analysis**<br>National Health Service (NHS) and Personal Social Services (PSS) perspectives.<br>**Currency and cost year**<br>GBP (£) 2014/15 cost year<br>**Discounting**<br>An annual discount rate of 3.5% was applied to costs occurring after the first year.<br>**Sensitivity analysis**<br>Findings were robust to changes in different unit cost estimates.<br>For example, in a scenario in which the cost of reablement increased by 50%, mean potential cost savings remained as high<br>as almost £1,500. Even when costs of home care, hospital, and reablement were simultaneously increased or decreased by 20%, the lowest estimate for cost reductions was £1,109 and the lowest probability that potential cost reductions would occur was 87.9%. If the assumed effect of reablement was changed from 50% to 40% of the figures reported by Lewin et al. (2013, 2014) then the mean net benefit reduced from £2,061 to £1,581; if it was changed from 50% to 60% of those figures, the mean net benefit increased to £2,541. | admission (0.99 at 12 months, 0.92 at 24 months), probabilities of hospital admission in reablement during the first year (those using home care: 60.5%, not using home care: 52.5%), probabilities of hospital admission in reablement during the second year (those using home care: 56.4%, not using home care: 48.9%), relative risk of ongoing home care (at 3 months: 0.66, at 12 months: 0.68) and probabilities of ongoing home care (64.9%).<br>**Costs**<br>Costs of reablement were calculated from different sources as there was no single source that provided a national average. Instead, mean, lower, and upper bound values were derived from various sources. A first estimate was based on the costs of providing reablement evaluated by Glendinning et al. (2010) for five sites in England. Values, uprated to 2014/15 prices with the Hospital and Community Health Services (HCHS) pay and price index, included a lowest cost of £1,756, a highest cost of £3,901, and mean cost of £2,279. This referred to a mean duration of reablement of 39 days. A second estimate was taken from the National Audit Report for Intermediate Care (NAIC, 2015) which provided the mean cost per person in reablement schemes as £1,484. This was based on annual service budgets of the commissioners and providers that voluntarily participated in the Audit. | **Intervention delivery costs**<br>Used as model parameters and identified from previous studies on the topic.<br>**Resource use costs**<br>Mean cost in the reablement group was £56,499 per person, ranging from £55,690 to £57,307 at a 95% confidence interval (CI). Mean cost in the standard care group was £58,560 (95% CI 57,800 to 59,319). The mean difference was -£2,061 (95% CI 1,933 to 2,129). In both groups, most of the total costs (over 80%) were social care related: If reablement was counted as a cost to local authorities, mean costs to this sector were £46,894 in the reablement group and £48,887 in the standard care group. On the same basis, 97% (£1,993) of the net benefit was due to reductions in costs to local authorities, and only 3% (£68) was due to reductions in costs to the NHS.<br>Costs in both reablement and standard care groups decreased with higher starting ages of cohorts, mean differences increased to -£3,244 (starting age 75) and -£3,844 (starting age 85). The decrease in costs was explained by the overall shorter period in which costs occurred, and the increase in mean differences reflected the relatively greater impact on hospital admission rates for older age groups (who have higher risks of hospital admission). The probability that reablement was cost-reducing increased to 99.9% (for starting age 75) and 100% (for starting age 85). |

### 7.3 Quality appraisal

The summary tables for the quality appraisals are presented in section 9.2.

### 7.4 Information available on request

The data that supports the findings of this study are available in the data extraction tables of this report. The search strategies conducted in the databases are available in section 9.1.

## Acknowledgements

The authors would like to thank Denise Moultrie, Ruth Crowder, Kerrie Phipps, Emily Clarke, Rhian Matthews and Rashmi Kumar for their time, expertise, and contributions during stakeholder meetings in guiding the focus of the review and interpretation of findings. We would also like to thank Elizabeth Gillen and Juliet Hounsome, Information Specialists at Cardiff University, for their expert contribution to the search strategy development and database searches.

## APPENDICES

### 9.1 Search strategies conducted in databases

Database: Ovid MEDLINE(R) ALL <1946 to January 14, 2025>

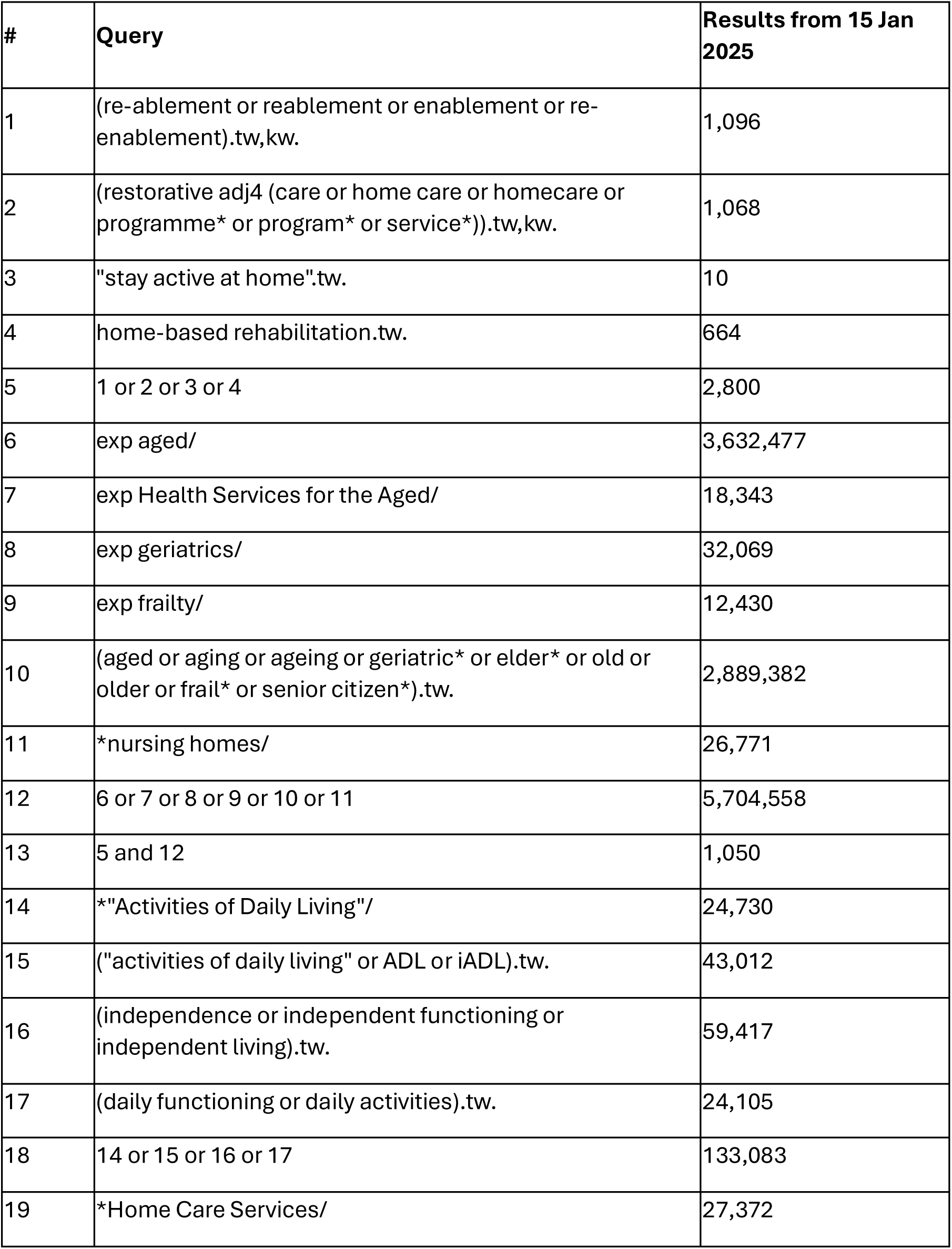
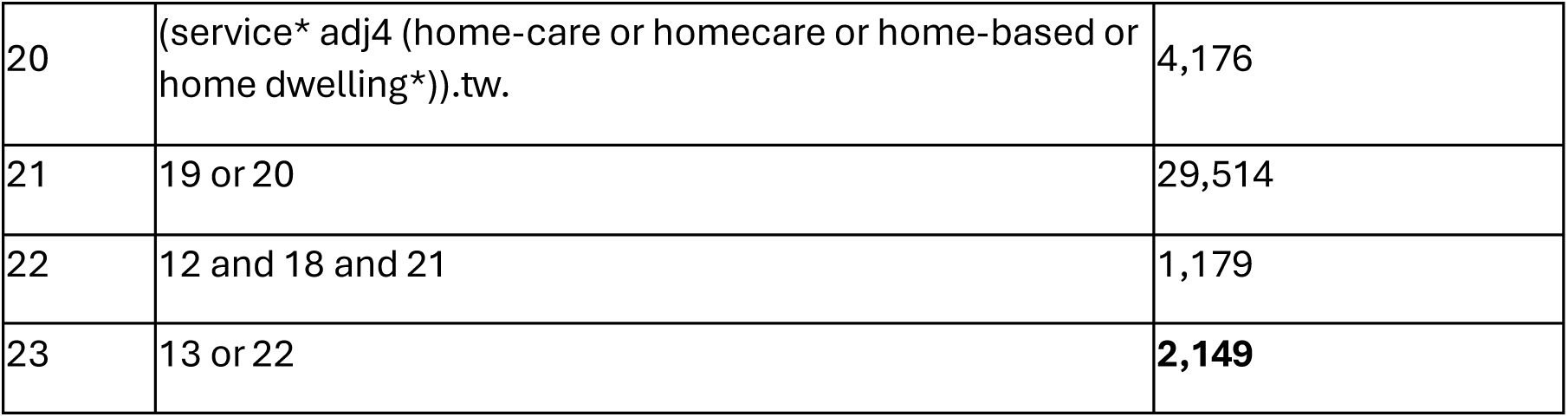

| # | Query | Results from 15 Jan 2025 |
| --- | --- | --- |
| 1 | (re-ablement or reablement or enablement or re-enablement).tw,kw. | 1,096 |
| 2 | (restorative adj4 (care or home care or homecare or programme* or program* or service*)).tw,kw. | 1,068 |
| 3 | "stay active at home".tw. | 10 |
| 4 | home-based rehabilitation.tw. | 664 |
| 5 | 1 or 2 or 3 or 4 | 2,800 |
| 6 | exp aged/ | 3,632,477 |
| 7 | exp Health Services for the Aged/ | 18,343 |
| 8 | exp geriatrics/ | 32,069 |
| 9 | exp frailty/ | 12,430 |
| 10 | (aged or aging or ageing or geriatric* or elder* or old or older or frail* or senior citizen*).tw. | 2,889,382 |
| 11 | *nursing homes/ | 26,771 |
| 12 | 6 or 7 or 8 or 9 or 10 or 11 | 5,704,558 |
| 13 | 5 and 12 | 1,050 |
| 14 | *"Activities of Daily Living"/ | 24,730 |
| 15 | ("activities of daily living" or ADL or iADL).tw. | 43,012 |
| 16 | (independence or independent functioning or independent living).tw. | 59,417 |
| 17 | (daily functioning or daily activities).tw. | 24,105 |
| 18 | 14 or 15 or 16 or 17 | 133,083 |
| 19 | *Home Care Services/ | 27,372 |
| 20 | (service* adj4 (home-care or homecare or home-based or home dwelling*)).tw. | 4,176 |
| 21 | 19 or 20 | 29,514 |
| 22 | 12 and 18 and 21 | 1,179 |
| 23 | 13 or 22 | **2,149** |

Database: Embase Classic+Embase <1947 to 2025 January 14>

| # | Query | Results from 15 Jan 2025 |
| --- | --- | --- |
| 1 | (re-ablement or reablement or enablement or re-enablement).tw,kw. | 1,450 |
| 2 | (restorative adj4 (care or home care or homecare or programme* or program* or service*)).tw,kw. | 1,198 |
| 3 | "stay active at home".tw. | 9 |
| 4 | home-based rehabilitation.tw. | 842 |
| 5 | 1 or 2 or 3 or 4 | 3,453 |
| 6 | exp aged/ | 4,185,234 |
| 7 | exp geriatrics/ | 61,885 |
| 8 | exp frailty/ | 34,833 |
| 9 | (aged or aging or ageing or geriatric* or elder* or old or older or frail* or senior citizen*).tw. | 4,212,210 |
| 10 | exp nursing home/ | 68,566 |
| 11 | 6 or 7 or 8 or 9 or 10 | 7,206,435 |
| 12 | 5 and 11 | 1,286 |
| 13 | *daily life activity/ | 17,762 |
| 14 | ("activities of daily living" or ADL or iADL).tw. | 62,164 |
| 15 | (independence or independent functioning or independent living).tw. | 81,881 |
| 16 | (daily functioning or daily activities).tw. | 35,774 |
| 17 | 13 or 14 or 15 or 16 | 179,185 |
| 18 | *home care/ | 37,399 |
| 19 | (service* adj4 (home-care or homecare or home-based or home dwelling*)).tw. | 5,146 |
| 20 | 18 or 19 | 40,231 |
| 21 | 11 and 17 and 20 | 1,375 |
| 22 | 12 or 21 | 2,584 |
| 23 | conference abstract.pt. | 5,338,860 |
| 24 | 22 not 23 | **2,268** |

Database: CINAHL (EBSCO) 15.01.2025

| # | Query | Results from 15 Jan 2025 |
| --- | --- | --- |
| 1 | TI ( re-ablement or reablement or enablement or re-enablement ) OR AB ( re-ablement or reablement or enablement or re-enablement ) | 747 |
| 2 | TI ( restorative N4 (care or home care or homecare or programme* or program* or service*) ) OR ( restorative N4 (care or home care or homecare or programme* or program* or service*) ) | 543 |
| 3 | TI "stay active at home" OR AB "stay active at home" | 8 |
| 4 | TI "home based rehabilitation" OR AB "home based rehabilitation" | 331 |
| 5 | OR 1-4 | 1607 |
| 6 | (MH "Aged+") | 978,444 |
| 7 | (MH "Geriatrics+") | 7,103 |
| 8 | (MH "Health Services for Older Persons") | 7,306 |
| 9 | TI ( aged or aging or ageing or geriatric* or elder* or old or older or frail* or "senior citizen*" ) AND AB ( aged or aging or ageing or geriatric* or elder* or old or older or frail* or "senior citizen*" ) | 145,135 |
| 10 | (MH "Nursing Homes+") | 31,980 |
| 11 | OR 6-10 | 1,035,069 |
| 12 | 5 AND 11 | 514 |
| 13 | (MM "Activities of Daily Living") | 12,646 |
| 14 | TI ( "activities of daily living" or ADL or iADL ) OR AB ( "activities of daily living" or ADL or iADL ) | 21,760 |
| 15 | TI ( independence or "independent functioning" or "independent living" ) OR AB ( independence or "independent functioning" or "independent living" ) | 24,784 |
| 16 | TI ( "daily functioning" or "daily activities" ) OR AB ( "daily functioning" or "daily activities" ) | 9,566 |
| 17 | OR 13-16 | 59,302 |
| 18 | (MH "Home Health Care+") | 52,434 |
| 19 | TI ( service* N4 (home-care or homecare or home-based or "home dwelling*") ) OR AB ( service* N4 (home-care or homecare or home-based or "home dwelling*") ) | 3,269 |
| 20 | 18 OR 19 | 53,558 |
| 21 | 11 AND 17 AND 20 | 1,214 |
| 22 | 12 OR 21 | **1,650** |

Database: CINAHL (EBSCO) 15.01.2025

| # | Query | Results from 15 Jan 2025 |
| --- | --- | --- |
| 1 | (re-ablement or reablement):ti,ab,kw (Word variations have been searched) | 56 |
| 2 | (restorative NEAR/4 (care or home care or homecare or programme* or program* or service*)):ti,ab,kw (Word variations have been searched) | 343 |
| 3 | (("stay active at home")):ti,ab,kw (Word variations have been searched) | 8 |
| 4 | (("home-based rehabilitation")):ti,ab,kw (Word variations have been searched) | 471 |
| 5 | OR 1-4 | 862 |
| 6 | MeSH descriptor: [Aged] explode all trees | 283,161 |
| 7 | MeSH descriptor: [Geriatrics] explode all trees | 302 |
| 8 | MeSH descriptor: [Frailty] explode all trees | 877 |
| 9 | MeSH descriptor: [Nursing Homes] explode all trees | 2140 |
| 10 | (aged or aging or ageing or geriatric* or elder* or old or older or frail* or "senior citizen" or "senior citizens"):ti,ab,kw (Word variations have been searched) | 1,030,043 |
| 11 | OR 6-10 | 1,030,246 |
| 12 | 5 AND 11 | 495 |
| 13 | MeSH descriptor: [Activities of Daily Living] explode all trees | 13,589 |
| 14 | ("activities of daily living" or ADL or iADL):ti,ab,kw (Word variations have been searched) | 18,478 |
| 15 | (independence or "independent functioning" or "independent living"):ti,ab,kw (Word variations have been searched) | 108,854 |
| 16 | ("daily functioning" or "daily activities"):ti,ab,kw (Word variations have been searched) | 9,445 |
| 17 | OR 13-16 | 136,697 |
| 18 | MeSH descriptor: [Home Care Services] explode all trees | 3,179 |
| 19 | (service* NEAR/4 (home-care or homecare or home-based or "home dwelling" or "home dwellings")):ti,ab,kw (Word variations have been searched) | 3,096 |
| 20 | 18 OR 19 | 3,550 |
| 21 | 11 AND 17 AND 20 | 738 |
| 22 | 12 AND 21 | **1170** |

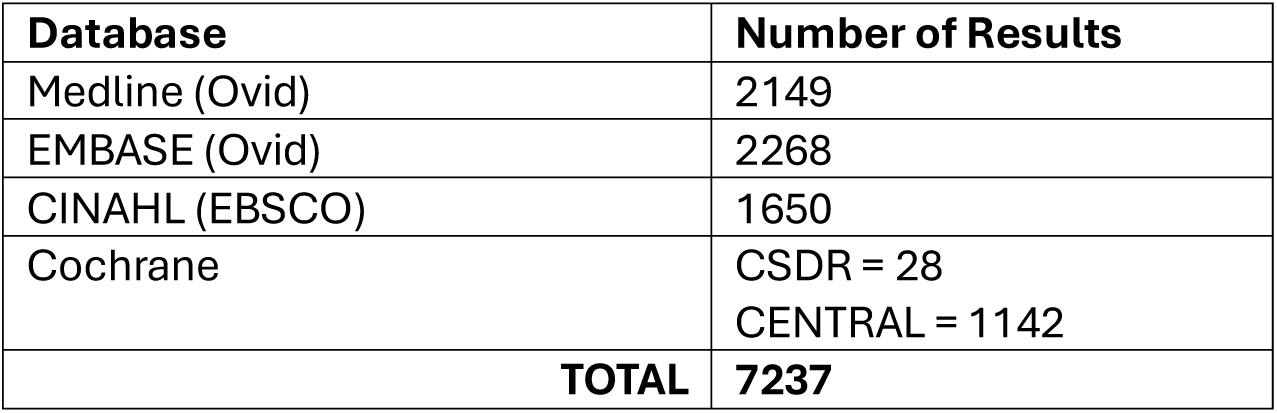

### 9.2 Quality appraisal

#### 9.2.1 Critical appraisal toolkit (CAT) Evidence Summary Table for outcomes of clinical effectiveness

9.2 Quality appraisal 9.2.1 Critical appraisal toolkit (CAT) Evidence Summary Table for outcomes of clinical effectiveness
| Study | Critical appraisal tool items |  |  |  |  |  |  |  |  |  |  |  | Overall quality* |
| --- | --- | --- | --- | --- | --- | --- | --- | --- | --- | --- | --- | --- | --- |
|  | 1 | 2 | 3 | 4 | 5 | 6 | 7 | 8 | 9 | 10 | 11 | 12 |  |
| Lewin & Vandermeulen, 2010 | S | M | W | S | M | M | W | W | M | S | S | S | Medium |
| Lewin et al., 2013a | S | S | M | W | M | W | M | W | M | S | S | S | Low |
| Lewin et al., 2014 | S | S | M | W | M | W | M | M | M | S | S | S | Low |
| Lewin et al., 2013b | S | M | M | S | M | W | W | M | M | S | S | S | Medium |
| Lewin et al., 2016 | S | M | M | S | M | M | M | W | M | S | S | S | Medium |
| Tuntland et al., 2015 | S | S | S | M | M | S | M | S | S | S | S | S | High |
| Langeland et al., 2019 | S | M | W | S | M | S | W | M | W | S | S | S | Medium |
| Parsons et al., 2013 | S | M | S | S | S | S | S | W | S | S | S | S | Medium |
| Parsons et al., 2017 | S | W | W | W | W | S | W | W | W | S | M | W | Low |
| Schmidt et al., 2024 | S | M | W | M | M | S | S | W | W | S | S | S | Medium |
| Winkel et al., 2015 | S | S | W | S | W | S | W | W | W | W | S | W | Low |
| Beresord et al., 2019 | S | S | S | S | S | S | W | W | W | S | S | S | Medium |
| Glendinning et al., 2010 | S | S | W | S | W | S | W | S | M | S | S | S | Medium |
| Whitehead et al., 2016 | S | M | S | S | M | S | S | S | S | S | M | W | Medium |
| Tinetti et al., 2002 | S | S | S | S | W | W | S | S | S | S | S | S | Medium |
| Tinetti et al., 2012 | S | S | M | M | W | W | M | S | M | S | S | S | Medium |

#### 9.2.2 JBI critical appraisal checklist for economic evaluations

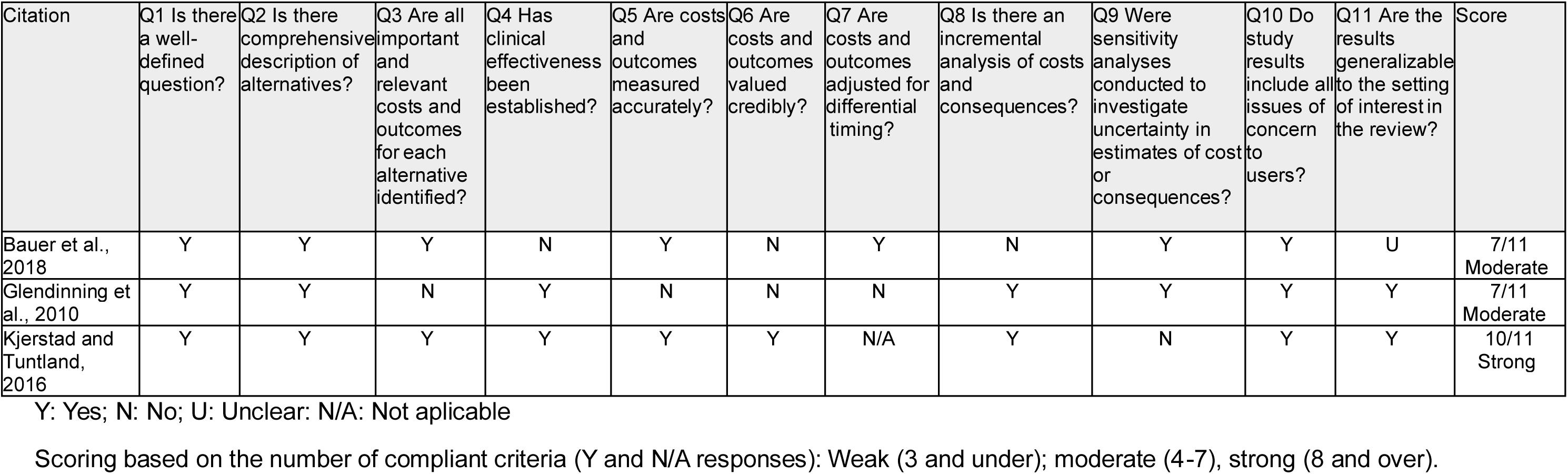

| Citation | Q1 Is there a well-defined question? | Q2 Is there a comprehensive description of alternatives? | Q3 Are all important and relevant costs and outcomes for each alternative identified? | Q4 Has clinical effectiveness been established? | Q5 Are costs and outcomes measured accurately? | Q6 Are costs and outcomes valued credibly? | Q7 Are costs and outcomes adjusted for differential timing? | Q8 Is there an incremental analysis of costs and consequences? | Q9 Were sensitivity analyses conducted to investigate uncertainty in estimates of cost or consequences? | Q10 Do study results include all issues of concern to users? | Q11 Are the results generalizable to the setting of interest in the review? | Score |
| --- | --- | --- | --- | --- | --- | --- | --- | --- | --- | --- | --- | --- |
| Bauer et al., 2018 | Y | Y | Y | N | Y | N | Y | N | Y | Y | U | 7/11<br>Moderate |
| Glendinning et al., 2010 | Y | Y | N | Y | N | N | N | Y | Y | Y | Y | 7/11<br>Moderate |
| Kjerstad and Tuntland, 2016 | Y | Y | Y | Y | Y | Y | N/A | Y | N | Y | Y | 10/11<br>Strong |

Y: Yes; N: No; U: Unclear; N/A: Not applicable
Scoring based on the number of compliant criteria (Y and N/A responses): Weak (3 and under); moderate (4-7), strong (8 and over).

### 9.3 Reasons for exclusion during full text screening

| Citation: | Reason for exclusion: |
| --- | --- |
| Burton et al., 2013 | Wrong intervention - not reablement. The intervention is a physiotherapist-led exercise programme. |
| Rooijackers et al., 2021 | Wrong intervention - not reablement. The intervention is staff training on enablement ethos, cultural change in care home. |
| Gill et al., 2004 | Wrong intervention - not reablement. The intervention is a prehabilitation program that focuses on improving underlying impairments. |
| Todo et al., 2021 | Wrong intervention - not reablement. The intervention is home based rehabilitation including exercise, practicing ADLs, improving the home environment, and carer support. |
| Chiang et al., 2020 | Wrong country. Study conducted in a non-OECD country (Taiwan). |
| Rooijackers et al., 2021 | Wrong intervention - not reablement. The intervention is staff training on enablement ethos, cultural change in care home |
| Rooijackers et al., 2021 | Wrong intervention - not reablement. The intervention is staff training on enablement ethos, cultural change in care home |
| Kjernsholen et al., 2024 | Duplicate study - duplicate paper of an included study |
| Kjerstad et al., 2016 | Duplicate study - duplicate paper of an included study |
| Talley et al. 2014 | Wrong intervention - not reablement. Restorative care was not multidisciplinary, did not involve AHPs, was not goal-oriented or person centred. |
| Walters et al, 2017 | Wrong intervention. Study is not time limited – intervention was intended to be delivered over 6 months in approximately six appointments of 30–60 minutes |
| Fahlström et al., 2017 | Wrong intervention - not reablement. Intervention was a falls prevention exercise programme and was not time-limed (intervention lasted 5 months). |
| Han et al., 2020 | Wrong population. Study assessed home based ADL training for stroke patients. |
| Lampe et al, 2022 | Wrong intervention - not reablement – intervention was intended support and assist with consultations, mediate and promote local offers for exercise and provide personal assistance. |
| Kjernsholen 2023 | Wrong intervention - Intervention length is over 12 weeks. The reablement sessions lasted for 45 minutes, 3 times a week for a maximum of 16 weeks. |
| Hatori et al, 2019 | Wrong intervention. Intervention was over a 5-month period. |
| Jacobi et al., 2020 | Wrong outcomes. Did not assess effectiveness, cost-effectiveness, costs, resource use or implementation, |
| Gitlin et al., 2006 | Wrong intervention - not reablement. The 6-month intervention aimed to reduce functional difficulties, fear of falling, and home hazards and enhance self-efficacy and adaptive coping in older adults with chronic conditions. |
| Robert et al, 2024 | Wrong intervention. Intervention on restorative and rehabilitative care and was delivered over a 6-month period post-discharge. |
| Nadash and Feldman, 2003 | Wrong publication type – not a primary study. |
| Frost et al, 2023 | Wrong intervention. Intervention was delivered over a 6-month period. |
| King et al., 2012 | Wrong intervention. No therapist involvement and the study involved paid carer training to deliver the intervention to clusters of older people. The intervention was not time-limited i.e. regular visits from paid carer and coordinator visits every 3 months, and a full re-assessment at 12 months. |
| Grönstedt et al., 2012 | Wrong intervention. Assesses a physical activity and exercise programme |
| Parsons et al., 2012 | Wrong publication type – not a primary study. |
| Parsons et al., 2020 | Wrong intervention - not reablement – intervention is post-acute care following injury |
| Senior et al., 2014 | Wrong intervention. Intervention includes rehabilitation delivered by home treatment teams and is not time-limited. |
| Assander et al., 2022 | Wrong outcomes – did not assess effectiveness or cost-effectiveness |
| Slater and Hasson, 2018 | Wrong study design – this was a descriptive study (not analytical). |

### 9.4 Summary of interventions described in the included studies using the TIDieR checklist (Hoffman et al., 2024)

9.4 Summary of interventions described in the included studies using the TIDieR checklist (Hoffman et al., 2024).
| Langeland et al., 2019 |  |
| --- | --- |
| **1. Brief name** | 10-week multicomponent reablement intervention |
| **2. Why** | Goals:<br>To restore and increase daily activities, physical function, health-related quality of life and coping as a sense of coherence.<br>To establish a dialog to identify activities that the individual perceives as being meaningful to improve. The intervention was targeted towards achieving these activity goals and therefore varied in content due to individual tailoring of goals, length and intensity of the intervention. |
| **3. What (materials)** | The Canadian Occupational Performance Measure (COPM) was used to measure participants' performance of daily activities and satisfaction with their performance and included three occupational performance areas: self-care, productivity and leisure. The Short Physical Performance Measure Battery (SPPB) to measure physical function, the EuroQol-5 Dimension 5 level version (EQ-5D-5L) to measure health-related quality of life, and the Sense of Coherence Questionnaire (SOC-13) to measure coping related to experiences of coherence. |
| **4. What (procedures)** | A member of the multi-disciplinary team completed the COPM interview in the participants' home. The participant then states up to five goals that they feel are essential. An individual rehabilitation plan was developed to align with these goals. The content of the intervention varied across participants due to the Individual tailoring. However, some of the basic features included the following: delivered up to a maximum of 10 weeks; person-centred, resource-oriented and interdisciplinary approach; promoting self-management and self-training, training in daily activities, adaptations and exercise programmes. Multiple home visits were conducted by health care personnel to deliver daily training to build confidence and relearn skills. |
| **5. Who provided** | An occupational therapist, physiotherapist, or nurse conducted the initial interview using the Canadian Occupational Performance Measure. Multiple home visits by health care personnel to deliver daily training to build confidence and relearn skills. |
| **6. How** | Face-to-face home visits. Multidisciplinary team with shared goals collaborated with participants throughout the intervention period. Daily activity training to build confidence and relearn tasks with a focus on personal hygiene, managing stairs, cleaning, writing, walking, family visits and daily exercise routines. |
| **7. Where** | Participant homes across 47 Norwegian municipalities. |
| **8. When and how much** | Reablement lasting 4-10 weeks. Most participants received between three and five visits per week for 4-6 weeks; however, duration/intensity of the variation varied across participants based on their individual needs/goals. |
| **9. Tailoring** | The intervention varied in content due to individual tailoring of goals, length and intensity of the intervention. |
| **10. Modifications** | Not reported. |
| **11. How well (planned)** | Intervention adherence or fidelity was not assessed. |
| **12. How well (actual)** | Intervention adherence or fidelity was not assessed. |
| **Tuntland et al., 2015** |  |
| **1. Brief name** | 10-week multicomponent reablement intervention |
| **2. Why** | Goals:<br>To investigate the effectiveness of reablement in home-dwelling older adults compared with usual care in relation to daily activities, physical functioning, and health-related quality of life.<br>To enhance independence in daily living activities that are important to the participants.<br>To enable people to age in place, be active and participate socially in the society.<br>The intervention was designed in line with the Canadian Model of Occupational Performance and Engagement (CMOP-E) which was used as a theoretical framework in the study. First RCT of reablement to be conducted in Europe to establish effectiveness of reablement compared to usual care. |
| **3. What (materials)** | The Canadian Occupational Performance Measure (COPM) was used to measure participants' performance of daily activities and satisfaction with their performance and included three occupational performance areas: self-care, productivity and leisure. Functional mobility assessed using the Timed Up and Go test. Jamar Dynamometer (hydraulic instrument) to measure grip strength. Health-related quality of life using the COOP/Wonka instrument. |
| **4. What (procedures)** | The initial assessment involved the occupational therapist and physical therapist using the COPM to identify difficulties in daily activities that were perceived important to the participants. Subsequently a rehabilitation plan was developed. The therapists supervised the homecare personnel/assistants around how best to encourage and assist the participants. |
| **5. Who provided** | Initial assessment provided by occupational therapist and physical therapist. Subsequent visits were completed by homecare personnel/assistants with supervision provided by the occupational therapist and physical therapist. |
| **6. How** | Face-to-face home visits. The focus was on supporting participants to perform the daily activities themselves, instead of letting others on their behalf. Individual components included training in daily activities, adaptations to the environment/or activity and exercise regimes. |
| **7. Where** | Participants' homes in one rural municipality in Norway. |
| **8. When and how much** | Maximum reablement period of 3 months. The number of visits depending on the participants' needs/goals. |
| **9. Tailoring** | Intervention was tailored according to the needs and aims of the individual participant therefore the number and type of intervention components varied across participants. |
| **10. Modifications** | Not reported. |
| **11. How well (planned)** | Intervention adherence or fidelity was not assessed. |
| **12. How well (actual)** | Intervention adherence or fidelity was not assessed. |
| **Lewin et al., 2016** |  |
| **1. Brief name** | Home Independence Program Coordinator (HIPC) intervention |
| **2. Why** | Goals:<br>Few Australian care homes employ health professionals who act as care managers under the home independence program (HIP) service model. This study evaluated a HIP service model (see Lewin and Vandermeulen, 2010) in which non-health professional replaced the health professionals as care managers. The aim of restorative care programmes is to reduce the needs for ongoing home care assistance and reduce admissions to residential care, hospital or emergency department visits. |
| **3. What (materials)** | Five outcomes measures: Activities of Daily Living (ADL) and Instrumental Activities of Daily Living (IADL) using a tool based on the modified Barthel Index and the Lawton and Brodie Scale, the Assessment of Quality of Life instrument (AQOL), the Modified Falls Efficacy Scale (MFES), and the Timed Up and Go (TUG).<br>Resource use data was also collected from the organisation's client information system. |
| **4. What (procedures)** | Comprehensive multidimensional assessment; Goal-oriented care planning in partnership with client; Targeted evidence-based interventions to optimize functioning in daily living activities; Minimized face-to-face contact – telephone support and follow-up; Education about self-management, healthy aging, use of medications, and illness/accident prevention strategies; Use of language and patterns of communication that encourage clients and families to participate in all care decisions and which promote their sense of autonomy rather than exerting power or control over the client; Recognition of the importance of the social support aspect of home care services for older people and the need to assist the client to develop other avenues for gaining this support; and Use of local resources – facilitated by a resource file. |
| **5. Who provided** | Training provided to the coordinators was delivered by Senior Allied Health Professionals working within a multidisciplinary HIP team. Non-health care professional delivered the intervention and acted as care managers. |
| **6. How** | Face-to-face home visits. The HIP service delivery model requires a multidisciplinary team of health professionals (namely occupational therapists, physiotherapists [physios] and nurses) to work in an interdisciplinary way, with a non-health professional acting as the care manager. The care manager implemented assessments, goal setting, care-planning, and worked with the client to implement interventions/strategies relevant to the individual's needs and goals. |
| **7. Where** | Participants homes in Perth, Australia. |
| **8. When and how much** | Multiple home visits until participants achieved their goals or up to 12 weeks. |
| **9. Tailoring** | The types of interventions included in an individual's care plan were based on an individual's goals, their capabilities, and the types of difficulties they were experiencing. |
| **10. Modifications** | This study evaluated a HIP service model in which non-health professional replaced the health professionals as care managers. Although the intervention was time-limited (up to 12 weeks), in a minority of cases, some participants received the intervention for longer than 12 weeks if they were progressing well to achieving their goals. |
| **11. How well (planned)** | Not assessed in this paper. |
| **12. How well (actual)** | Not assessed in this paper. |
| **Lewin and Vandermeulen, 2010** |  |
| **1. Brief name** | Home Independence Programme with health professionals as care managers intervention |
| **2. Why** | Goal:<br>HIP was developed as an early intervention programme directed at optimising functioning, preventing or delaying further functional decline, promoting healthy ageing and encouraging the self-management of chronic diseases. |
| **3. What (materials)** | Activities of Daily Living (ADL) and Instrumental Activities of Daily Living (IADL) using a tool based on the modified Barthel Index and the Lawton and Brodie Scale, the Assessment of Quality of Life instrument (AQOL), the Modified Falls Efficacy Scale (MFES), the Timed Up and Go (TUG), and the Philadelphia Geriatric Morale Scale (PGMS). |
| **4. What (procedures)** | Key components of HIP: Comprehensive multidimensional assessment; Goal-oriented care planning in partnership with client; Targeted evidence-based interventions to optimize functioning in daily living activities; Minimized face-to-face contact – telephone support and follow-up; Education about self-management, healthy aging, use of medications, and illness/accident prevention strategies; Use of language and patterns of communication that encourage clients and families to participate in all care decisions and which promote their sense of autonomy rather than exerting power or control over the client; Recognition of the importance of the social support aspect of home care services for older people and the need to assist the client to develop other avenues for gaining this support; and Use of local resources – facilitated by a resource file.<br>Types of interventions included in HIP included the following: the promotion of active engagement in a range of daily living activities using task analysis and redesign, work simplification and assistive technology where appropriate; strength balance and endurance programmes for improving or maintaining mobility; chronic disease self-management; falls prevention strategies; medication, continence and nutrition management; and the improvement or maintenance of skin integrity |
| **5. Who provided** | Involved a multidisciplinary team (consisting of a nurse, physiotherapist and occupational therapist). One member of the multidisciplinary team worked with the participant to implement assessments, goal setting, care-planning, and worked with the client to implement interventions/strategies relevant to the individual's needs and goals. |
| **6. How** | Paper reports that details of how exactly the service operates is available in the 'How To' HIP user manual (unable to access this manual). |
| **7. Where** | HIP trialled in one metropolitan area of Perth, Australia. |
| **8. When and how much** | Multiple home visits until participants achieved their goals or up to 12 weeks. |
| **9. Tailoring** | The types of interventions included in an individual's care plan were based on an individual's goals, their capabilities, and the types of difficulties they were experiencing. |
| **10. Modifications** | Although the intervention was time-limited (up to 12 weeks), in a minority of cases, some participants received the intervention for longer than 12 weeks if they were progressing well to achieving their goals. |
| **11. How well (planned)** | Not assessed |
| **12. How well (actual)** | Not assessed |
| **Kjerstad and Tutland, 2016** |  |
| **1. Brief name** | 10-week multicomponent reablement intervention |
| **2. Why** | Goals:<br>Reablement has the potential to reduce the expected growth in long-term care expenditure.<br>This study was a cost-effectiveness analysis conducted alongside a randomised control (see Tuntland et al., 2015) to evaluate the cost-effectiveness of a reablement programme compared to usual care in Norway. |
| **3. What (materials)** | The Canadian Occupational Performance Measure (COPM) was used to measure participants' performance of daily activities and satisfaction with their performance and included three occupational performance areas: self-care, productivity and leisure. Functional mobility assessed using the Timed Up and Go test. Jamar Dynamometer (hydraulic instrument) to measure grip strength. Health-related quality of life using the COOP/Wonka instrument. |
| **4. What (procedures)** | The initial assessment involved the occupational therapist and physical therapist using the COPM to identify difficulties in daily activities that were perceived important to the participants. Subsequently a rehabilitation plan was developed. The therapists supervised the homecare personnel/assistants around how best to encourage and assist the participants. |
| **5. Who provided** | Initial assessment provided by occupational therapist and physical therapist. Subsequent visits were completed by homecare personnel/assistants with supervision provided by the occupational therapist and physical therapist. |
| **6. How** | Face-to-face home visits. The focus was on supporting participants to perform the daily activities themselves, instead of letting others on their behalf. Individual components included training in daily activities, adaptations to the environment/or activity and exercise regimes. |
| **7. Where** | Participants' homes in one rural municipality in Norway. |
| **8. When and how much** | Maximum reablement period of 3 months. The number of visits depending on the participants' needs/goals. |
| **9. Tailoring** | Intervention was tailored according to the needs and aims of the individual participant therefore the number and type of intervention components varied across participants. |
| **10. Modifications** | Not reported. |
| **11. How well (planned)** | Intervention adherence or fidelity was not assessed. |
| **12. How well (actual)** | Intervention adherence or fidelity was not assessed. |
| **Lewin et al., 2013a** |  |
| **1. Brief name** | Home Independence Programme with health professionals as care managers intervention |
| **2. Why** | Goals:<br>To test the effectiveness of the Home Independence Program (HIP) in reducing the need for ongoing services. HIP was developed to target older individuals referred for home care, regardless of their referral source, and has been shown to significantly reduce the need for ongoing services and to be effective in improving people's functioning on a range of outcome measures, including measures of functional dependency, confidence and morale. |
| **3. What (materials)** | Primary Assessment Form which includes Activities of Daily Living (ADL) and Instrumental Activities of Daily Living (IADL) scales based on the Modified Barthel Index and the Lawton and Brody Scale with the scoring modified to increase according to the amount of assistance required on a task; the timed up and go; the Modified Falls Efficacy Scale and the Assessment of Quality of Life Scale. |
| **4. What (procedures)** | Initially, individuals have either been referred to HIP directly or invited to participate at initial assessment for home-care services. In brief, the areas of functioning and types of interventions that are incorporated into HIP include the promotion of active engagement in a range of daily living activities using task analysis and redesign, work simplification and assistive technology where appropriate; strength, balance and endurance programmes for improving or maintaining mobility; chronic disease self-management; falls prevention strategies; medication, continence and nutrition management; and the improvement or maintenance of skin integrity. If HIP individuals need continued assistance from a home care service at discharge from HIP, this is arranged. |
| **5. Who provided** | Involved a multidisciplinary team (consisting of a nurse, physiotherapist and occupational therapist). One member of the multidisciplinary team worked with the participant to implement assessments, goal setting, care-planning, and worked with the client to implement interventions/strategies relevant to the individual's needs and goals. |
| **6. How** | Paper reports that details of how exactly the service operates is available in the 'How To' HIP user manual (unable to access this manual). |
| **7. Where** | Participants' home in Perth suburbs. |
| **8. When and how much** | Individuals participate in HIP until they achieve their goals or for up to 12 weeks, whichever occurs first. Sufficient service defined as three HIP visits a week. |
| **9. Tailoring** | Not explicitly reported. |
| **10. Modifications** | Not reported |
| **11. How well (planned)** | Not assessed |
| **12. How well (actual)** | Not assessed |
| **Lewin et al., 2013b** |  |
| **1. Brief name** | HIP and Personal Enablement Program (PEP) intervention |
| **2. Why** | Goals:<br>To compare retrospectively the home care use and cost over a period of nearly 5 years of older individuals who had participated in either of two reablement home care services (one targeted post discharge and the other targeted community-based referrals) or who had received a conventional home care service. |
| **3. What (materials)** | The primary outcome measure was use of home care services at different time points following commencement of the intervention (HIP, PEP, conventional HACC home care). The cumulative cost of home care services was examined as a secondary outcome measure. |
| **4. What (procedures)** | HIP is a short-term, individualized service designed to promote independence and minimize the need for ongoing support services. It is goal-oriented and promotes active engagement in daily living activities using task analysis and redesign, work simplification, and assistive technology. Depending on an individual's goals it may also include: strength, balance, and endurance programs for improving or maintaining function and mobility; chronic disease self-management; falls prevention strategies; medication, continence, and nutrition management; and strategies to assist the individual to reconnect socially.<br>PEP, like HIP, has an enablement focus, but unlike HIP, PEP can provide post-acute nursing if required. PEP also differs from HIP by having an 8-week time limit. |
| **5. Who provided** | Involved a multidisciplinary team (consisting of a nurse, physiotherapist and occupational therapist). One member of the multidisciplinary team worked with the participant to implement assessments, goal setting, care-planning, and worked with the client to implement interventions/strategies relevant to the individual's needs and goals. |
| **6. How** | Paper reports that details of how exactly the service operates is available in the 'How To' HIP user manual (unable to access this manual). |
| **7. Where** | Participants' home in Australia |
| **8. When and how much** | The HIP service usually has a 12-week time limit. PEP has an 8-week time limit. Participants must have received at least a “minimum dose” of intervention, which was defined as three reablement service visits. |
| **9. Tailoring** | HIP is a short-term, individualized service designed to promote independence and minimize the need for ongoing support services. It is goal-oriented and promotes active engagement in daily living activities using task analysis and redesign, work simplification, and assistive technology. |
| **10. Modifications** | Not reported |
| **11. How well (planned)** | Not assessed |
| **12. How well (actual)** | Not assessed |
| **Parsons et al., 2013** |  |
| **1. Brief name** | Restorative home care intervention |
| **2. Why** | Goals:<br>To determine the impact of a restorative model of home care on social support and physical function among community-dwelling older people.<br>This study sought to determine whether provision of restorative home care to a sample of community-dwelling older people in New Zealand would result in an improvement in physical function and social support when compared with a group receiving standard home care. More specifically, the study explored the impact of aligning service delivery through individually tailored activities to support the older person in attaining his/her goals. |
| **3. What (materials)** | Physical function was determined by the Short Physical Performance Battery (SPPB), a validated test comprising an assessment of standing balance, a timed 2.4-m walk test, and a timed test of 5 repetitions of rising from a chair and sitting down. Social support was ascertained by the Duke Social Support Index (DSSI). The assessor used a goal-setting tool (Towards Achieving Realistic Goals in Elders Tool [TARGET]) during the initial assessment process of the participant to establish the aims of the rehabilitation episode. The assessment phase incorporated 2 outcome tools, namely, the Nottingham Extended Activities of Daily Living Scale (NEADL) and the EuroQoL 5D (EQ-5D), into the Support Needs Assessment (SNA) tool which examines a number of areas including cognition, informal caregiver stress, safety, and nutrition. The NEADL was used to quantify an individual's level of functioning, and the EQ-5D is a descriptive tool, measuring health status, that features mental health and pain in the questionnaire. |
| **4. What (procedures)** | The assessor used a goal-setting tool (Towards Achieving Realistic Goals in Elders Tool [TARGET]) during the initial assessment process of the participant to establish the aims of the rehabilitation episode. The assessment phase incorporated 2 outcome tools, namely, the Nottingham Extended Activities of Daily Living Scale (NEADL) and the EuroQoL 5D (EQ-5D), into the SNA tool. After assessment, a long-term goal was identified together with necessary short-term goals through a process of activity breakdown to form a goal ladder. The goal ladder was then passed from the needs assessment agency to the home care organization. |
| **5. Who provided** | The support plan comprised a detailed list of the tasks to be undertaken. This may have included the use of allied health professionals (occupational therapist, physical therapist, speech-language pathologist, dietician) to provide expert guidance in the tasks required to attain a participant's goals. Both groups were assessed by a needs assessor, and the assessment findings formed the basis for the quantity and content of the services to be delivered by the contracted home care organisation. All activities described in the support plans were undertaken by home care aides employed by the home care organizations. |
| **6. How** | The services provided to participants as part of their home care were analysed and categorized as domestic tasks (vacuuming), personal care (showering assistance), shopping (with and/or without the participant), and individualized activities (activities identified specifically for the individual participant). Individualized activities predominantly focused on assisting participants to access the community and were concerned with increasing function outside the home. These activities were individualized walking or exercise programs or other activities aimed at improving functional ability. |
| **7. Where** | Home care in an urban area in New Zealand. |
| **8. When and how much** | Not reported. |
| **9. Tailoring** | Plans/goals were individualized to participants following the assessment phase. |
| **10. Modifications** | Not reported. |
| **11. How well (planned)** | Not reported. |
| **12. How well (actual)** | Not reported. |
| **Parsons et al., 2017** |  |
| **1. Brief name** | Community Flexible Integrated Responsive Support Team (FIRST) intervention |
| **2. Why** | Goals:<br>To establish the effectiveness of restorative home support service on institutional-free survival in frail older people referred for needs assessment. This study describes the intervention and randomised evaluation of a new form of home care, termed Community Flexible Integrated Responsive Support Team (FIRST). |
| **3. What (materials)** | The primary endpoint was death or permanent residential care placement. Secondary endpoints were assessed on the interRAI Home Care (interRAI HC) instrument, this instrument comprises domains of physical function, health, social support and service use. The EuroQoL thermometer visual analogue scale was used to assess current health status. Health service use was collected from the interRAI HC instrument and the regional health board. Informal caregiver health-related quality of life (HRQoL) was assessed by the physical and mental component subscales of the 36-item Short-Form questionnaire (SF-36, a reported survey of health). Informal caregiver experience was evaluated by the Caregiver Reaction Assessment (CRA) Instrument (24 questions with Likert responses, relating to how a caregiver perceives their caring role). |
| **4. What (procedures)** | Targeted assessments for those with complex needs are assessed using the interRAI Home Care Assessment by needs assessors and those with non-complex needs are assessed using the interRAI Contact Assessment. Goal facilitation is integrated into the care plan. Each (distal) goal is broken into (proximal) sub-goals and illustrated using a goal ladder. The distal goal at the top of the ladder is 'where they want to be', the proximal sub-goal at the bottom is 'where they are now' and the rungs of the ladder are 'how they get there'. ADL exercises are incorporated into care plans, for example, instead of standing once and walking to the bathroom, the client would be encouraged to sit and stand multiple times before walking. |
| **5. Who provided** | A registered nurse case manager coordinator undertakes a comprehensive geriatric assessment (CGA) and sets mutually agreed and meaningful short and long-term goals for the older person. Physical activities are facilitated by an unregulated support worker, akin to a healthcare assistant specially trained in rehabilitation, InterRAI Home Care instruments were administered by trained research registered nurses. |
| **6. How** | Physical activities would be conducted by support workers and healthcare assistants. |
| **7. Where** | Home environments in Hamilton, New Zealand. |
| **8. When and how much** | Frequently visiting the older person up to four times daily, seven days a week, including the evening if required. |
| **9. Tailoring** | Plans/goals were individualized to participants. Changes to the delivery of care, for example, if a client deteriorates over a few days, increased visits or time on each visit can be incorporated as needed. Conversely, if the client improves and becomes more independent, inputs can reduce accordingly. |
| **10. Modifications** | Does not specifically report if modifications occurred however, it was stated that changes in care could be increased or reduce dependent on the need of the client. |
| **11. How well (planned)** | Not assessed. |
| **12. How well (actual)** | Not assessed. |
| **Lewin et al., 2014** |  |
| **1. Brief name** | Home Independence Program (HIP) intervention |
| **2. Why** | Goals:<br>To determine whether older individuals who participated in a reablement (restorative) program rather than immediately receiving conventional home care services had a reduced need for ongoing support and lower home care costs over the next 57 months (nearly 5 years). |
| **3. What (materials)** | Data on aged care usage (HACC services, residential aged care approval) and HACC costs; Healthcare usage (ED presentations and unplanned inpatient admissions) and costs; and Total health and home-care costs (sum of 1 and 2). |
| **4. What (procedures)** | HIP is a short-term individualised service designed to promote independence and minimise the need for ongoing support services. It is goal-oriented and promotes active engagement in daily living activities using task analysis and redesign, work simplification and assistive technology. Depending on an individual's goals, it may also |
|  | include strength, balance and endurance programmes for improving or maintaining mobility; chronic disease self-management; falls prevention strategies; medication, continence and nutrition management; and strategies to assist the individual to reconnect socially. |
| **5. Who provided** | Involved a multidisciplinary team (consisting of a nurse, physiotherapist and occupational therapist). One member of the multidisciplinary team worked with the participant to implement assessments, goal setting, care-planning, and worked with the client to implement interventions/strategies relevant to the individual's needs and goals. |
| **6. How** | Paper reports that details of how exactly the service operates is available in the 'How To' HIP user manual (unable to access this manual). |
| **7. Where** | Participant's home in Australia. |
| **8. When and how much** | The service usually has a 12-week time limit and participants must have had at least 3 visits. |
| **9. Tailoring** | HIP is a short-term individualised service so the intervention can differ depending on the requirements of the participant. |
| **10. Modifications** | Not reported. |
| **11. How well (planned)** | Not assessed |
| **12. How well (actual)** | Not assessed |
| **Schmidt et al, 2024** |  |
| **1. Brief name** | Short-Term Restorative Care program |
| **2. Why** | Evaluation of whether the programme delivers its aims |
| **3. What (materials)** | Data were collected from an aged care provider (BallyCara Ltd) located in Australia |
| **4. What (procedures)** | This study reported descriptives of the population that undertook the program and the activities completed by participants during the program, and investigated if the program is associated with changes across four domains of function (Modified Barthel Index, Lower Extremity Functional Scale, Upper Extremity Functional Index and Berg Balance Scale). |
| **5. Who provided** | MDT including both a physician and AHP led the intervention and was delivered by AHPs. The Short-Term Restorative Care (STRC) Programme is an innovative flexible care programme that provides early intervention care which aims to optimise the functioning and independence of older people and reverse and/or slow functional decline. |
| **6. How** | STRC is delivered in the form of a tailored, multidisciplinary package of services which includes the provision of services and assistance such as physiotherapy, social work, nursing support, personal care and the provision of assistive technologies. It aims to enable older people to regain independence and autonomy rather than commencing long term aged care prematurely. |
| **7. Where** | At home, at residential care home, or both. |
| **8. When and how much** | The participants underwent sessions with several allied health professions multiple times per week, depending on the goals of the participant. Up to 8 weeks in total. |
| **9. Tailoring** | It is a programme tailored to each individual |
| **10. Modifications** | None reported |
| **11. How well (planned)** | Not assessed |
| **12. How well (actual)** | Not assessed |
| **Winkel et al, 2014** |  |
| **1. Brief name** | Copenhagen Municipality reablement Program |
| **2. Why** | When older adults experience a decrease in functional independence including decreased ability to perform activities of daily living (ADL) tasks, rehabilitation services are required. The aim of this pilot study was to evaluate if a home-based reablement program influenced the ADL ability of older adults |
| **3. What (materials)** | Activities of Daily Living (ADL) and ADL-Interview (ADL-I) was conducted to measure individualised goals and ADL task performance. |
| **4. What (procedures)** | A referral officer screened potential participants for eligibility. One purpose was to determine the types of ADL tasks, that participants wanted to work on to gain autonomy. Then, an occupational therapist and a home carer met with the participant at the first visit at home. The role of the occupational therapist was to further specify ADL task performance problems of relevance to the participant, set goals and plan and adjust interventions during the program. The occupational therapist, therefore, performed an ADL-I identified tasks that the participant reported as difficult to perform, and then focused on those specific tasks that the participant found meaningful to work on. |
| **5. Who provided** | Home carer supervised by an occupational therapist. |
| **6. How** | The reablement program consisted of three 1-h home visits by an occupational therapist in combination with sessions of 45 min per week for a period of 12 weeks at home by home carers. If appropriate the reablement sessions were split into smaller parts. The therapist and the home carer met with the participant at all three home visits. The reablement approach was based on principles to support the participant's sense of coherence and feeling of self-management. Thus, the participant was encouraged to pursue his/her goals and was supported and stimulated to participate throughout the reablement process. |
| **7. Where** | Home visits in two local districts in Copenhagen |
| **8. When and how much** | Three 1-h home visits by an occupational therapist in combination with sessions of 45 min per week for a period of 12 weeks |
| **9. Tailoring** | Based on the interview and the performance analysis, the occupational therapist formulated goals for the reablement intervention together with the participant and the home carer. |
| **10. Modifications** | ADL-Interviews identified tasks that the participant reported as difficult to perform, and then focused on those specific tasks that the participant found meaningful to work on. |
| **11. How well (planned)** | Intervention adherence or fidelity was not assessed. |
| **12. How well (actual)** | Intervention adherence or fidelity was not assessed. |
| **Beresford et al, 2019** |  |
| **1. Brief name** | Models of English reablement interventions |
| **2. Why** | To describe reablement services in England and develop a service model typology; to conduct a mixed-methods comparative evaluation of service models investigating outcomes, factors that have an impact on outcomes, costs and cost-effectiveness, and user and practitioner experiences. |
| **3. What (materials)** | Health-related quality of life (EQ-5D-5L), Social care-related quality of life (ASCOT SCT-4), Mental health (GHQ-12), Functional status (Barthel Index), Functional ability (Nottingham Extended Activities of Daily Living scale [NEADL]), resource use (Services and Care Pathway Questionnaire [SCPQ]). |
| **4. What (procedures)** | Three English social care reablement services, each representing a different models of service delivery. In England, reablement comprises an assessment by a specialist practitioner during which person-centred goals are co-created with the service user. This is followed by a time-limited period (typically 4–6 weeks) in which trained workers conduct home visits in order to support the achievement of these goals through the regaining of functional skills and/or identifying new ways of carrying out their activities of daily living. |
| **5. Who provided** | Four distinct patterns of staffing were identified within a cluster analysis which aimed to understand the different patterns of staffing and skill mix in the teams. This included, reablement with occupational therapy, homecare reablement, multidisciplinary reablement and reablement workers. |
| **6. How** | A range of reablement services and inputs across three sites. |
| **7. Where** | Work package two of this study focused on three research sites in England. |
| **8. When and how much** | Interventions ranging from 4 to 6 weeks |
| **9. Tailoring** | Not stated |
| **10. Modifications** | Not stated |
| **11. How well (planned)** | Not stated |
| **12. How well (actual)** | Process evaluation involving staff and services users revealed that service users had a limited understanding of reablement. The merging of reablement and assessment functions, and the lack of other in-house social care provision for older people increased the caseload volume for assessors. The importance of flexibility in the timing and duration of visits, and an expectation that this would reduce over the course of reablement, was noted. Some believed that this was not sufficiently specified, or incentivised, in contracts with outsourced providers. Service users believed that positive outcomes had been achieved through the skills and input of the reablement workers. |
| **Tinetti et al., 2012** |  |
| **1. Brief name** | Restorative Model of Posthospital Home Care |
| **2. Why** | To compare readmissions of Medicare recipients of usual domiciliary care and a matched group of recipients of a restorative model of home care. |
| **3. What (materials)** | Outcome and Assessment Information Set (OASIS) version B is used to record admission to home healthcare services, transfer to another site of care (e.g., hospital), resumption of care (after a hospital stay), follow-up (at least 60 days), discharge from home health care, and death. |
| **4. What (procedures)** | The restorative care model was based on principles adapted from geriatric medicine, nursing, rehabilitation, goal attainment, chronic care management, and behavioural change theory. |
| **5. Who provided** | The agency's nursing, physical therapy, occupational therapy, and home health aide (HHA) staff provided care through all six offices under the approval of participants' physicians |
| **6. How** | A multidisciplinary work group of expert home care clinicians familiarised staff with the goal of optimising patient function, developed methods to assess ADLs and quantify increments of improvement, and focused staff on developing individual functional goals with patients and linking interventions to patients' goals. |
| **7. Where** | Homecare |
| **8. When and how much** | Not stated |
| **9. Tailoring** | Functional goals were developed with patients and interventions were linked to patients' goals. |
| **10. Modifications** | Not stated |
| **11. How well (planned)** | Intervention adherence or fidelity was not assessed. |
| **12. How well (actual)** | Intervention adherence or fidelity was not assessed. |
| **Tinetti et al., 2002** |  |
| **1. Brief name** | Restorative care intervention |
| **2. Why** | Illness and hospitalisation often trigger functional decline among older persons. Home care services implemented for functional decline provide an opportunity to intervene to improve outcomes. |
| **3. What (materials)** | All descriptive and outcome data were ascertained using the OASIS-B. A self-care progress report helped to clarify goals, establish a baseline, standardize the assessment of patients, clarify care responsibility across the multiple providers and track patients incremental progress towards reaching those goals. |
| **4. What (procedures)** | All care was provided by the home care agency's nursing and physical and occupational therapy, and home health aide staff. The restorative care model was based on the following principals: nursing, rehabilitation and goal attainment. The treatment plan included various combination of exercise and training; behavioural changes; environmental adjustments and adaptive equipment; counselling and support; training and education of patient, family and friends and medication adjustments. |
| **5. Who provided** | Home care agency's nursing and physical and occupational therapy, and home health aide staff |
| **6. How** | The treatment plan included various combination of exercise and training; behavioural changes; environmental adjustments and adaptive equipment; counselling and support; training and education of patient, family and friends and medication adjustments. |
| **7. Where** | Home care |
| **8. When and how much** | Implementation occurred over 3 months. |
| **9. Tailoring** | The establishment of goals was based on input from patient, family and home care staff, and agreement among this group on the process for reaching these goals. |
| **10. Modifications** | None reported. |
| **11. How well (planned)** | Not assessed. |
| **Whitehead et al., 2016** |  |
| **1. Brief name** | Short-Term Restorative Care Program (STRC) |
| **2. Why** | Although studies have suggested that there are reductions in the amount of homecare provided following reablement in comparison to traditional homecare, there are outstanding questions about the optimum model of service delivery: one such aspect is occupational therapy provision. |
| **3. What (materials)** | Questionnaires (EQ-5D, SF-36, NEADL, ASCOT) and screening of patient recruitment and intervention logs. |
| **4. What (procedures)** | A targeted Activities of Daily Living (ADL) programme, delivered by an occupational therapist incorporating goal setting, teaching/practising techniques, equipment/adaptations and provision of advice/support. This was in addition to usual care. |
| **5. Who provided** | Homecare reablement provided by reablement workers (social care workers) under the direction of a reablement care team leader (social care team manager). Intervention group had the additional ADL programme managed and delivered by an occupational therapist. |
| **6. How** | The aim of the enhanced programme was to maximise independence in ADL activities including, but not limited to, washing, dressing, bathing and showering, feeding, indoor mobility, transfers, stair mobility, toileting, meal preparation and kitchen activities, outdoor mobility and community access. |
| **7. Where** | Home care. |
| **8. When and how much** | Delivered for six weeks. |
| **9. Tailoring** | The programme was agreed with the participant and individually tailored to their needs. It included the following: goal setting using the TARGET; practising activities, and/or a graded process of re-learning and building the skills to manage ADL independently; equipment provision and environmental or activity modification; and case management involving advice to the person and their support network. |
| **10. Modifications** | None stated. |
| **11. How well (planned)** | Intervention adherence or fidelity was not assessed. |
| **12. How well (actual)** | Intervention adherence or fidelity was not assessed. |
| **Bauer et al., 2018** |  |
| **1. Brief name** | Home-care reablement schemes |
| **2. Why** | The study was carried out to inform recommendations of a national guideline. |
| **3. What (materials)** | Costs included those of the intervention, home care and hospital admission. Costs identified from previously published evidence. |
| **4. What (procedures)** | Model parameters for the relative effects of difference in service use were taken from the Australian randomised controlled trial (RCT) (Lewin et al., 2013a). |
| **5. Who provided** | An inter-disciplinary health and social care team carried out comprehensive multidimensional assessment. |
| **6. How** | Multidimensional assessment and goal-oriented care planning in partnership with the person who used service and their family and carers. |
| **7. Where** | Home care. |
| **8. When and how much** | Unclear, but model cycles ran for 1 year each time. |
| **9. Tailoring** | Not reported. |
| **10. Modifications** | Not reported. |
| **11. How well (planned)** | Intervention adherence or fidelity was not assessed. |
| **12. How well (actual)** | Intervention adherence or fidelity was not assessed. |
| **Glendinning et al., 2010** |  |
| **1. Brief name** | Five English home care reablement services |
| **2. Why** | This study investigated the effectiveness and cost-effectiveness of home care re-ablement services compared to traditional home care services. |
| **3. What (materials)** | Questionnaires (EQ-5D and ASCOT) and Client Service Receipt Inventory monitoring health and social care resource use. |
| **4. What (procedures)** | Individual outcomes (self-perceived health, perceived quality of life, health-related quality of life and social care outcomes), social care service use (details of all social care services used by trial participants completed by care providers), NHS service and voluntary organisations service use (self-reported service use). |
| **5. Who provided** | Five different services described. Variety in care provision teams. |
| **6. How** | Five different services described. Variety in care delivered. |
| **7. Where** | Home care. |
| **8. When and how much** | Six weeks, with some able to be extended to 8 weeks if required. |
| **9. Tailoring** | Not explicitly reported but intervention was person centred and individually tailored. |
| **10. Modifications** | Not reported. |
| **11. How well (planned)** | Intervention adherence or fidelity was not assessed. |
| **12. How well (actual)** | Intervention adherence or fidelity was not assessed. |

## The Health and Care Research Wales Evidence Centre

Our dedicated team works together with Welsh Government, the NHS, social care, research institutions and the public to deliver vital research to tackle health and social care challenges facing Wales.

Funded by Welsh Government, through Health and Care Research Wales, the Evidence Centre answers key questions to improve health and social care policy and provision across Wales.

Along with our collaborating partners, we conduct reviews of existing evidence and new research, to inform policy and practice needs, with a focus on ensuring real-world impact and public benefit that reaches everyone.

**Director:** Professor Adrian Edwards

**Associate Directors:** Dr Alison Cooper, Dr Natalie Joseph-Williams, Dr Ruth Lewis

## Footnotes

1 **Allied Health Professions**: include art therapists, drama therapists, music therapists, podiatrists, dietitians, occupational therapists, orthoptists, prosthetists and orthotists, paramedics, physiotherapists, speech and language therapists, psychologists.

